# Vertebral Augmentation for Symptomatic Vertebral Hemangiomas: A Systematic Review and Meta-analysis of Pain Relief, Cement Leakage, and Recurrence

**DOI:** 10.64898/2026.08.18.26360715

**Authors:** Farzan Fahim, Melika Javani, Farzin Mohammad Moradi, Amirmahdi Mojtahedzadeh, Afarinesh Hasheminejad, Ali Khorram, Mahtab Karimi, Mahsa Faramin Lashkarian, Alireza Hosseini Nejad, Fatemeh Eskandari, Zahra Mohammadi, Amirhossein Rastegar, Sarina Simabi, Reza Yazdanpanah, Alireza Zali

## Abstract

**Background:** Vertebroplasty and balloon kyphoplasty are used for symptomatic vertebral hemangiomas, although comparative evidence is limited. We summarized pain relief, cement leakage, and recurrence after vertebral augmentation and assessed whether direct comparison of the two techniques was feasible.

**Methods:** Five databases were searched from inception to January 2, 2026, with an update on July 5, 2026. Because only one small cohort directly compared vertebroplasty with kyphoplasty, outcomes were pooled as single-arm proportions or, for early pain change, as a mean difference using random-effects models. Prespecified subgroup, sensitivity, small-study effect, and influence analyses were performed.

**Results:** Forty-four studies were included: 33 case series, 10 cohort studies, and one randomized trial. Kyphoplasty-specific evidence comprised one dedicated series and one comparative cohort. Any cement leakage occurred in 10.5% of patients (14 studies; 95% CI 5.7-18.4%), while trim-and-fill gave an exploratory adjusted estimate of 20.4%. Early pain reduction averaged 5.13 points on a 0-10 scale (8 studies; 95% CI 4.48-5.77; I2=89.4%). Complete or near-complete pain relief occurred in 79.4% of patients (10 studies), and recurrence, progression, or retreatment occurred in 3.9% (13 studies). Symptomatic cement leakage was uncommon at 0.4%.

**Conclusion:** The available literature, which is mainly retrospective and vertebroplasty-based, supports substantial pain relief with infrequent symptomatic complications. Kyphoplasty data remain insufficient for a reliable technique comparison. Prospective studies with standardized clinical and imaging outcomes are needed.

## 1. Introduction

Vertebral hemangiomas are common benign vascular lesions of the spine and are often found incidentally on imaging. Prevalence estimates reach 10-12% in the general population [1]. Most lesions remain asymptomatic and need no treatment, while about 1-3% become symptomatic or show aggressive behavior [2]. Clinical presentation ranges from persistent axial pain to vertebral expansion, cortical destruction, pathological fracture, and epidural extension. In advanced cases, spinal cord or nerve root compression may lead to neurological deficits [3–5].

Treatment is selected according to pain, lesion morphology, mechanical stability, epidural disease, and neurological status. Observation is usual for asymptomatic lesions. Symptomatic or aggressive lesions may require radiotherapy, embolization, percutaneous augmentation, surgical decompression, or a combination of these approaches [3,6–8]. Percutaneous vertebroplasty injects polymethyl methacrylate directly into the vertebral body to provide stabilization and pain control [1,9]. Balloon kyphoplasty creates a cavity before cement injection and may allow lower-pressure delivery and partial restoration of vertebral height [10–13].

Most clinical evidence for vertebral augmentation in this condition concerns vertebroplasty [1,14]. Reports of kyphoplasty are less frequent and usually involve small series or mixed treatment cohorts. The studies also differ in the definition of aggressive disease, postoperative imaging, pain assessment, adjunctive treatment, and follow-up [3,6,15–17]. These differences complicate interpretation of cement leakage and recurrence. A technique may appear safer when leakage is assessed only by fluoroscopy, because routine computed tomography detects small asymptomatic leaks more often. Direct comparisons between vertebroplasty and kyphoplasty are especially scarce.

A focused synthesis is therefore needed to define what can be concluded from the available data and where uncertainty remains because of study design and sparse reporting. This systematic review and meta-analysis evaluated clinical outcomes after vertebroplasty- or kyphoplasty-based treatment of symptomatic vertebral hemangiomas. The main outcomes were early pain reduction, complete or near-complete pain relief, any cement leakage, symptomatic cement leakage, and recurrence, progression, or retreatment. We also examined treatment context, leakage ascertainment, sensitivity analyses, and influence diagnostics. A direct comparison of vertebroplasty and kyphoplasty was planned, but its feasibility depended on the amount of comparative evidence identified.

## 2. Methods

### 2.1 Registration and reporting

The review was conducted according to the PRISMA 2020 statement [18]. The protocol was registered prospectively in PROSPERO (CRD420261439539) [19]. The completed PRISMA checklist is provided as Supplementary File 1. The protocol planned a head-to-head comparison of vertebroplasty and kyphoplasty. After study selection, the comparative evidence was found to be too sparse for that analysis. The resulting amendment to single-arm synthesis was made before pooled estimates were calculated and is described below.

### 2.2 Information sources and search strategy

PubMed, Embase, Web of Science Core Collection, Scopus, and the Cochrane Library were searched from inception to January 2, 2026. An updated search was completed on July 5, 2026. No date or language restriction was applied. Non-English reports were translated for eligibility assessment and data extraction. The search combined terms for vertebral or spinal hemangioma with terms for vertebroplasty, kyphoplasty, vertebral augmentation, cement augmentation, and polymethyl methacrylate. Complete database-specific strategies are provided in Supplementary File 2.

As a representative example, the PubMed search syntax was constructed as follows:

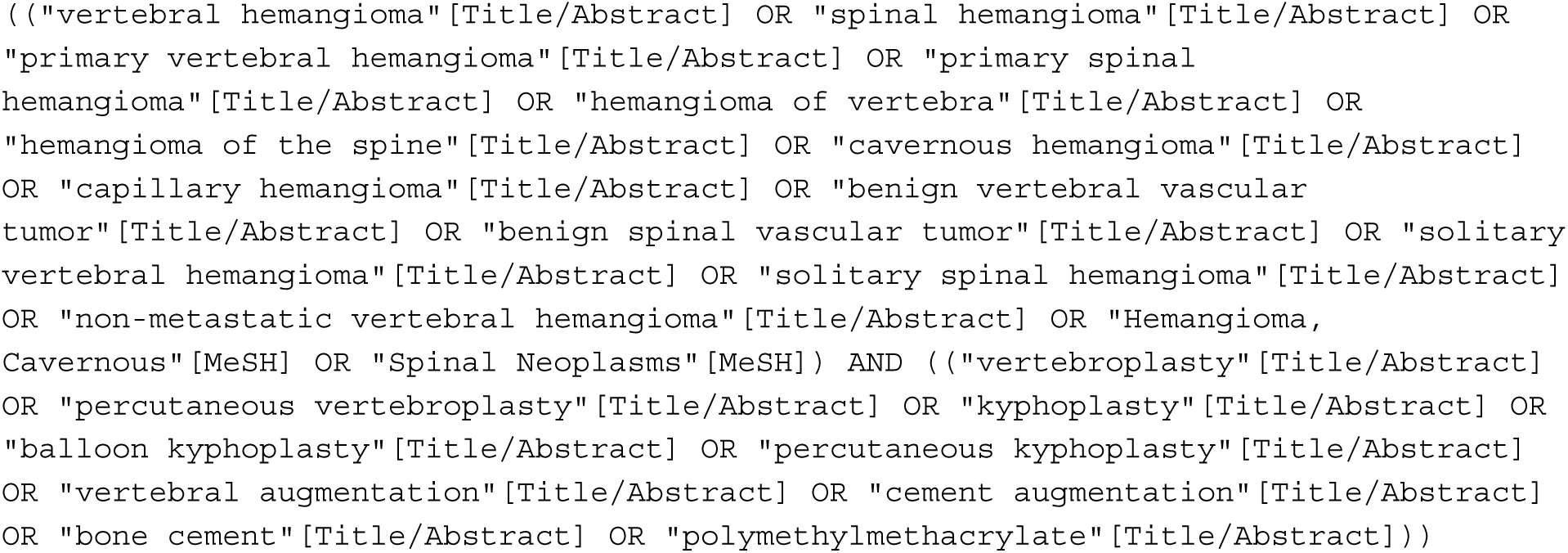

### 2.3 Eligibility criteria

Studies were eligible when they included patients with symptomatic or aggressive vertebral hemangioma and reported outcomes after percutaneous vertebroplasty or balloon kyphoplasty, used alone or with decompression, embolization, radiofrequency ablation, intralesional ethanol, or another adjunctive procedure. Randomized trials, prospective or retrospective cohorts, and case series with at least five patients were eligible. Studies with a smaller hemangioma subgroup within a broader augmentation cohort were retained only when relevant outcome data could be extracted.

The primary outcomes were pain relief measured on a Visual Analogue Scale and cement leakage, including any radiographic leakage and symptomatic leakage. Secondary outcomes included recurrence, progression, retreatment, reoperation, disability, neurological recovery, and other reported adverse events. Case reports, reviews, editorials, conference abstracts without a full report, technical notes without an eligible cohort, animal studies, duplicate reports, studies of other pathologies, and reports without extractable outcomes were excluded.

### 2.4 Study selection

Records were managed in EndNote 21 and duplicates were removed before screening. F.E., M.K., and R.Y. screened titles and abstracts using a standardized form. F.M.M. and A.H.N. assessed full texts. A calibration sample of 20 full-text reports was reviewed by both assessors, with 85% agreement, as recorded in the final PRISMA flow diagram. Remaining reports were divided between the assessors after calibration. Disagreements were resolved by discussion, with senior review by F.F. when required. The list of included and excluded full-text reports is provided as Supplementary Files 3 and 6.

### 2.5 Data extraction

F.F. and F.M.M. developed the extraction form and variable definitions. A.K. and A.H. extracted study and patient characteristics, operative details, pain and functional outcomes, cement leakage, recurrence, retreatment, neurological outcomes, follow-up, and adverse events. F.F. and F.M.M. cross-checked the completed dataset against the source reports. Disagreements were resolved by re-examination of the original article. The final extraction workbook is provided as Supplementary File 4.

When several postoperative assessments were reported, the longest follow-up with extractable data was used for descriptive outcomes. For early pain change, the earliest postoperative assessment within approximately six weeks was used. Missing standard deviations were derived from reported confidence intervals, standard errors, p values, ranges, or interquartile ranges when possible, using established methods [20,21]. A study was excluded from the quantitative synthesis of an outcome when the required variance or denominator could not be obtained.

### 2.6 Risk of bias

A.K. and F.M.M. independently used the Joanna Briggs Institute critical appraisal checklist appropriate to each study design [22]. Studies with randomized allocation were assessed as randomized trials. Non-randomized studies with two or more treatment groups were assessed as cohort studies, and single-arm studies were assessed as case series. Disagreements were resolved by consensus, with adjudication by F.F. The completed study-level checklists are provided as Supplementary File 5.

### 2.7 Statistical analysis

Only one small cohort provided extractable outcomes for vertebroplasty and kyphoplasty within the same study population [36]. A pairwise random-effects comparison was therefore not feasible for most outcomes. Each outcome was pooled across evaluable studies as a single-arm estimate. Treatment technique and other clinical sources of heterogeneity were examined only when the available data allowed a meaningful subgroup analysis.

Analyses were performed in R version 4.5.1 using the metafor, meta, and dmetar packages [23]. Any cement leakage, symptomatic cement leakage, complete or near-complete pain relief, and recurrence, progression, or retreatment were pooled with binomial-normal random-effects models. Exact Clopper-Pearson confidence intervals were calculated at study level, and pooled estimates were back-transformed from the logit scale. Early change in VAS score was analyzed with a restricted maximum-likelihood random-effects model and Hartung-Knapp-Sidik-Jonkman adjustment [24]. A within-patient correlation of 0.50 was used in the primary analysis, with 0.25 and 0.75 assessed in sensitivity analyses. A 95% prediction interval was calculated for early pain change.

Heterogeneity was quantified with I2. Any cement leakage was explored by routine postoperative CT versus no or unclear routine CT. Other outcomes were explored by stand-alone augmentation versus adjunctive or combined treatment. Sensitivity analyses varied denominators, risk-of-bias restrictions, treatment context, response definitions, and correlation assumptions as relevant. Small-study effects were examined with funnel plots, Egger regression [25], and trim-and-fill [26] when sufficient studies were available. Leave-one-out analysis, Baujat plots, externally studentized residuals, DFFITS, Cook’s distance, and hat values were used to identify influential reports. These analyses were treated cautiously for sparse-event proportions. Two-sided p<0.05 was considered statistically significant.

## 3. Results

### 3.1 Study selection

The searches identified 6679 records: 2527 from Scopus, 857 from PubMed, 98 from Web of Science, 1157 from Embase, and 2040 from the Cochrane Library. After removal of 4747 duplicates, 1,932 records underwent title and abstract screening and 1,843 were excluded. All 89 reports sought for full-text review were retrieved. Forty-five reports were excluded because of an ineligible population (n=18), intervention or comparison (n=5), study design (n=10), or design-related concerns and lack of relevant outcome data (n=12). Forty-four studies were included: 33 case series, 10 cohort studies, and one randomized trial (Figure 1).

**Figure 1.**
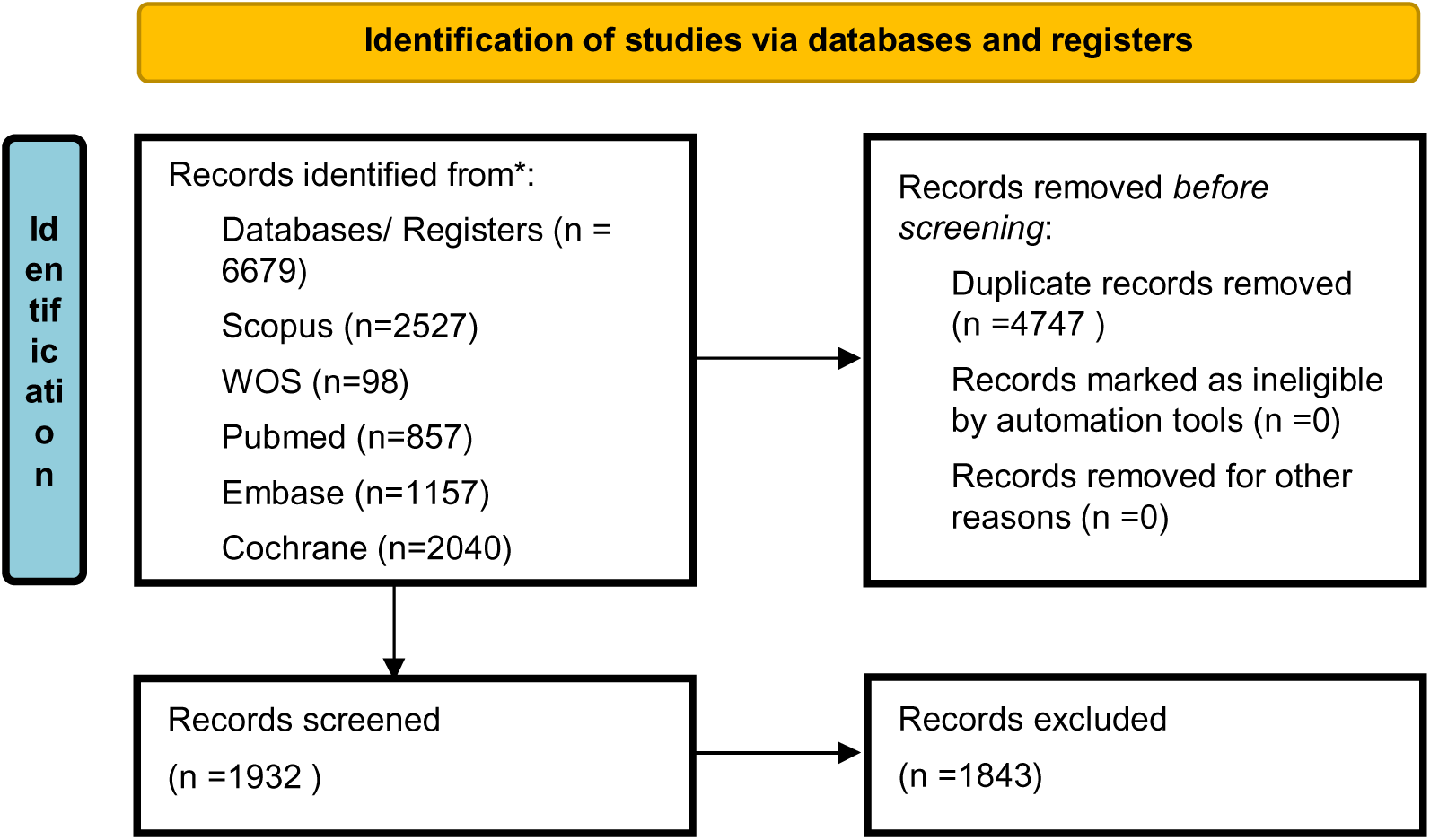
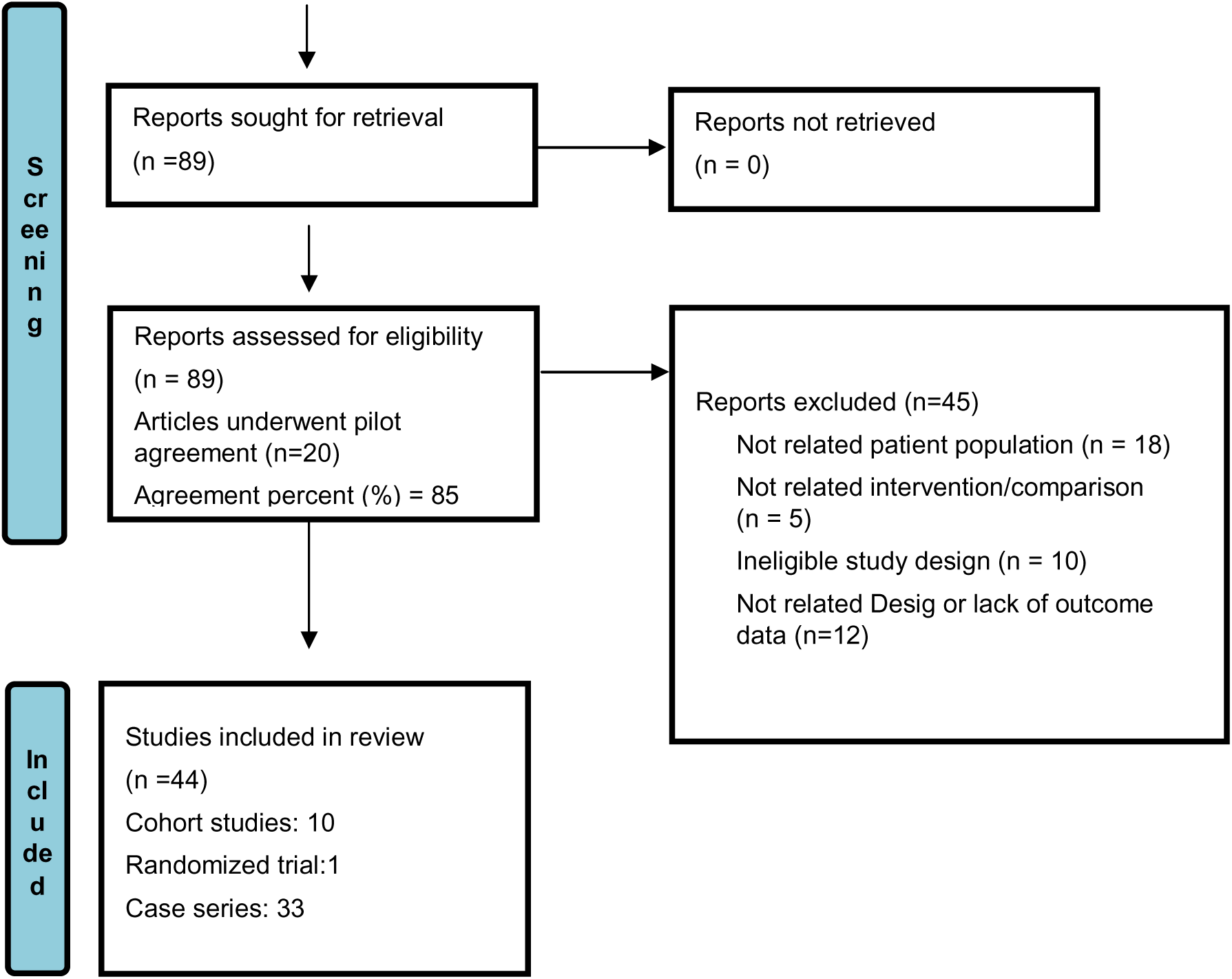
PRISMA 2020 flow diagram of study identification, screening, eligibility assessment, and inclusion.

### 3.2 Study and disease characteristics

The 44 studies were conducted across Europe, Asia, the Middle East, and North America. Most were retrospective, single-center reports, although the evidence also included prospective case series, comparative cohorts, one multicenter series, and one randomized trial. Eligible hemangioma samples ranged from very small subgroups within broad vertebral augmentation cohorts to dedicated series of more than 100 patients. This variation was important because some large procedural safety reports contained only a few patients with hemangioma, while several smaller studies were designed specifically around this condition.

Symptomatic axial pain was the main indication in non-aggressive lesions. Studies of aggressive disease more often included cortical destruction, vertebral expansion, pathological fracture, epidural extension, or neurological deficit. Thoracic and lumbar lesions predominated, whereas cervical and sacral lesions were reported less often. Age, sex, baseline disability, and the proportion of aggressive lesions were inconsistently reported. Neurological deficits were concentrated in cohorts of aggressive thoracic disease and were usually treated within a combined surgical or endovascular strategy. Detailed study characteristics and the extractable hemangioma subgroup from each report are provided in Table 1 and Supplementary File 4.

**Table 1.**
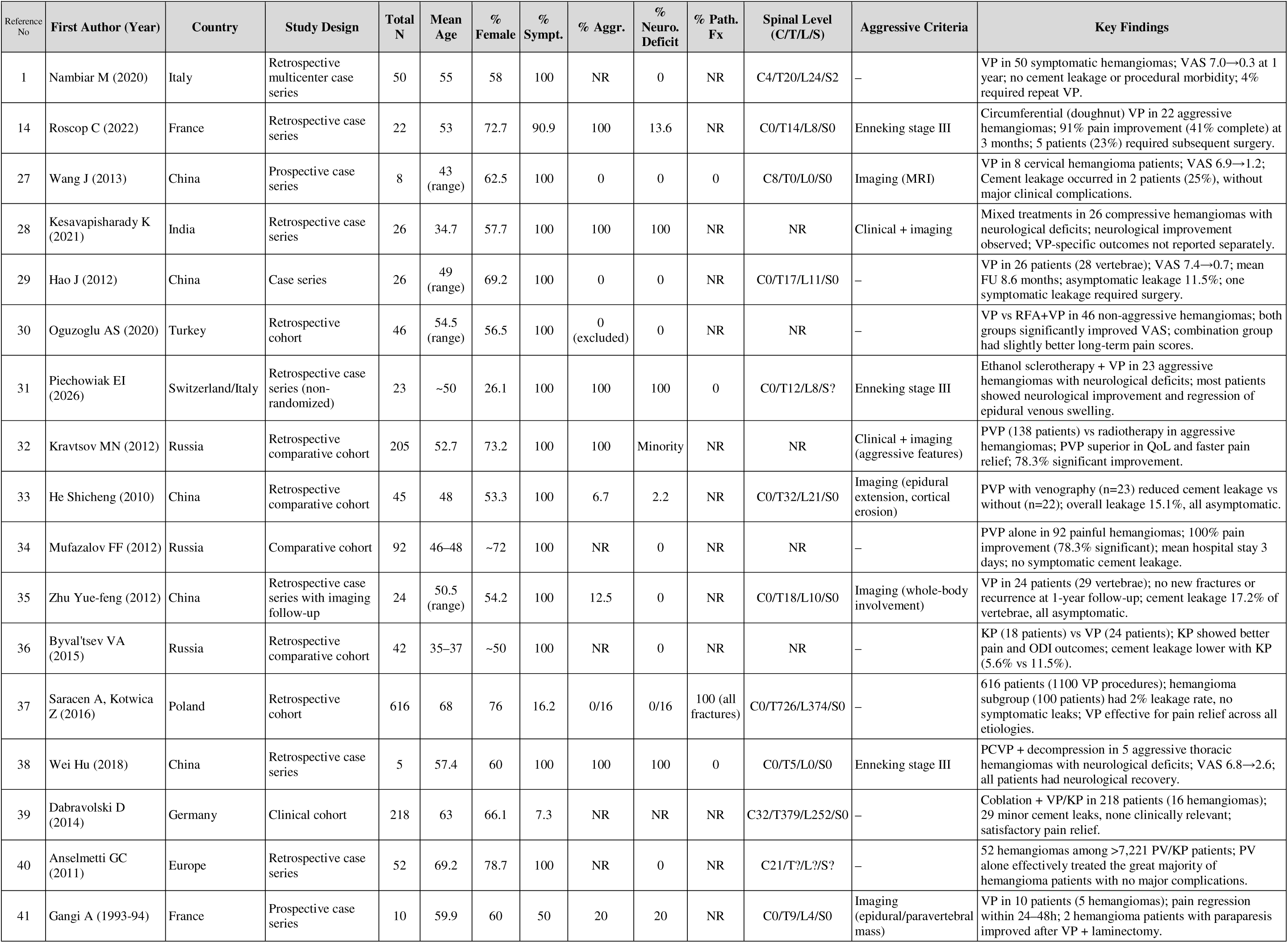
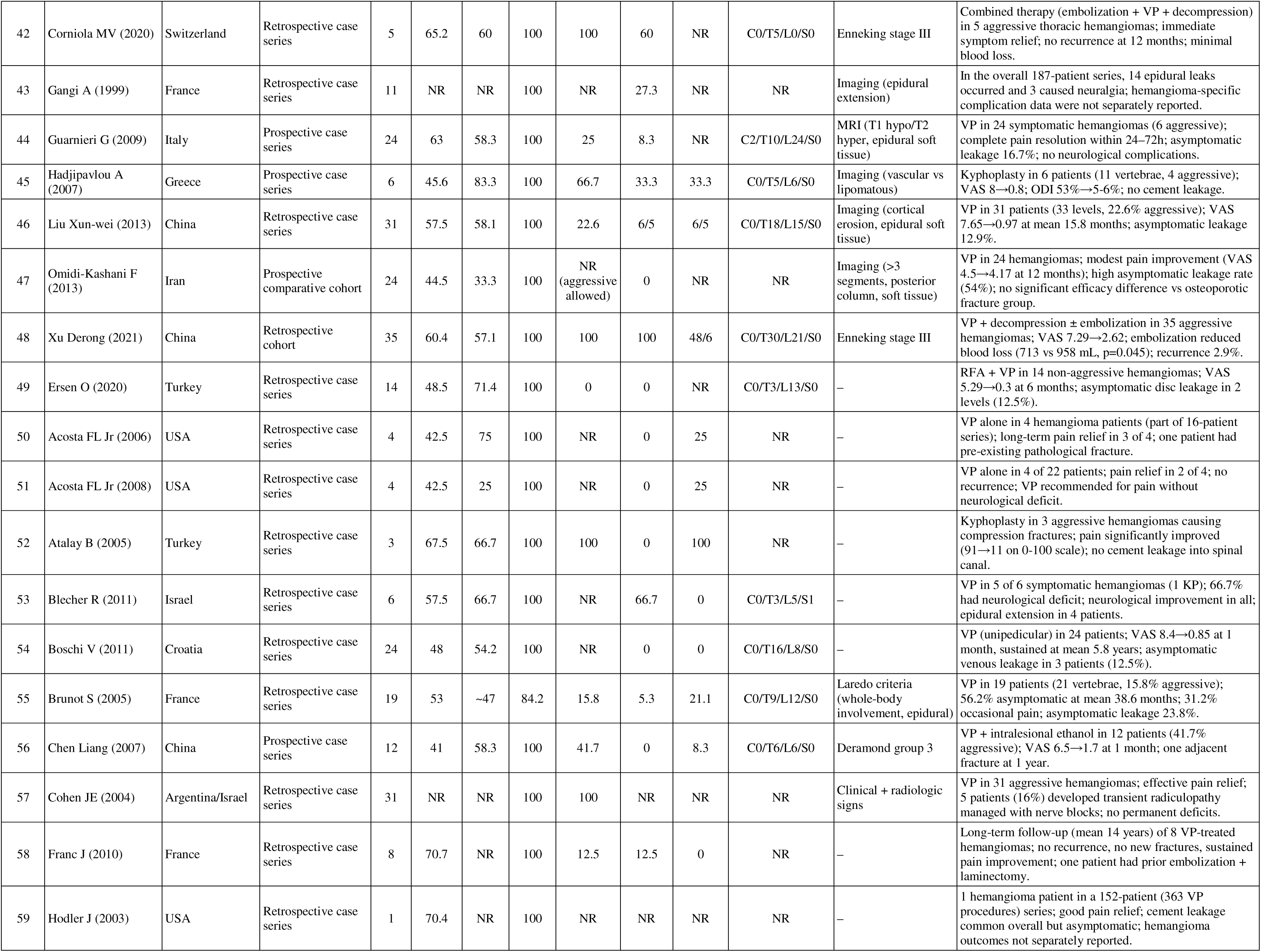
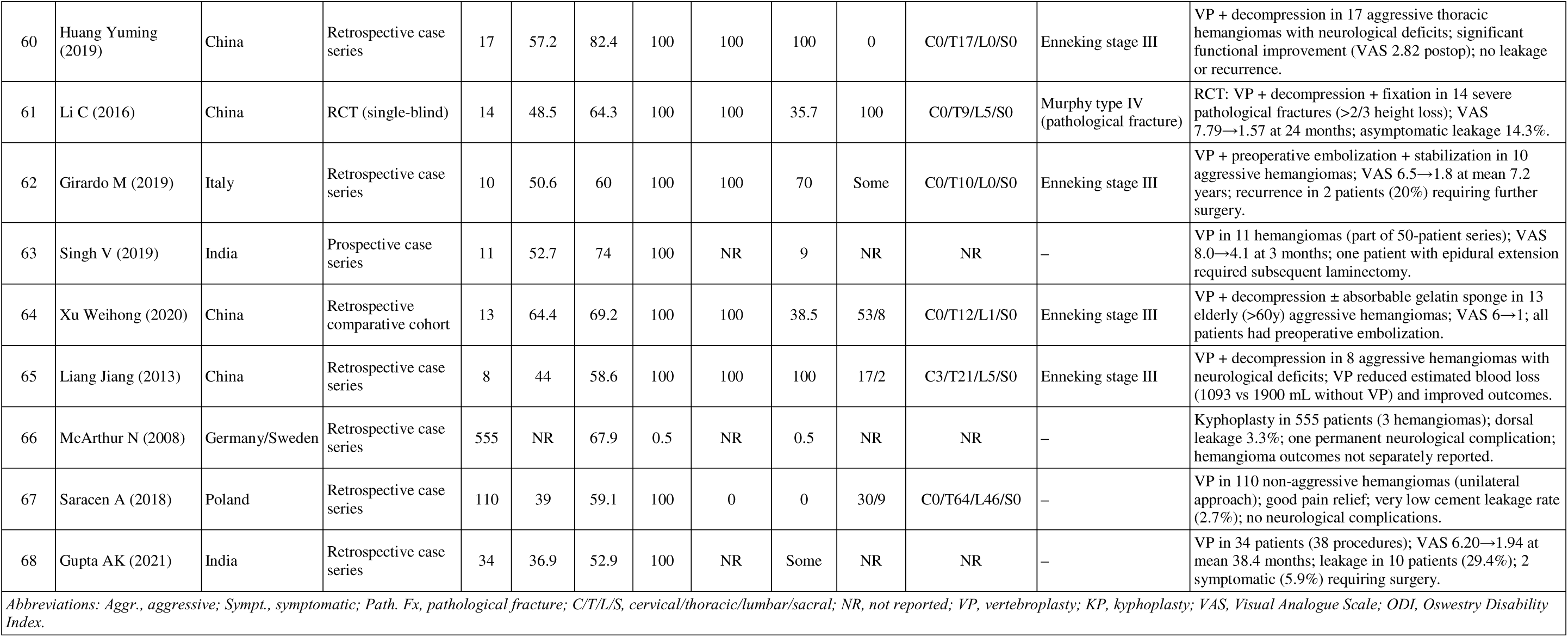
Characteristics of the 44 included studies.

### 3.3 Procedural characteristics

Polymethyl methacrylate was the cement reported throughout the included augmentation literature. Vertebroplasty was usually performed through a transpedicular approach, with unilateral or bilateral access selected according to lesion location and the intended cement distribution. Some reports used venography before injection, while others relied on fluoroscopic monitoring alone. Reported cement volumes and injection endpoints varied with vertebral level, lesion size, cortical integrity, and treatment strategy. Several articles did not provide enough technical detail to compare cement viscosity, injection pressure, or the completeness of lesion filling.

Stand-alone vertebroplasty was the most frequent procedure for painful lesions without major neural compression. Radiofrequency ablation, intralesional ethanol, or sclerotherapy was combined with augmentation in selected non-aggressive or locally aggressive lesions [30,31,49,56]. When epidural disease or neurological deficit was present, augmentation was commonly integrated with preoperative embolization, decompression, and, when required, fixation [14,38,48,60–65]. The purpose of cement in these multimodal procedures was generally to stabilize the vertebral body and reduce intraoperative bleeding rather than to treat the epidural component directly.

Kyphoplasty-specific evidence was limited to one dedicated series of six patients [45], one direct comparative cohort of 42 patients [36], and very small hemangioma subgroups within broader reports [39,52,53,66]. The comparative cohort reported greater improvement in pain and disability and a lower observed leakage rate with kyphoplasty, but treatment allocation was not randomized and the sample was small. These data were insufficient for an outcome-level pooled comparison between kyphoplasty and vertebroplasty.

### 3.4 Clinical outcomes and complications

Most vertebroplasty series reported a rapid reduction in pain, sometimes within 24-72 hours, followed by sustained improvement during later follow-up [41,44]. Continuous Visual Analogue Scale data were available in only part of the literature. Other reports used complete relief, partial relief, analgesic use, or return to activity as the clinical endpoint. Complete pain relief was common, although response definitions and assessment times differed. Long-term series described maintained benefit over several years, but loss to follow-up and the absence of a control group limited interpretation [54,58].

Neurological recovery was mainly described in multimodal series that combined augmentation with decompression, embolization, or fixation. Most of these reports described improvement after treatment, but neurological scales and follow-up intervals were not consistent [38,48,60–65]. Functional outcomes such as the Oswestry Disability Index were reported much less often than pain scores. Imaging follow-up was also variable. Some studies described regression of epidural vascular components, whereas intraosseous tumor response was rarely measured with a standardized volumetric method.

Radiographic cement leakage ranged from 0% to 54% across individual reports [1,47]. Many leaks were detected only on imaging and had no clinical consequence. The reported frequency depended partly on whether postoperative CT was routine and on how venous, discal, paravertebral, or epidural leakage was defined. Symptomatic leakage requiring treatment was uncommon. Reported events included nerve root compression, spinal canal leakage, and a small number of decompressive procedures [29,37,43,68]. Pulmonary cement emboli were usually asymptomatic when they were reported.

Recurrence, progression, or retreatment was infrequent in most series. Repeat augmentation was used for persistent pain or incomplete filling in some patients, while later decompression or stabilization was required when aggressive disease progressed or neurological compression remained [1,14,45,48,62]. Follow-up ranged from several months to more than a decade, and many studies did not specify a uniform imaging schedule. The longer series generally reported durable pain control, but the small number of patients at extended follow-up limited precise estimation of late recurrence [54,58].

### 3.5 Risk of bias

Risk-of-bias findings are summarized in Tables 2-4. Case series often reported clear diagnostic criteria, intervention details, and clinical outcomes. Consecutive or complete recruitment was frequently unclear, and several reports did not explain whether all eligible patients were included. Follow-up was sometimes described only as a mean or range, without the number of patients assessed at each time point. These limitations were particularly relevant for recurrence and late complications.

**Table 2.**
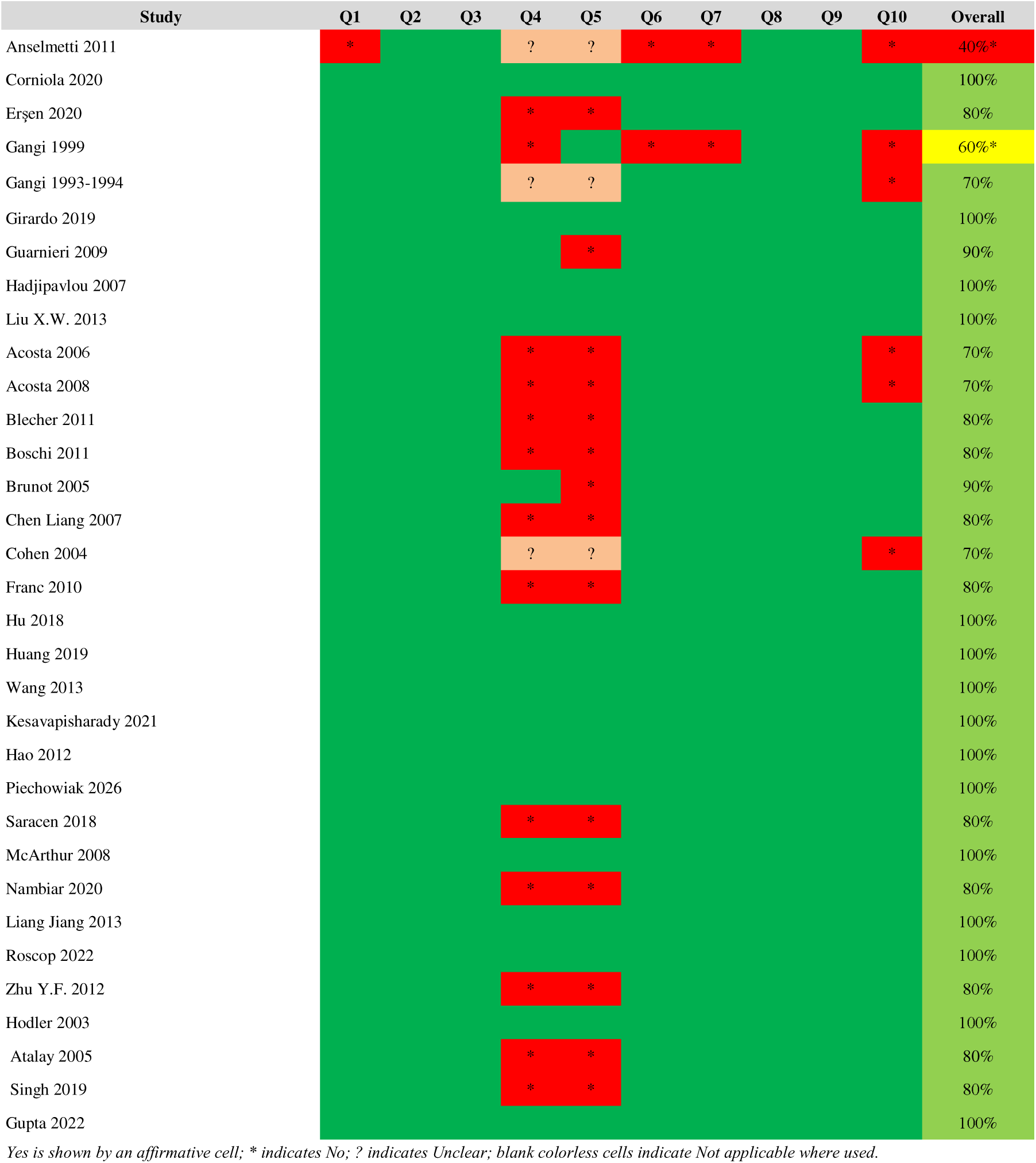
JBI risk-of-bias assessment for case series.

| Study | Q1 | Q2 | Q3 | Q4 | Q5 | Q6 | Q7 | Q8 | Q9 | Q10 | Overall |
| --- | --- | --- | --- | --- | --- | --- | --- | --- | --- | --- | --- |
| Anselmetti 2011 | * |  |  | ? | ? | * | * |  |  | * | 40%* |
| Corniola 2020 |  |  |  |  |  |  |  |  |  |  | 100% |
| Erşen 2020 |  |  |  | * | * |  |  |  |  |  | 80% |
| Gangi 1999 |  |  |  | * |  | * | * |  |  | * | 60%* |
| Gangi 1993-1994 |  |  |  | ? | ? |  |  |  |  | * | 70% |
| Girardo 2019 |  |  |  |  |  |  |  |  |  |  | 100% |
| Guarnieri 2009 |  |  |  |  | * |  |  |  |  |  | 90% |
| Hadjipavlou 2007 |  |  |  |  |  |  |  |  |  |  | 100% |
| Liu X.W. 2013 |  |  |  |  |  |  |  |  |  |  | 100% |
| Acosta 2006 |  |  |  | * | * |  |  |  |  | * | 70% |
| Acosta 2008 |  |  |  | * | * |  |  |  |  | * | 70% |
| Blecher 2011 |  |  |  | * | * |  |  |  |  |  | 80% |
| Boschi 2011 |  |  |  | * | * |  |  |  |  |  | 80% |
| Brunot 2005 |  |  |  |  | * |  |  |  |  |  | 90% |
| Chen Liang 2007 |  |  |  | * | * |  |  |  |  |  | 80% |
| Cohen 2004 |  |  |  | ? | ? |  |  |  |  | * | 70% |
| Franc 2010 |  |  |  | * | * |  |  |  |  |  | 80% |
| Hu 2018 |  |  |  |  |  |  |  |  |  |  | 100% |
| Huang 2019 |  |  |  |  |  |  |  |  |  |  | 100% |
| Wang 2013 |  |  |  |  |  |  |  |  |  |  | 100% |
| Kesavapisharady 2021 |  |  |  |  |  |  |  |  |  |  | 100% |
| Hao 2012 |  |  |  |  |  |  |  |  |  |  | 100% |
| Piechowiak 2026 |  |  |  |  |  |  |  |  |  |  | 100% |
| Saracen 2018 |  |  |  | * | * |  |  |  |  |  | 80% |
| McArthur 2008 |  |  |  |  |  |  |  |  |  |  | 100% |
| Nambiar 2020 |  |  |  | * | * |  |  |  |  |  | 80% |
| Liang Jiang 2013 |  |  |  |  |  |  |  |  |  |  | 100% |
| Roscop 2022 |  |  |  |  |  |  |  |  |  |  | 100% |
| Zhu Y.F. 2012 |  |  |  | * | * |  |  |  |  |  | 80% |
| Hodler 2003 |  |  |  |  |  |  |  |  |  |  | 100% |
| Atalay 2005 |  |  |  | * | * |  |  |  |  |  | 80% |
| Singh 2019 |  |  |  | * | * |  |  |  |  |  | 80% |
| Gupta 2022 |  |  |  |  |  |  |  |  |  |  | 100% |

Yes is shown by an affirmative cell; \* indicates No; ? indicates Unclear; blank colorless cells indicate Not applicable where used.

**Table 3.**
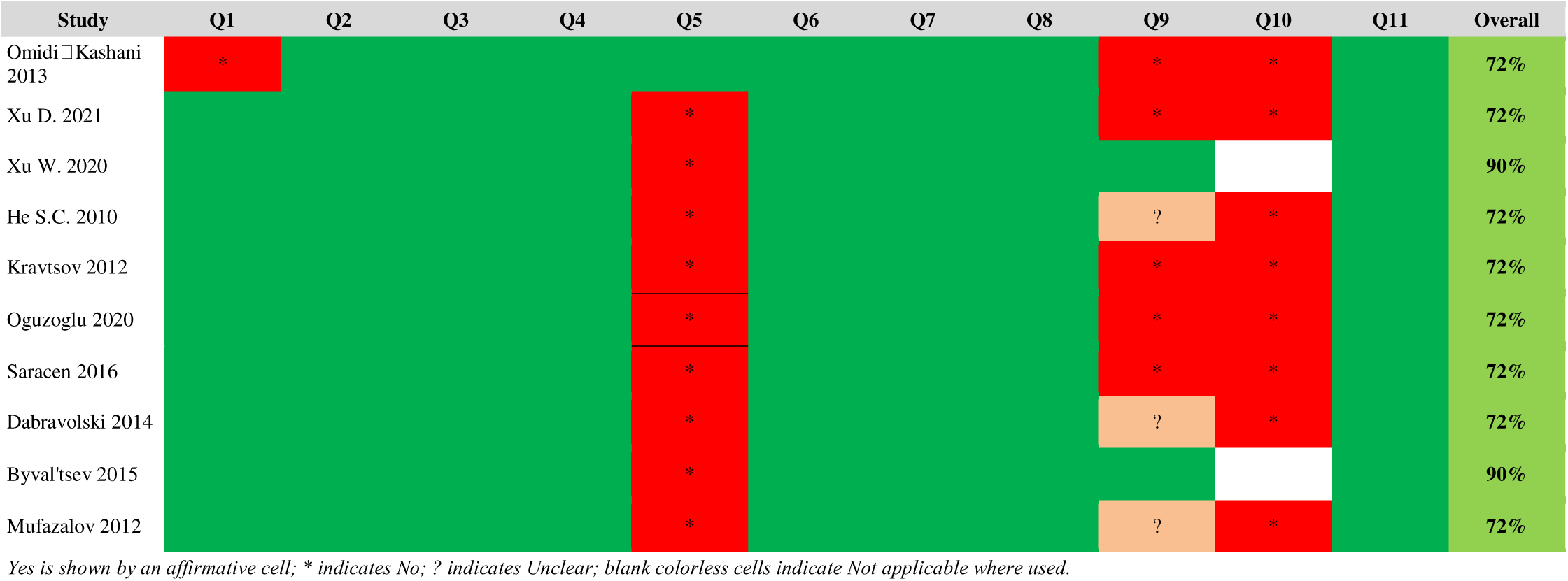
JBI risk-of-bias assessment for cohort studies.

**Table 4.** JBI risk-of-bias assessment for the randomized controlled trial.

| Study | Q1 | Q2 | Q3 | Q4 | Q5 | Q6 | Q7 | Q8 | Q9 | Q10 | Q11 | Q12 | Q13 | Overall |
| --- | --- | --- | --- | --- | --- | --- | --- | --- | --- | --- | --- | --- | --- | --- |
| Li C. 2016 |  | ? |  |  |  | * |  |  | ? |  |  |  |  | 75% |

Yes is shown by an affirmative cell; \* indicates No; ? indicates Unclear; blank colorless cells indicate Not applicable where used.

The cohort studies generally measured treatment exposure and outcomes in a similar manner across groups, but control of confounding was limited. Treatment choice was commonly influenced by lesion severity, epidural extension, fracture, neurological status, or surgeon preference. Follow-up strategies and losses were incompletely reported in several cohorts. The single randomized trial had unclear allocation concealment, no reported blinding of outcome assessors, and no explicit intention-to-treat analysis. These design features restrict causal comparison between procedures, even when individual reports met several checklist criteria.

### 3.6 Meta-analysis

A direct vertebroplasty-versus-kyphoplasty meta-analysis could not be performed because only one small comparative cohort reported extractable group-level outcomes [36]. The pooled results therefore describe vertebral augmentation in a literature dominated by vertebroplasty and should not be interpreted as comparative estimates.

Eight studies with 187 patients contributed to early VAS change (Figure 2). The pooled reduction was 5.13 points on a 0-10 scale (95% CI 4.48-5.77; I2=89.4%; 95% prediction interval 3.32-6.93). Stand-alone and adjunctive treatment subgroups did not differ significantly (5.26 versus 4.96 points; p=0.629). Sensitivity estimates ranged from 5.10 to 5.35 points. Egger regression was not significant (p=0.768). Xu et al. [48] and Liu et al. [46] contributed most to heterogeneity, while Oguzoglu et al. [30] was flagged by the externally studentized residual.

**Figure 2.**
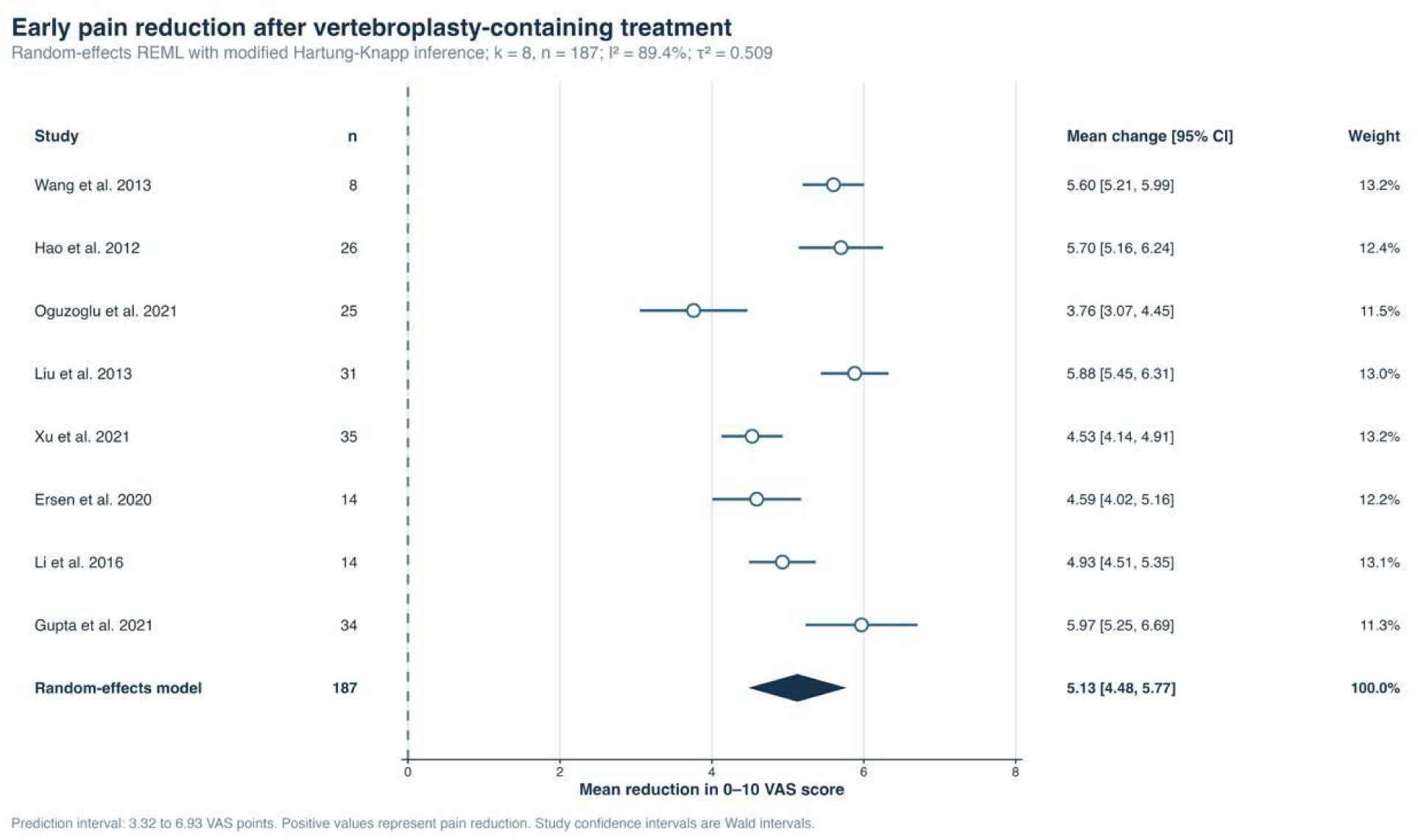
Pooled early mean reduction in Visual Analogue Scale score after vertebroplasty-containing treatment (REML random-effects model with Hartung-Knapp adjustment; k=8, n=187).

Ten studies with 335 patients reported complete or near-complete pain relief (Figure 3). The pooled proportion was 79.4% (95% CI 65.1-88.8%; I2=41.6%). The stand-alone and other or mixed treatment subgroups did not differ (81.6% versus 75.0%; p=0.984). Sensitivity estimates ranged from 72.8% to 83.7%, and leave-one-out estimates ranged from 75.1% to 81.7%. Egger regression was not significant (p=0.407), and trim-and-fill did not identify missing studies.

**Figure 3.**
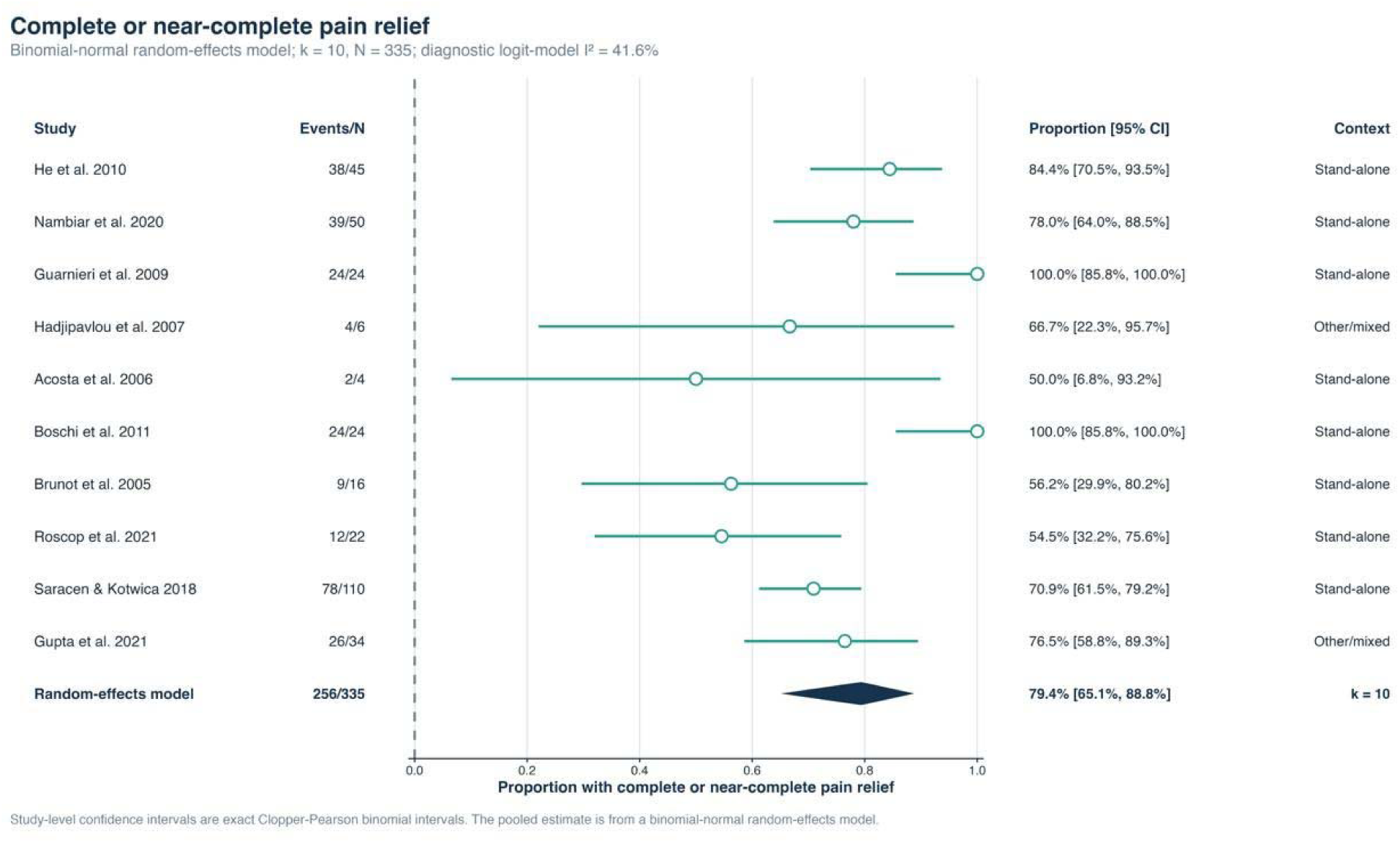
Pooled proportion of complete or near-complete pain relief (binomial-normal random-effects model; k=10, N=335).

Fourteen studies with 383 patients contributed to any cement leakage (Figure 4). The pooled proportion was 10.5% (95% CI 5.7-18.4%; I2=53.6%). Routine CT and no or unclear routine CT subgroups did not differ significantly (11.3% versus 9.1%; p=0.718). Sensitivity estimates ranged from 8.6% to 13.4%, and leave-one-out estimates ranged from 9.2% to 12.8%. Egger regression suggested asymmetry (p=0.043). Trim-and-fill imputed five studies and produced an exploratory adjusted estimate of 20.4% (95% CI 12.3-32.0%). Saracen et al. [67] was the most influential report.

**Figure 4.**
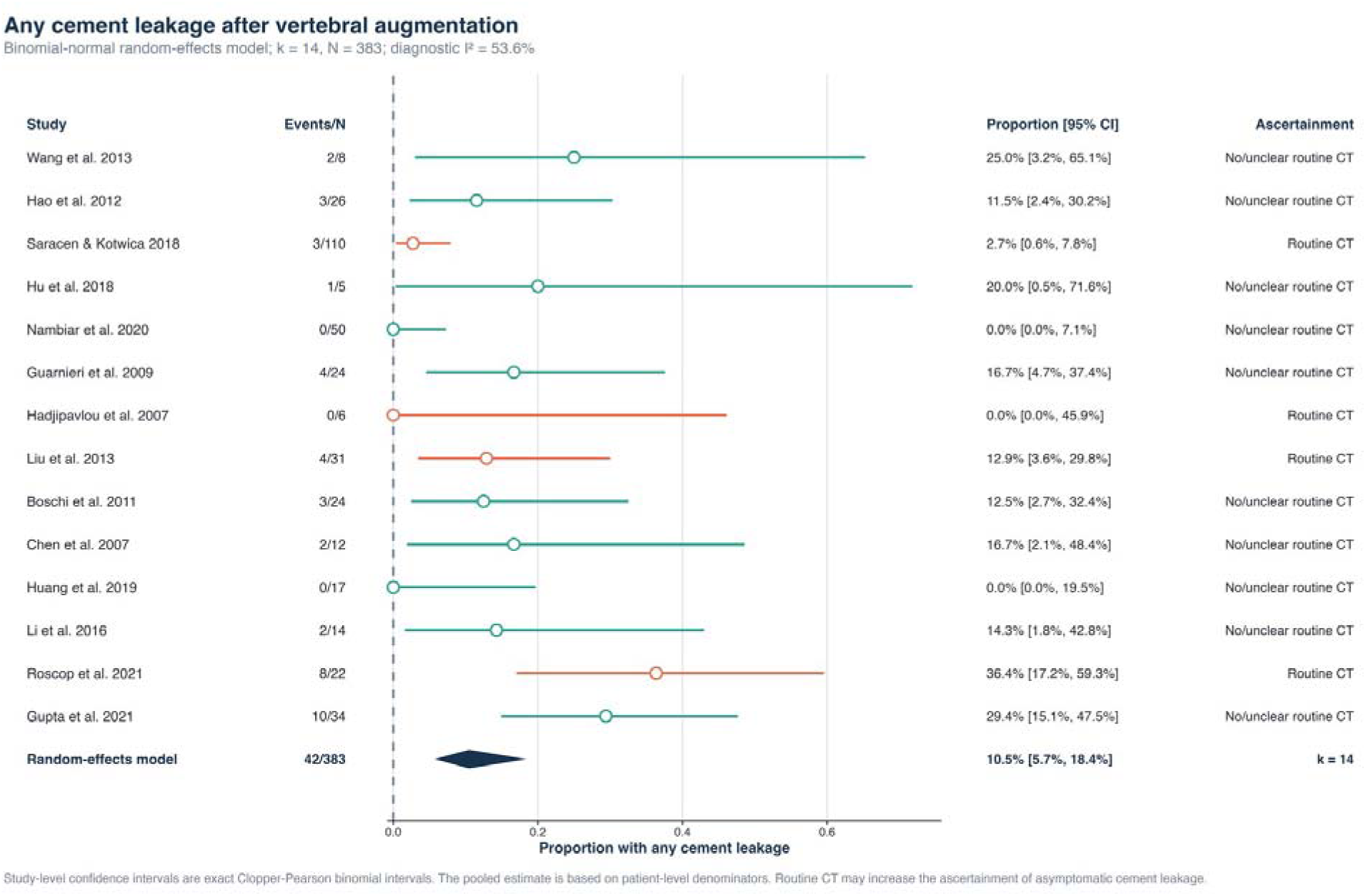
Pooled proportion of any cement leakage after vertebral augmentation (binomial-normal random-effects model; k=14, N=383).

Twenty studies with 546 patients and four events contributed to symptomatic cement leakage (Figure 5). The pooled proportion was 0.4% (95% CI 0.0-3.3%; I2=0%). Treatment-context subgroups did not differ (0.4% versus 0.9%; p=0.300). The modified Egger test was nominally significant (p=0.049), and trim-and-fill gave an exploratory adjusted estimate of 4.06% (95% CI 2.47-6.62%). These diagnostics are unstable with so few events. Gupta et al. [68] was the most influential report.

**Figure 5.**
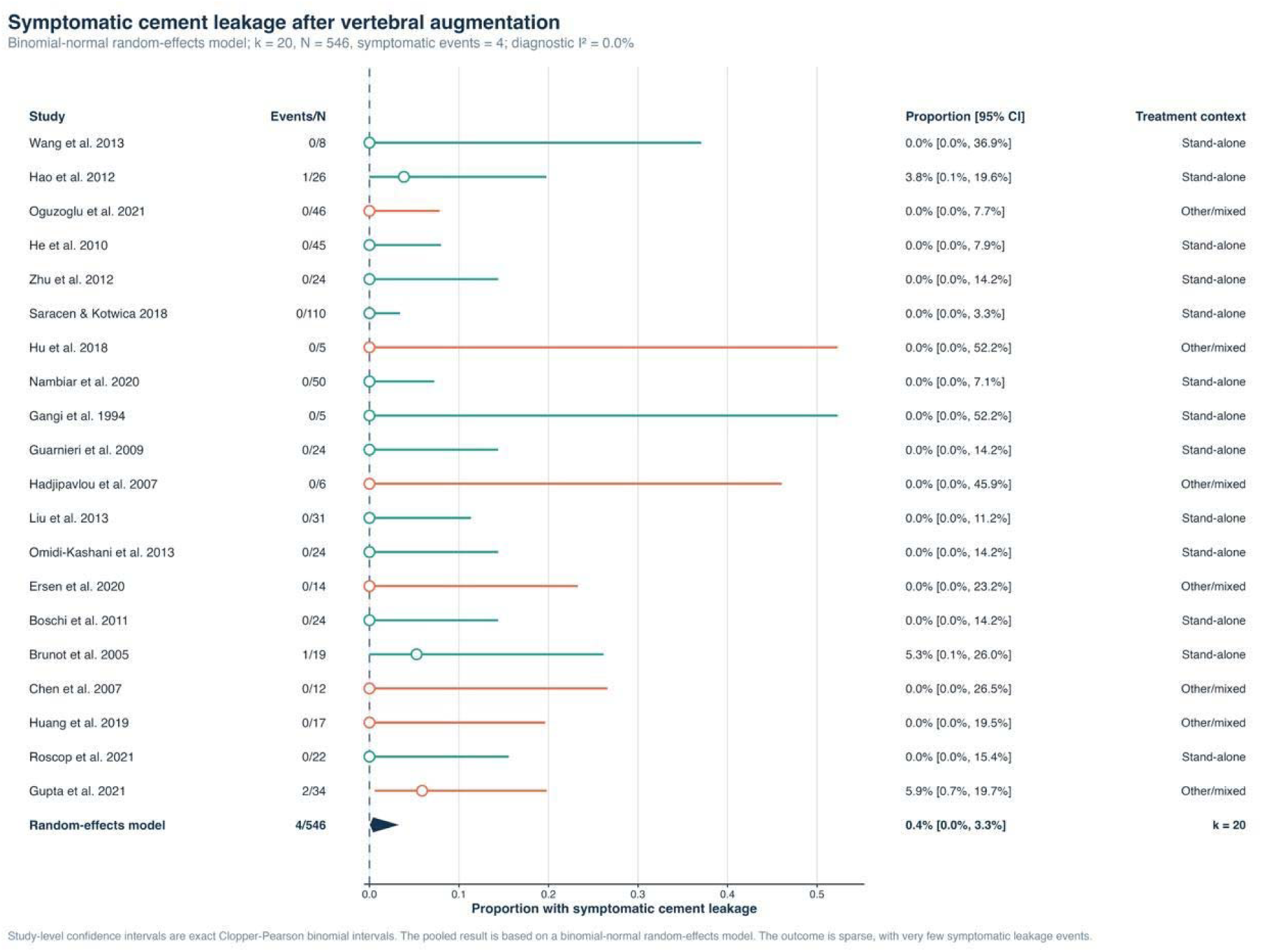
Pooled proportion of symptomatic cement leakage (binomial-normal random-effects model; k=20, N=546).

Thirteen studies with 258 patients and ten events contributed to recurrence, progression, or retreatment (Figure 6). The pooled proportion was 3.9% (95% CI 2.1-7.1%; I2=0%). Treatment-context subgroups did not differ (3.5% versus 4.2%; p=0.501), and leave-one-out estimates ranged from 3.2% to 4.3%. Egger regression was not significant (p=0.177). Trim-and-fill imputed three studies and gave an exploratory adjusted estimate of 7.0% (95% CI 4.3-11.4%). Girardo et al. [62] was the most influential report.

**Figure 6.**
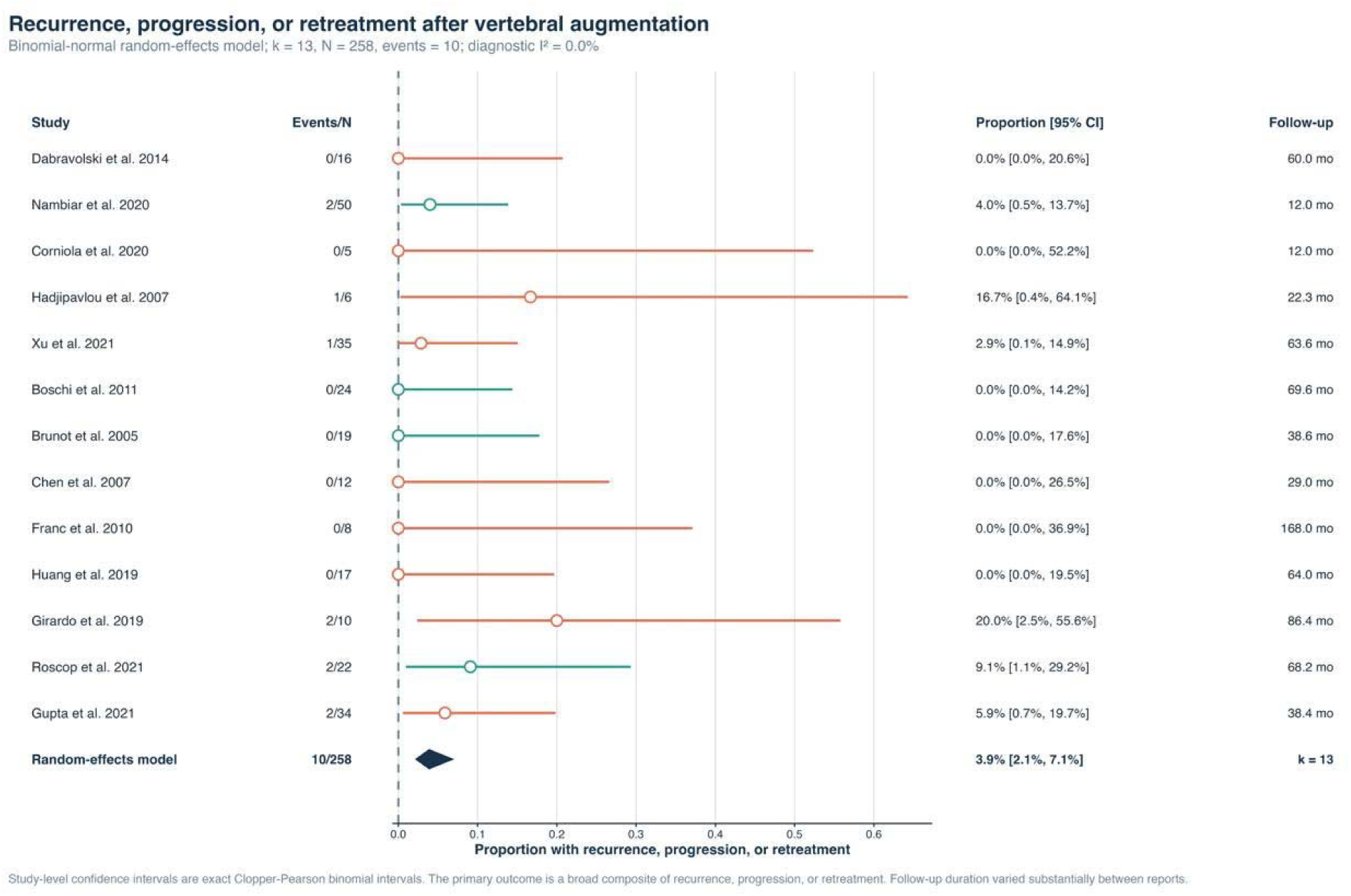
Pooled proportion of recurrence, progression, or retreatment (binomial-normal random-effects model; k=13, N=258).

## 4. Discussion

### 4.1 Main findings

This systematic review brought together 44 studies of vertebral augmentation for symptomatic or aggressive vertebral hemangioma. The evidence consistently indicated meaningful pain improvement after treatment. The pooled early VAS reduction was 5.13 points on a 0-10 scale, and about four of five patients achieved complete or near-complete pain relief. Radiographic cement leakage occurred more often than clinically symptomatic leakage. Recurrence, progression, or retreatment was uncommon in the pooled analysis. These findings give a quantitative summary of outcomes that were previously dispersed across small case series and mixed procedural cohorts.

The two pain analyses describe related but different aspects of recovery. The early VAS analysis measured the average change in pain intensity among patients with continuous scores. Its 95% prediction interval remained clinically favorable, although the high I2 showed that the magnitude of benefit varied considerably among studies. The complete or near-complete relief analysis included a broader set of reports using categorical responses. Its lower heterogeneity may partly reflect the simpler endpoint, but definitions of complete relief and the timing of assessment were not uniform. Taken together, the analyses support a substantial analgesic effect while showing that the exact size and durability of benefit cannot be predicted precisely for an individual patient.

The separation of any radiographic leakage from symptomatic leakage was clinically important. The pooled proportion for any leakage was 10.5%, compared with 0.4% for symptomatic events. A small epidural, venous, discal, or paravertebral leak detected on postoperative imaging does not carry the same consequence as neural compression or a clinically relevant embolic event. Combining these outcomes would overstate procedural morbidity. At the same time, the very low number of symptomatic events produced wide confidence intervals, so the pooled estimate should not be interpreted as absence of serious risk.

The evidence base was dominated by vertebroplasty. Only one small cohort directly compared vertebroplasty with kyphoplasty, and few additional reports provided kyphoplasty-specific hemangioma outcomes. A comparative meta-analysis was therefore not feasible. The pooled estimates describe augmentation in general within a predominantly vertebroplasty-treated population. They do not show equivalence between techniques and cannot establish that one approach is safer or more effective.

### 4.2 Clinical interpretation

Pain relief after cement augmentation may result from mechanical stabilization, thermal effects during cement polymerization, and occlusion of vascular spaces within the hemangioma [9,10]. These mechanisms are compatible with the rapid improvement described in many series and with sustained pain control during longer follow-up [44,54,58]. The large pooled VAS change is also consistent with a clinically important response rather than a small statistical difference. However, most studies lacked an untreated or sham control group. Regression to the mean, concurrent analgesia, postoperative care, and additional procedures may have contributed to the observed improvement.

The high heterogeneity in early VAS change probably reflects several clinical and methodological differences. Study populations ranged from patients with pain-predominant, non-aggressive lesions to patients with pathological fracture, epidural extension, or neurological deficit. Some patients received augmentation alone, while others underwent ablation, embolization, decompression, or fixation. Baseline pain intensity, follow-up timing, cement volume, lesion level, and pain-scale reporting also varied. The subgroup comparison by treatment context did not identify a significant difference, but broad subgroup labels cannot fully account for this diversity.

Complete or near-complete pain relief was reported in 79.4% of patients. This result is readily understandable for clinical counseling, although it is less standardized than a continuous change score. Some authors required absence of pain, while others combined complete and marked improvement. Several studies assessed the outcome within weeks, whereas others reported the best response or the status at the last follow-up. The pooled proportion should therefore be presented as the frequency of a major reported response across the available literature, rather than as a uniform endpoint measured at one fixed time.

Adequate cement distribution may influence both symptom control and later retreatment. Persistent pain and recurrence were sometimes described after incomplete lesion filling, and repeat augmentation produced further improvement in selected patients [1,14,45,62]. Even so, none of the included studies formally evaluated filling completeness as an independent predictor. More extensive filling may improve structural stabilization, but it may also increase leakage exposure. The available data cannot define an optimal cement volume or distribution pattern for hemangioma.

Kyphoplasty creates a cavity before cement delivery and may permit lower-pressure injection. It can also be considered when vertebral collapse or height restoration is a procedural concern [10–13]. The single direct cohort reported favorable pain, disability, and leakage findings with kyphoplasty [36], and the dedicated kyphoplasty series reported marked improvement without leakage [45]. Both studies were small, and treatment selection was susceptible to lesion characteristics and operator preference. Evidence from osteoporotic compression fractures cannot be transferred directly because vertebral hemangiomas contain vascular channels and may have cortical defects or epidural components. The present review does not support a conclusion of procedural superiority.

The leakage analysis also requires attention to imaging practice. Routine postoperative CT can identify small cement deposits that are not visible on fluoroscopy or plain radiography. The subgroup analysis did not find a significant difference between routine CT and no or unclear routine CT, but only four studies were in the routine CT group and heterogeneity was substantial. Funnel asymmetry and trim-and-fill produced a higher exploratory estimate for any leakage. These analyses raise the possibility of under-reporting, although trim-and-fill is difficult to interpret for single proportions and heterogeneous observational studies.

For aggressive lesions with epidural extension or neurological compromise, augmentation cannot remove the extraosseous component or directly decompress neural tissue. Several included series combined embolization and decompression with augmentation to control bleeding and provide anterior column support [48,56,60–65]. Neurological improvement in these cohorts reflects the complete treatment strategy rather than cement injection alone. Vertebral augmentation in this setting should be viewed as one component of multimodal care, with the sequence of embolization, cement injection, decompression, and fixation adapted to lesion anatomy and mechanical stability.

The pooled recurrence, progression, or retreatment proportion of 3.9% is encouraging, but it is based on ten events in 258 patients. Follow-up varied greatly, and many reports did not use scheduled long-term MRI or CT. Late progression may therefore have been missed, especially when follow-up ended after pain improvement. The higher trim-and-fill estimate of 7.0% also indicates that the observed proportion may be conservative. Long-term series describing durable control remain informative [54,58], but they cannot replace standardized prospective surveillance.

### 4.3 Comparison with previous reviews

The findings are consistent with previous reviews that described favorable symptom control after vertebroplasty or kyphoplasty for symptomatic and aggressive vertebral hemangioma [1,3,6]. Earlier reviews were mainly narrative because the literature consisted of small, clinically diverse series. They emphasized the usefulness of augmentation for pain and the need for decompression when an epidural component caused neurological compromise. The present analysis supports this clinical distinction and provides outcome-specific pooled estimates for pain, leakage, and recurrence.

The review of surgical management by Piper et al. found that patients with neurological deficits often received combinations of decompression, embolization, radiotherapy, stabilization, and vertebral augmentation [69]. Our findings are compatible with that treatment framework. The pooled pain results primarily apply to augmentation-containing treatment, while neurological recovery in aggressive disease cannot be attributed to augmentation alone. This distinction is important because combining pain-predominant and neurologically compromised populations can create an impression that one procedure addresses all manifestations of the disease.

The observed leakage proportion is lower than some estimates reported for vertebral augmentation across mixed spinal disorders. General leakage risk-factor analyses include osteoporotic fractures and malignant lesions with different cortical and venous anatomy [70]. Their results provide useful procedural context, but the estimates cannot be applied directly to hemangioma. Within the present review, leakage varied from absent to more than half of treated patients, largely because imaging protocols, denominators, and definitions differed. Separating radiographic from symptomatic leakage provides a more clinically interpretable summary.

A further contribution of this review is the detailed examination of robustness and influence. Sensitivity analyses showed that the principal pain and proportion estimates were generally stable across alternative decisions. Influence diagnostics identified several reports that contributed strongly to individual models, particularly Saracen et al. for any leakage [67], Gupta et al. for symptomatic leakage [68], and Girardo et al. for recurrence [62]. Reporting these effects gives a clearer account of how much the pooled results depend on particular cohorts.

### 4.4 Clinical implications

For a patient with focal pain arising from a vertebral hemangioma and no major neurological compromise, vertebroplasty has the broadest condition-specific evidence. The expected clinical course is a marked reduction in pain, often early after treatment. Patient counseling should still acknowledge that the supporting literature is mainly retrospective and that a minority of patients have persistent pain or require another procedure. Alternative causes of axial pain should be considered before intervention, particularly when imaging features are not clearly concordant with symptoms.

Procedure planning should integrate lesion morphology with clinical presentation. Cortical disruption, epidural extension, venous drainage, vertebral collapse, and spinal level may affect access and leakage risk. Slow cement injection under continuous imaging, careful attention to posterior wall integrity, and cessation of injection when extraosseous spread is seen remain important even though pooled symptomatic leakage was uncommon. A low event proportion does not remove the possibility of neural injury or the need for immediate management when symptomatic leakage occurs.

Kyphoplasty may be selected when cavity creation, cement containment, or partial height restoration is considered useful. The choice should be based on anatomy and operator experience rather than an assumption of proven superiority. The current evidence cannot support a routine preference for kyphoplasty, and it does not provide a reliable comparative estimate for leakage or pain. Patients should be informed when a treatment decision relies on extrapolation from other vertebral disorders.

Aggressive disease with spinal canal extension, instability, pathological fracture, or neurological deficit requires a broader strategy. Preoperative embolization may reduce vascularity, decompression addresses neural compression, and fixation may be required when structural stability is compromised. Cement augmentation can support the vertebral body and may assist hemostasis, but it does not replace these procedures when the epidural component is clinically significant [48,61,62,65]. Multidisciplinary planning between spine surgeons, interventional radiologists, and other relevant specialists is appropriate for these complex lesions.

Follow-up should reflect the original disease pattern. Clinical reassessment of pain and neurological function is reasonable after augmentation, with cross-sectional imaging when symptoms persist, recur, or were initially associated with aggressive features. Routine CT immediately after treatment improves detection of small leaks but exposes patients to additional radiation and may identify clinically irrelevant findings. The review cannot determine one universal imaging schedule. Longer surveillance is more relevant for aggressive lesions, incomplete filling, residual epidural disease, or previous retreatment.

### 4.5 Limitations

This review has several strengths. The search covered five databases without date or language restrictions and was updated shortly before manuscript completion. Study selection, extraction, and risk-of-bias assessment used prespecified procedures. Continuous pain change, categorical pain response, any radiographic leakage, symptomatic leakage, and recurrence-related outcomes were analyzed separately. Sensitivity, small-study effect, leave-one-out, and influence analyses were used to examine the stability of each pooled result. This outcome-specific approach avoided treating every detected cement leak as a clinically important complication.

The main limitation is the near absence of direct comparative evidence. Most studies were retrospective, uncontrolled, and conducted at single centers. The choice of vertebroplasty, kyphoplasty, or an adjunctive procedure was usually influenced by lesion anatomy and disease severity. Consequently, differences between treatment groups may reflect selection rather than the procedure itself. The one randomized trial evaluated a narrow surgical population and had several reporting limitations. It does not resolve the comparison between stand-alone vertebroplasty and kyphoplasty.

Clinical heterogeneity was substantial. Studies included non-aggressive painful lesions, aggressive lesions with cortical destruction, pathological fractures, and patients with epidural extension or neurological deficit. Some reports assessed augmentation alone, while others assessed complete multimodal pathways. Definitions of aggressive disease, pain response, leakage, recurrence, and retreatment varied. Follow-up ranged from months to more than a decade. These differences limit the direct applicability of a single pooled estimate to every clinical presentation.

Outcome ascertainment was another source of uncertainty. CT detects small leaks more sensitively than fluoroscopy or plain radiography, but postoperative imaging was not standardized. Neurological and functional outcomes were often described without validated scales. Pain was assessed at different time points, and categorical response definitions were inconsistent. Recurrence was sometimes defined by renewed pain, sometimes by imaging progression, and sometimes by the need for another procedure. No study used a consistent volumetric method for intraosseous or epidural response.

Several pooled outcomes contained few events. Symptomatic leakage was based on four events and recurrence-related outcomes on ten events. Confidence intervals, subgroup tests, Egger regression, and trim-and-fill are unstable in this setting. Funnel-plot asymmetry may reflect true heterogeneity, different surveillance methods, or chance rather than publication bias alone. The adjusted trim-and-fill estimates were therefore treated as exploratory and should not replace the observed pooled proportions.

The analysis used aggregate study data. Patient-level relationships among lesion level, aggressive morphology, cement volume, adjunctive treatment, and outcome could not be examined. Several large procedural cohorts reported only small hemangioma subgroups, while some dedicated series did not supply all required denominators or variance measures.

Kyphoplasty data were particularly sparse. Publication and selective outcome reporting remain possible because favorable pain outcomes may be more likely to appear in small case series than unsuccessful procedures or late complications.

### 4.6 Future research

Future studies should use prospective multicenter designs with clearly defined symptomatic and aggressive disease categories. A direct comparison of vertebroplasty and kyphoplasty would be valuable, but recruitment may be difficult because symptomatic vertebral hemangioma is uncommon and procedural selection depends strongly on anatomy. A well-designed registry with prospective enrollment, standardized variables, and treatment-specific follow-up may be more feasible than a single-center randomized trial.

Outcome definitions require harmonization. Pain should be recorded with a validated scale at baseline and at prespecified early and late time points. Disability, analgesic use, neurological status, and health-related quality of life should be reported alongside pain. Complete relief, partial response, recurrence, progression, retreatment, and reoperation need explicit definitions. Reports should provide patient denominators at every assessment and distinguish missing follow-up from absence of an event.

Procedural reporting should include access route, cement type and viscosity, injected volume, injection endpoint, use of venography, cavity creation, cortical defects, epidural extension, and all adjunctive procedures. Leakage should be classified by anatomical location and clinical consequence. Immediate imaging findings should be separated from symptoms requiring medication, intervention, decompression, or intensive monitoring. Such detail would allow future analyses to examine which technical factors are associated with pain control and clinically relevant complications.

Standardized CT or MRI criteria would improve assessment of epidural and intraosseous response. Studies of aggressive disease should document the size of the epidural component, spinal canal compromise, vertebral collapse, and neurological grade before and after treatment. Longer follow-up is required to capture recurrence and later retreatment. Central review of imaging and individual-participant-data collaboration could help identify predictors that cannot be evaluated from published aggregate data.

Research should also separate pain-predominant lesions from lesions requiring decompression for neurological compromise. These populations have different treatment goals and should not be combined without stratification. For the first group, the main questions concern pain response, leakage, and the relative role of vertebroplasty and kyphoplasty. For the second group, studies should evaluate the sequence and contribution of embolization, augmentation, decompression, and fixation within the overall treatment pathway.

## 5. Conclusion

Vertebral augmentation is associated with substantial and often rapid pain relief in patients with symptomatic vertebral hemangioma. Radiographic cement leakage occurs more often than clinically important leakage, while symptomatic leakage and recurrence-related events are uncommon in the published literature. These estimates are derived mainly from retrospective vertebroplasty series with variable imaging and follow-up. Kyphoplasty-specific evidence remains too sparse to establish a comparative advantage or a reliable difference in leakage. Vertebroplasty has the broader evidence base for pain-predominant lesions without major neurological compromise. Aggressive lesions with epidural extension, instability, or neurological deficit usually require augmentation within a multimodal strategy that may include embolization, decompression, and fixation. Treatment selection should be individualized according to lesion anatomy and clinical presentation. Prospective multicenter studies with standardized pain, functional, imaging, and recurrence outcomes are needed to define the relative role of each augmentation technique.

## Declarations

## Acknowledgements

None.

## Preprint statement

This manuscript has not been posted as a preprint.

## Funding

The authors received no financial support for the research, authorship, or publication of this article.

## Competing interests

The authors declare that they have no competing interests.

## Ethics approval

Ethics approval was not required because this study is a systematic review and meta-analysis of published data.

## Consent to participate

Not applicable.

## Consent for publication

Not applicable.

## Data availability

The included-study list, extraction workbook, search strategies, and risk-of-bias checklists are supplied as supplementary files. Additional materials are available from the corresponding author on reasonable request.

## Declaration of generative AI and AI-assisted technologies

During preparation of this work, the authors used ChatGPT by OpenAI for language refinement and translation support. The authors reviewed and edited all output, verified the scientific content and references against the source material, and take full responsibility for the manuscript.

## Author contributions

F.F.: conceptualization, methodology, supervision, validation, and writing - review and editing. M.J.: data curation, formal analysis, visualization, and writing - original draft and review and editing. F.M.M.: methodology, investigation, study selection, risk-of-bias assessment, validation, and writing - review and editing. A.M., M.F.L., Z.M., A.R., and S.S.: investigation, data curation, validation, and writing - review and editing. A.H. and A.K.: data extraction, data curation, and writing - review and editing; A.K. also contributed to risk-of-bias assessment. M.K., F.E., and R.Y.: title and abstract screening, investigation, and writing - review and editing. A.H.N.: full-text assessment, investigation, and writing - review and editing. A.Z.: supervision, resources, and critical revision of the manuscript. All authors read and approved the final manuscript.

## Supplementary figures

*The following figures provide subgroup, sensitivity, small-study effect, and influence analyses for the five pooled outcomes. Each caption states the main result shown in the figure*.

**Figure S1.**
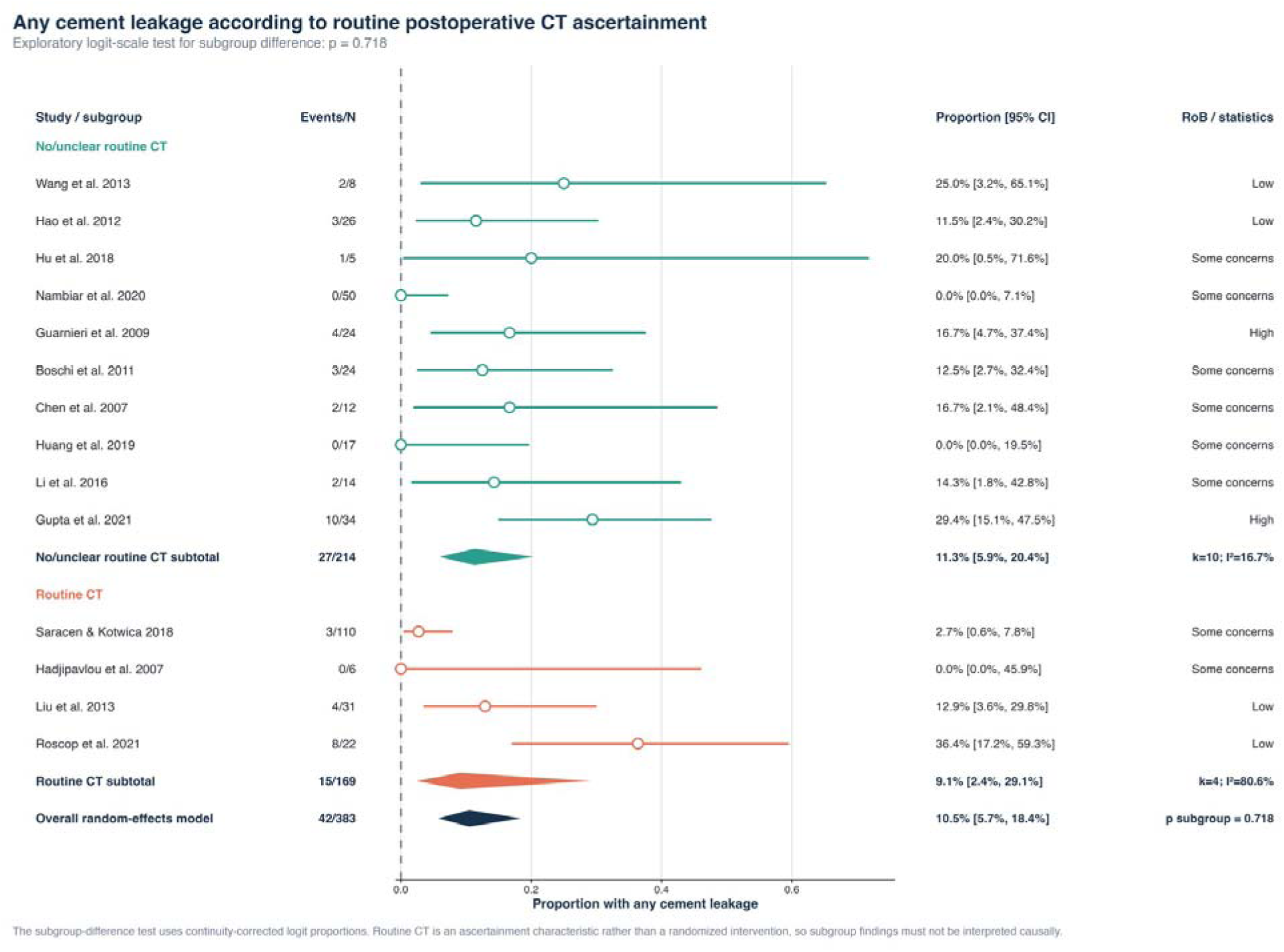
Subgroup analysis of any cement leakage by routine postoperative CT ascertainment. *The pooled proportions were 11.3% with routine CT and 9.1% with no or unclear routine CT. The subgroup difference was not significant (p=0.718)*.

**Figure S2.**
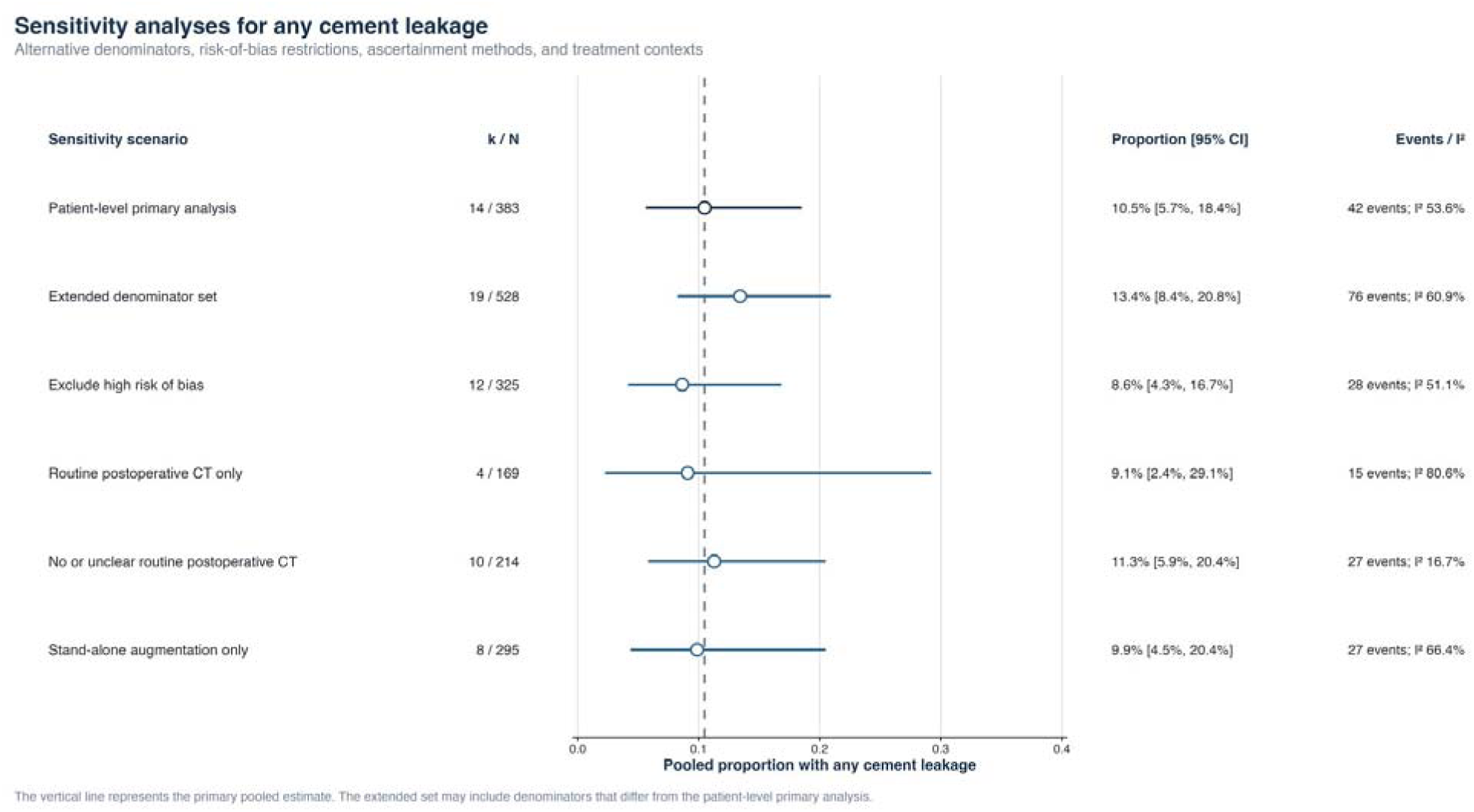
Sensitivity analyses for any cement leakage. *Alternative denominators, risk-of-bias restrictions, ascertainment methods, and treatment contexts produced pooled estimates from 8.6% to 13.4%*.

**Figure S3.**
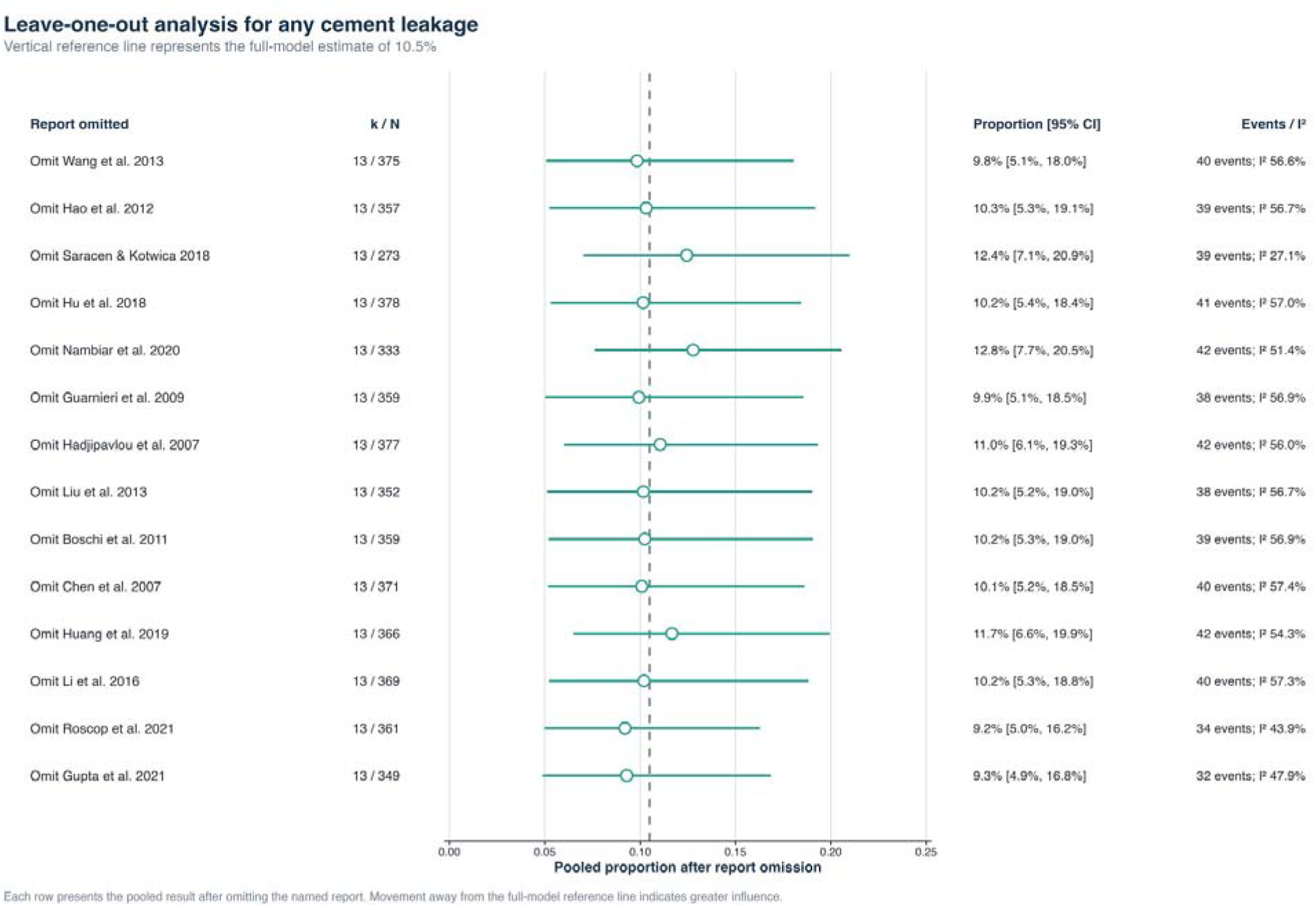
Leave-one-out analysis for any cement leakage. *Removing one study at a time gave pooled estimates from 9.2% to 12.8%, without changing the main interpretation*.

**Figure S4.**
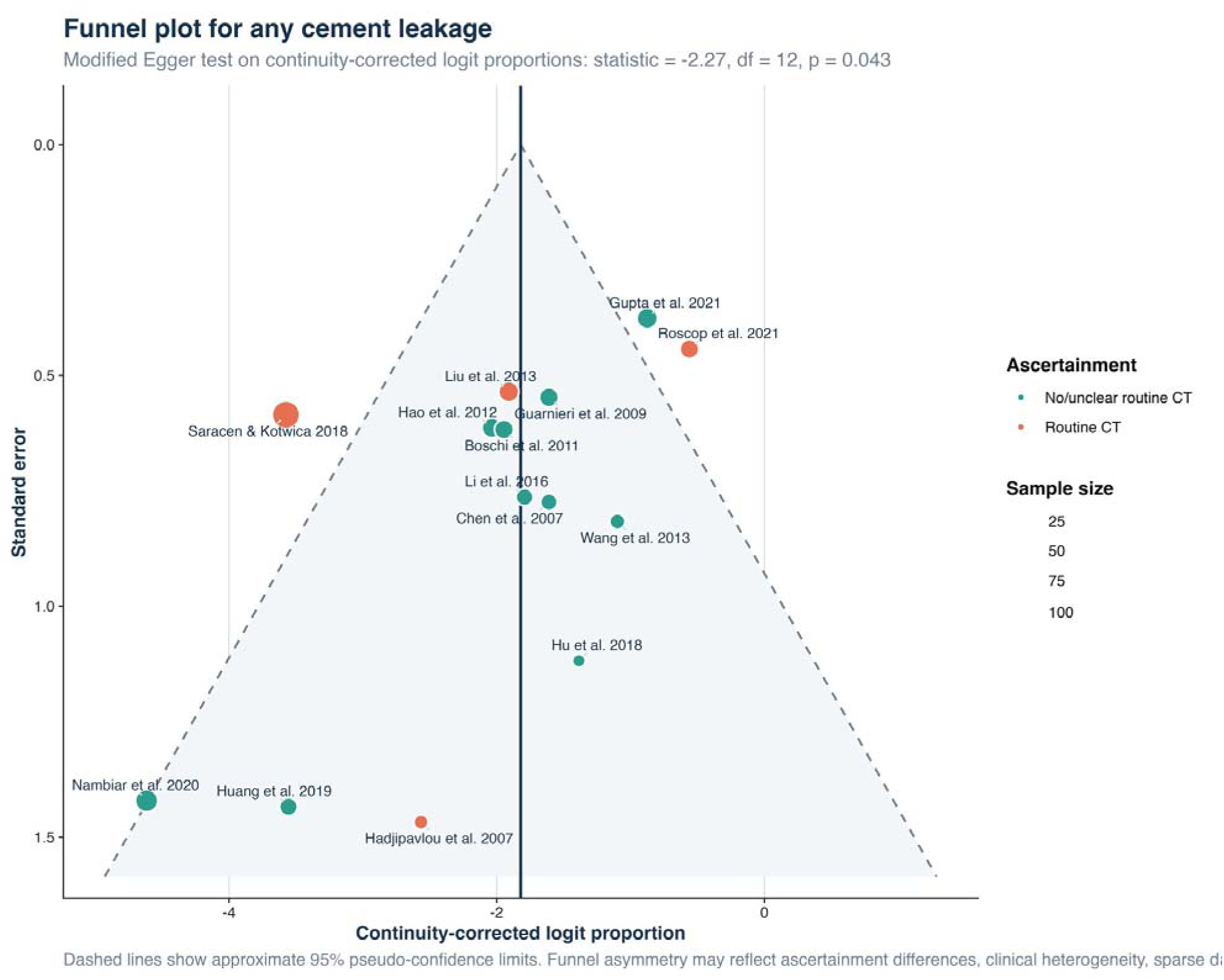
Funnel plot for any cement leakage. *The plot showed asymmetry, consistent with the modified Egger test result (p=0.043)*.

**Figure S5.**
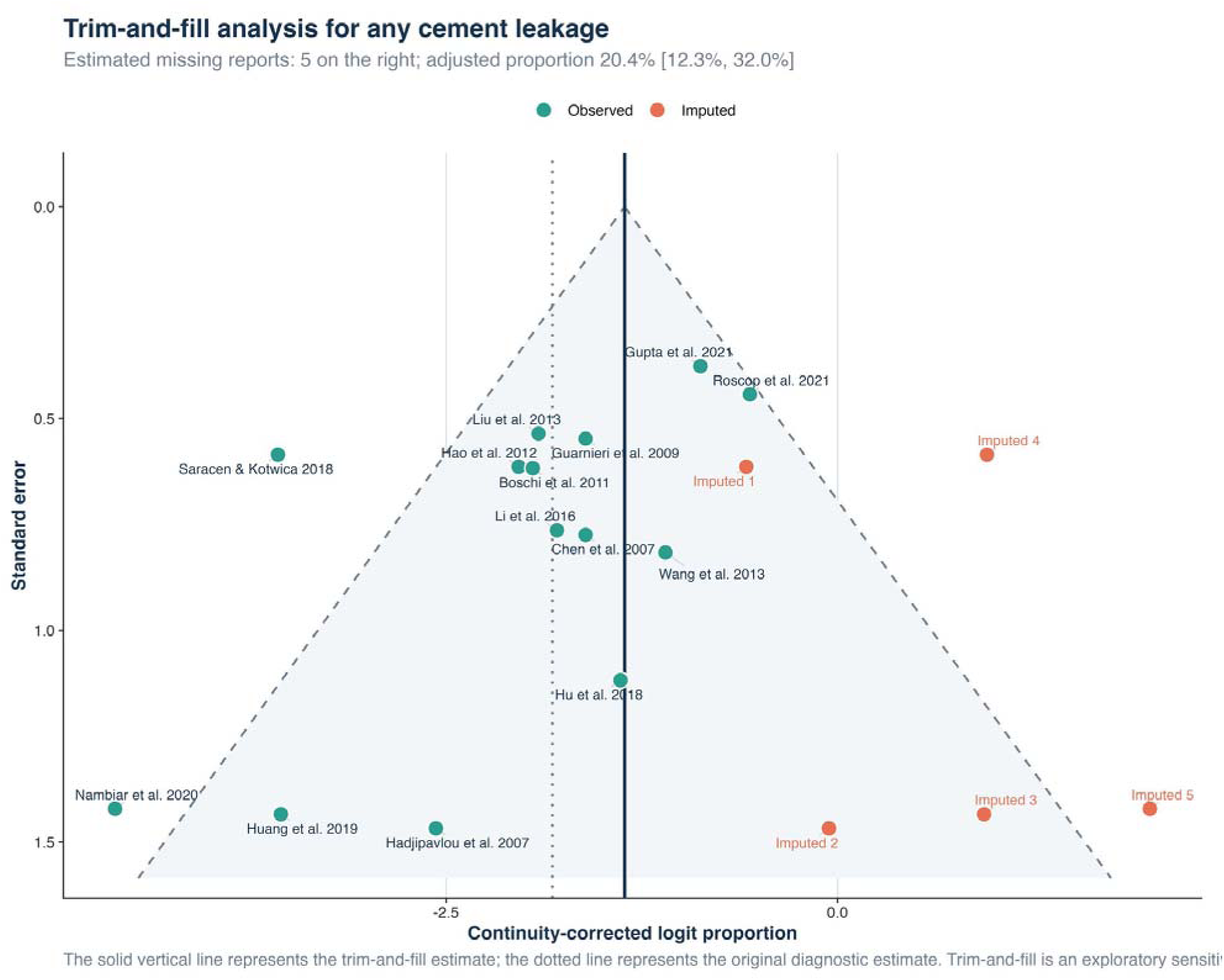
Trim-and-fill analysis for any cement leakage. *Five studies were imputed on the right side, giving an exploratory adjusted estimate of 20.4% (95% CI 12.3-32.0%)*.

**Figure S6.**
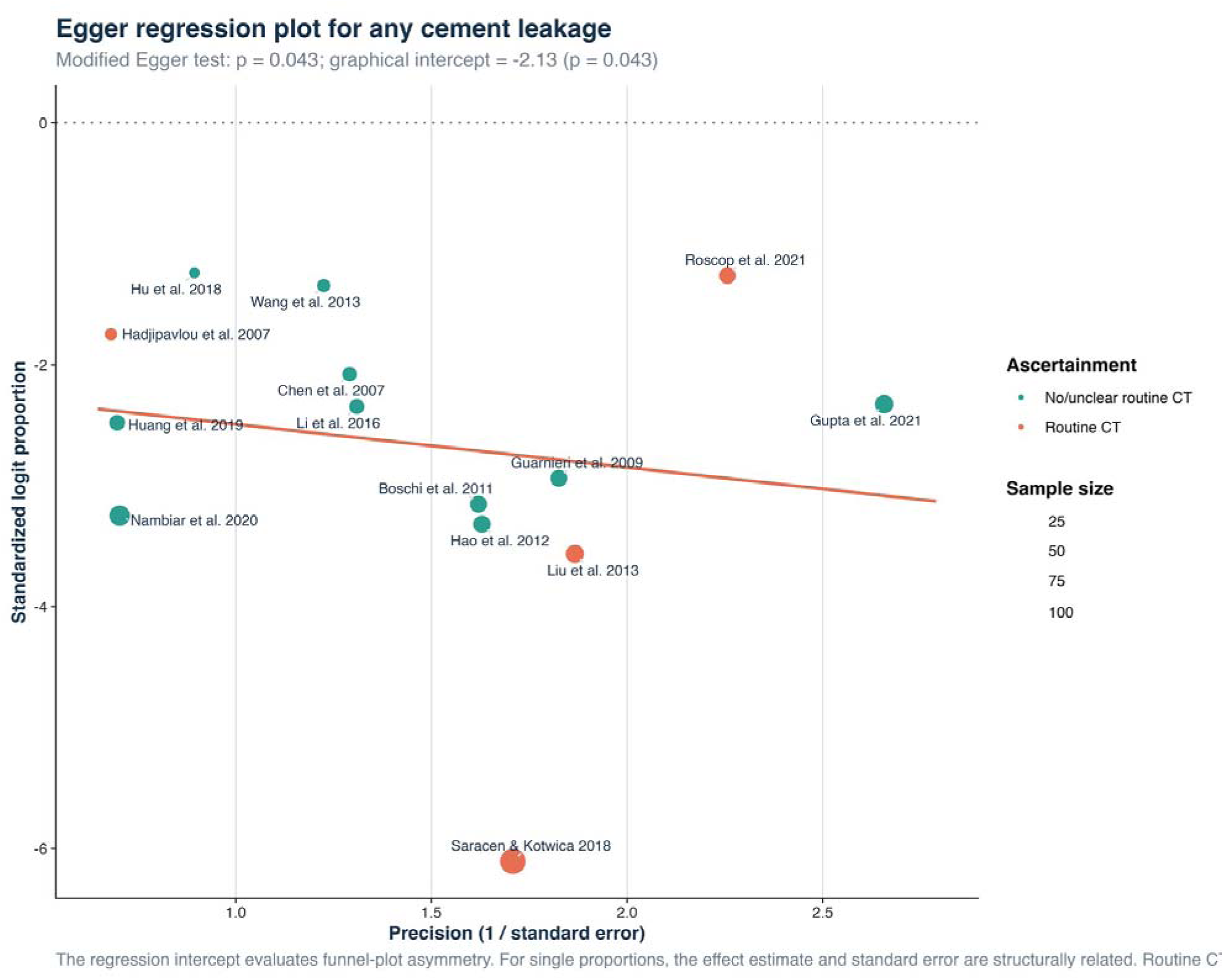
Egger regression for any cement leakage. *The modified Egger test was significant at p=0.043, suggesting possible small-study effects or under-ascertainment*.

**Figure S7.**
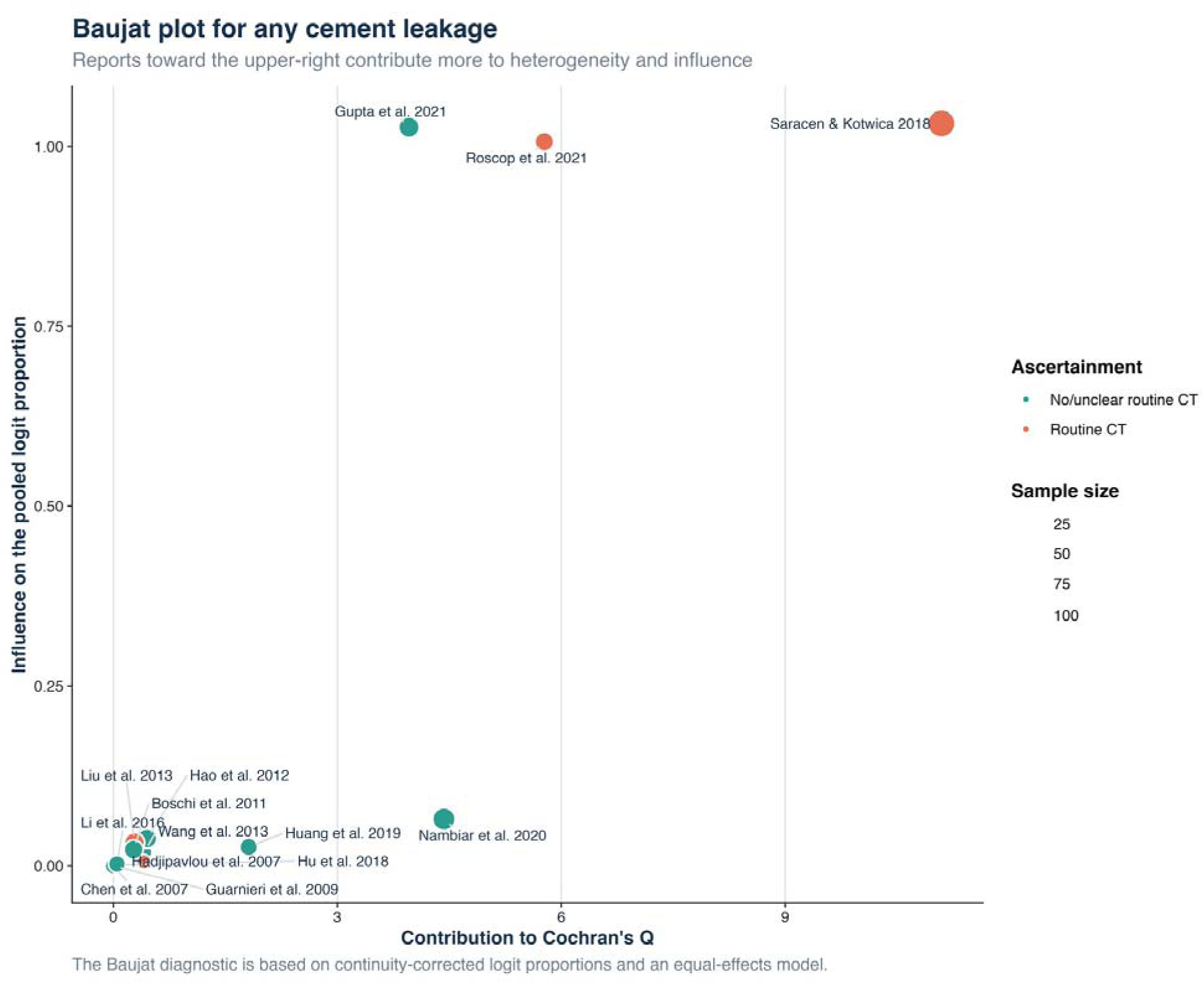
Baujat plot for any cement leakage. *Saracen et al. contributed most to influence, while Gupta et al. and Roscop et al. contributed to heterogeneity at higher leakage rates*.

**Figure S8.**
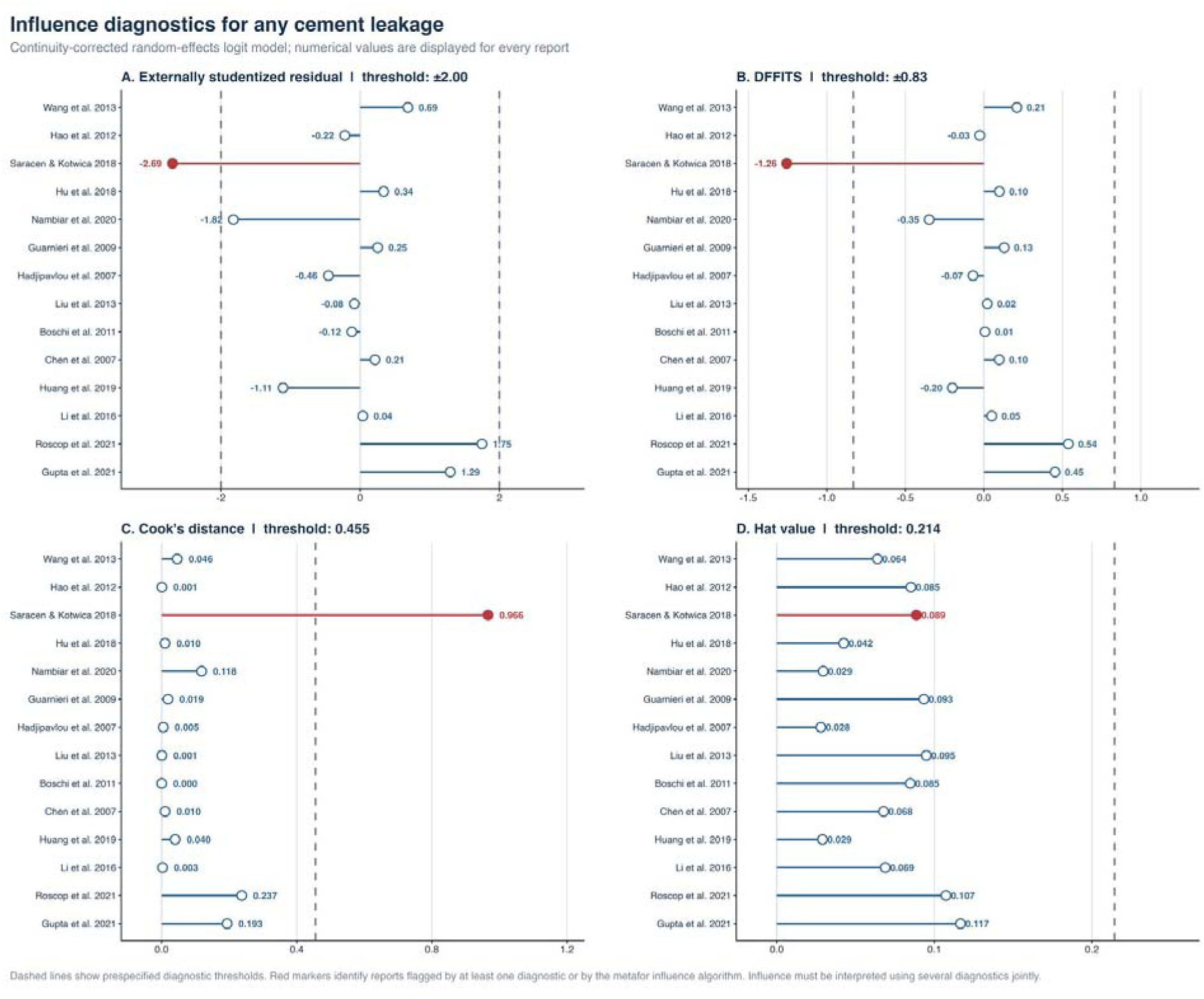
Influence diagnostics for any cement leakage. *Saracen et al. exceeded the prespecified thresholds for the externally studentized residual and Cook’s distance*.

**Figure S9.**
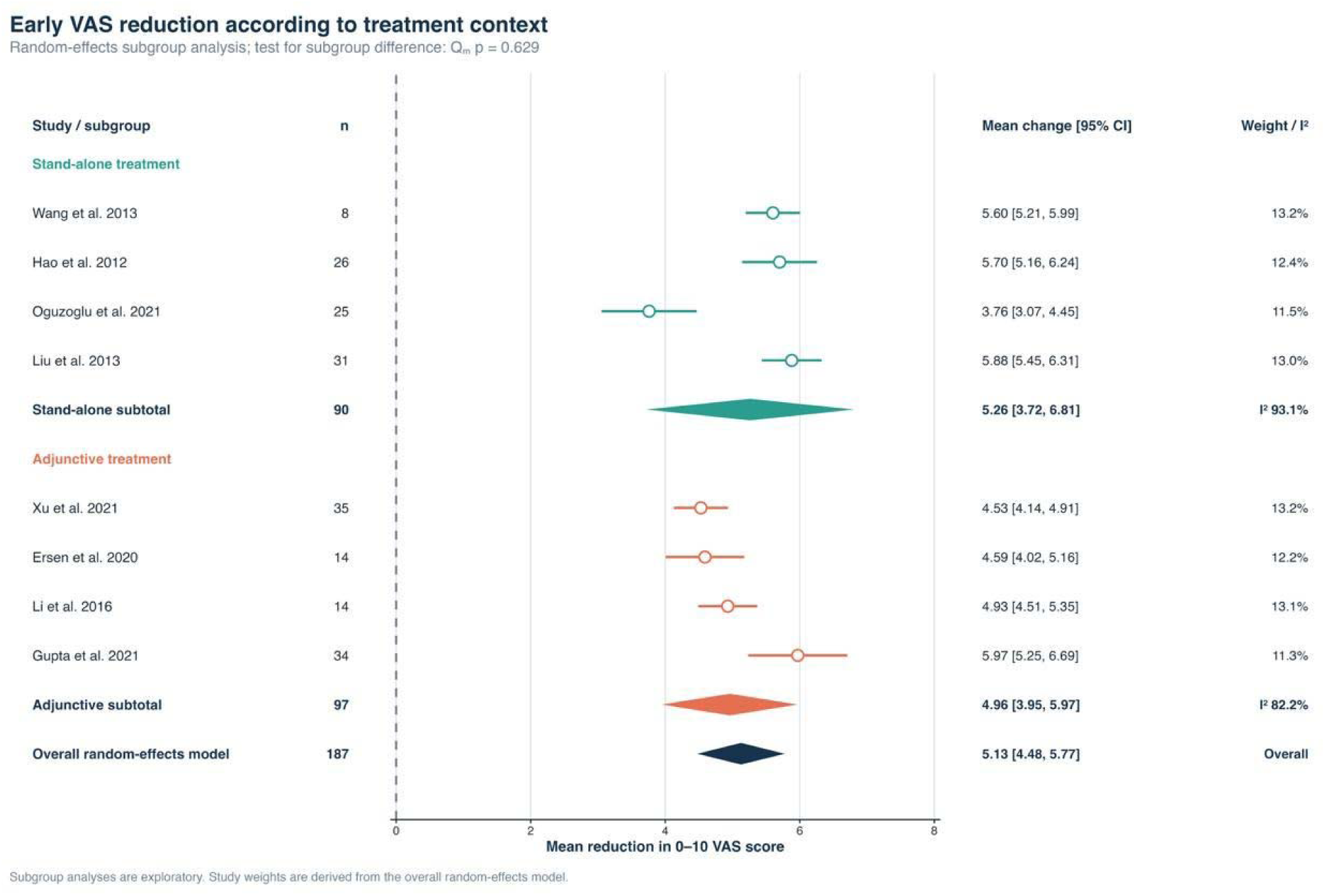
Subgroup analysis of early VAS reduction by treatment context. *Mean reductions were 5.26 points for stand-alone treatment and 4.96 points for adjunctive or combined treatment (p=0.629)*.

**Figure S10.**
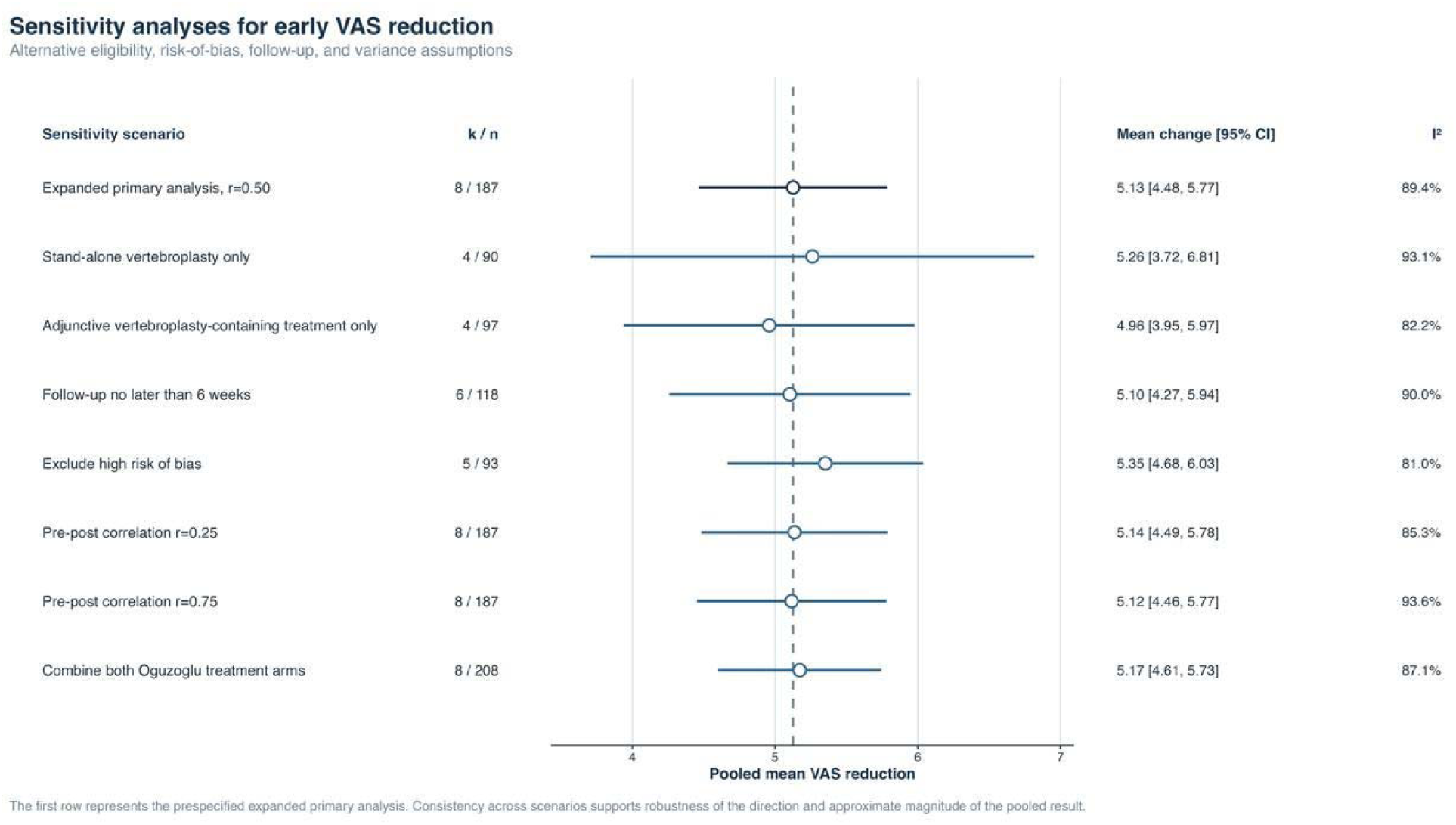
Sensitivity analyses for early VAS reduction. *Alternative eligibility, follow-up, risk-of-bias, and pre-post correlation assumptions produced estimates from 5.10 to 5.35 points*.

**Figure S11.**
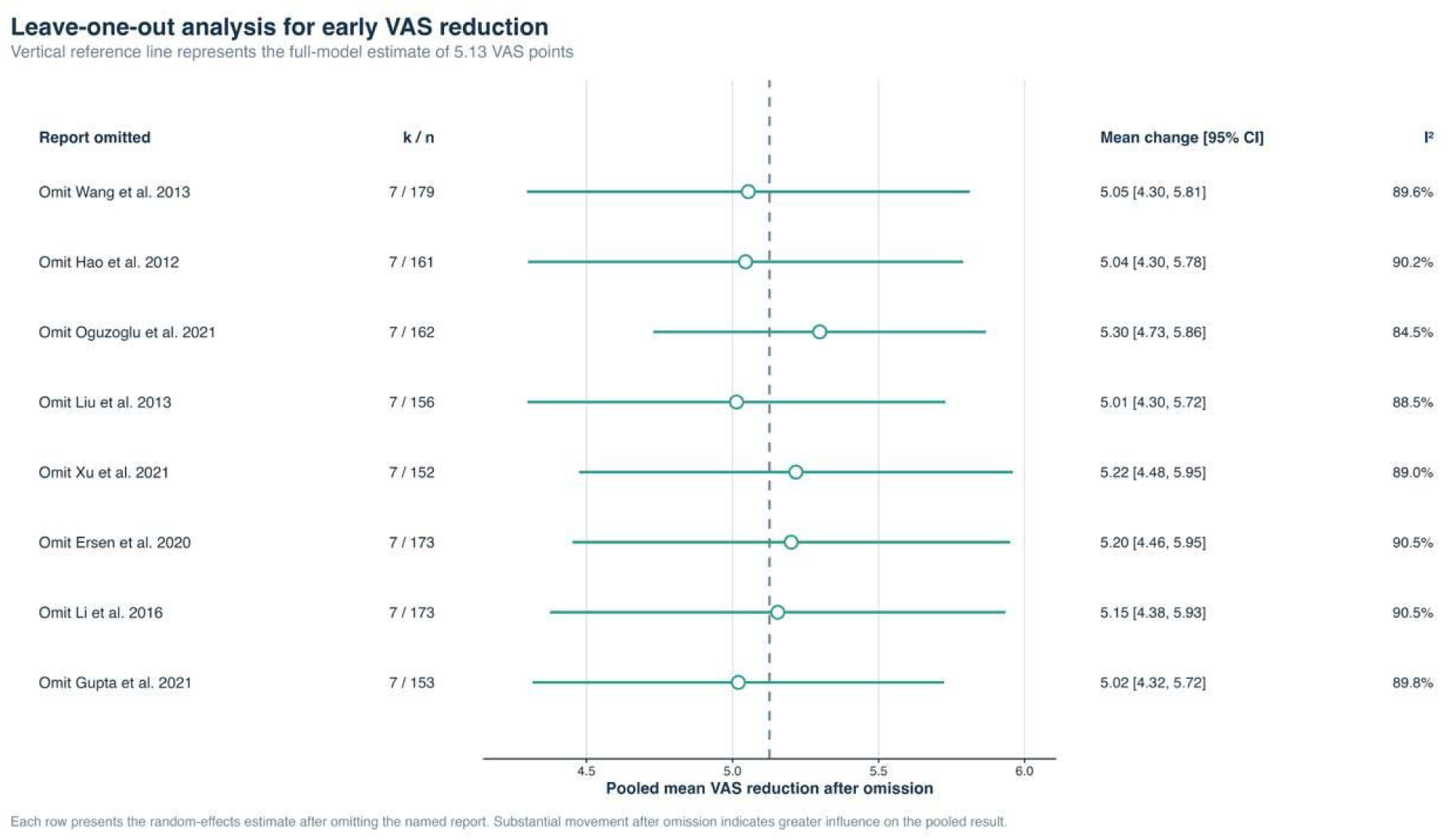
Leave-one-out analysis for early VAS reduction. *No single study materially changed the pooled pain reduction*.

**Figure S12.**
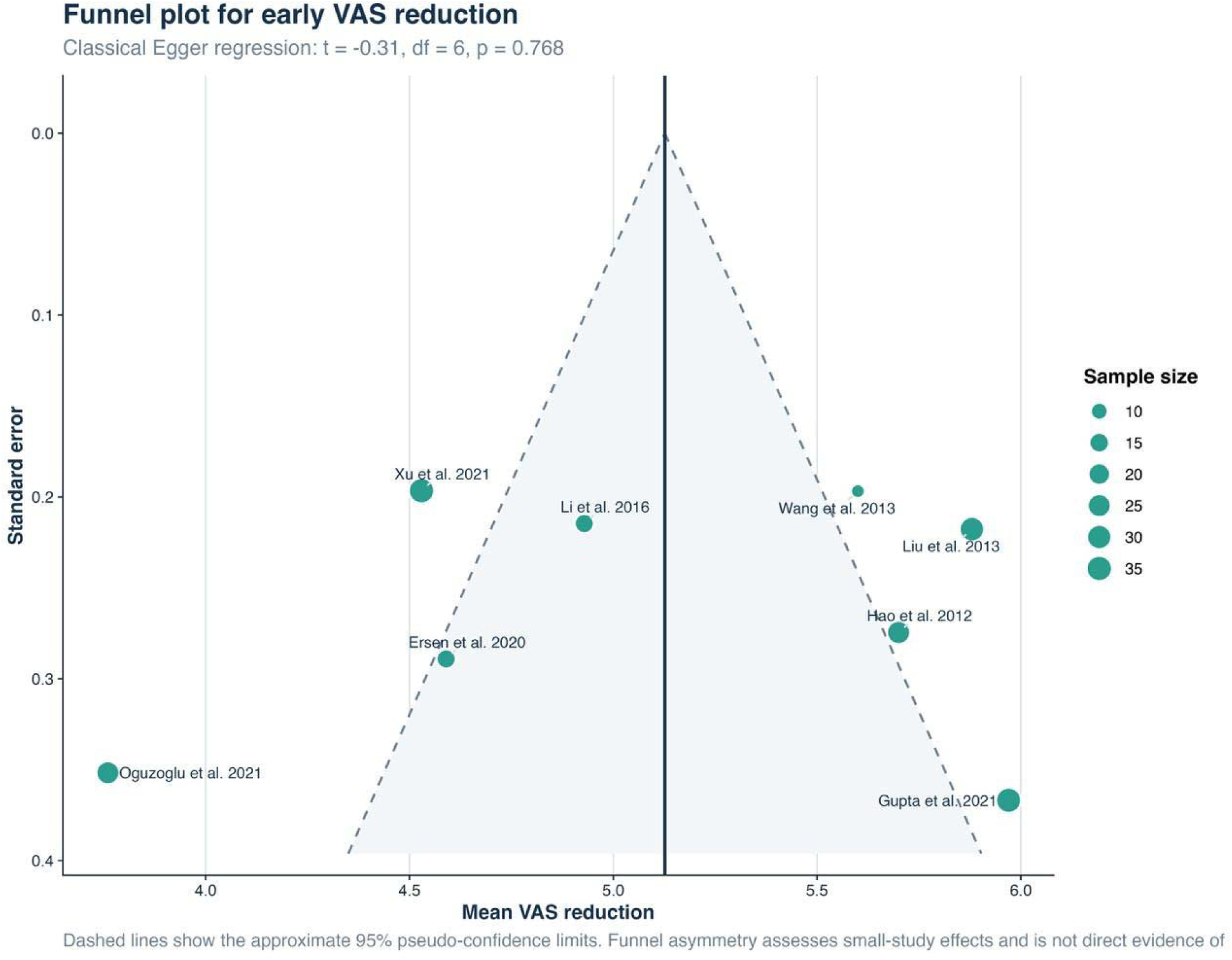
Funnel plot for early VAS reduction. *Visual asymmetry was limited and Egger regression was not significant (p=0.768)*.

**Figure S13.**
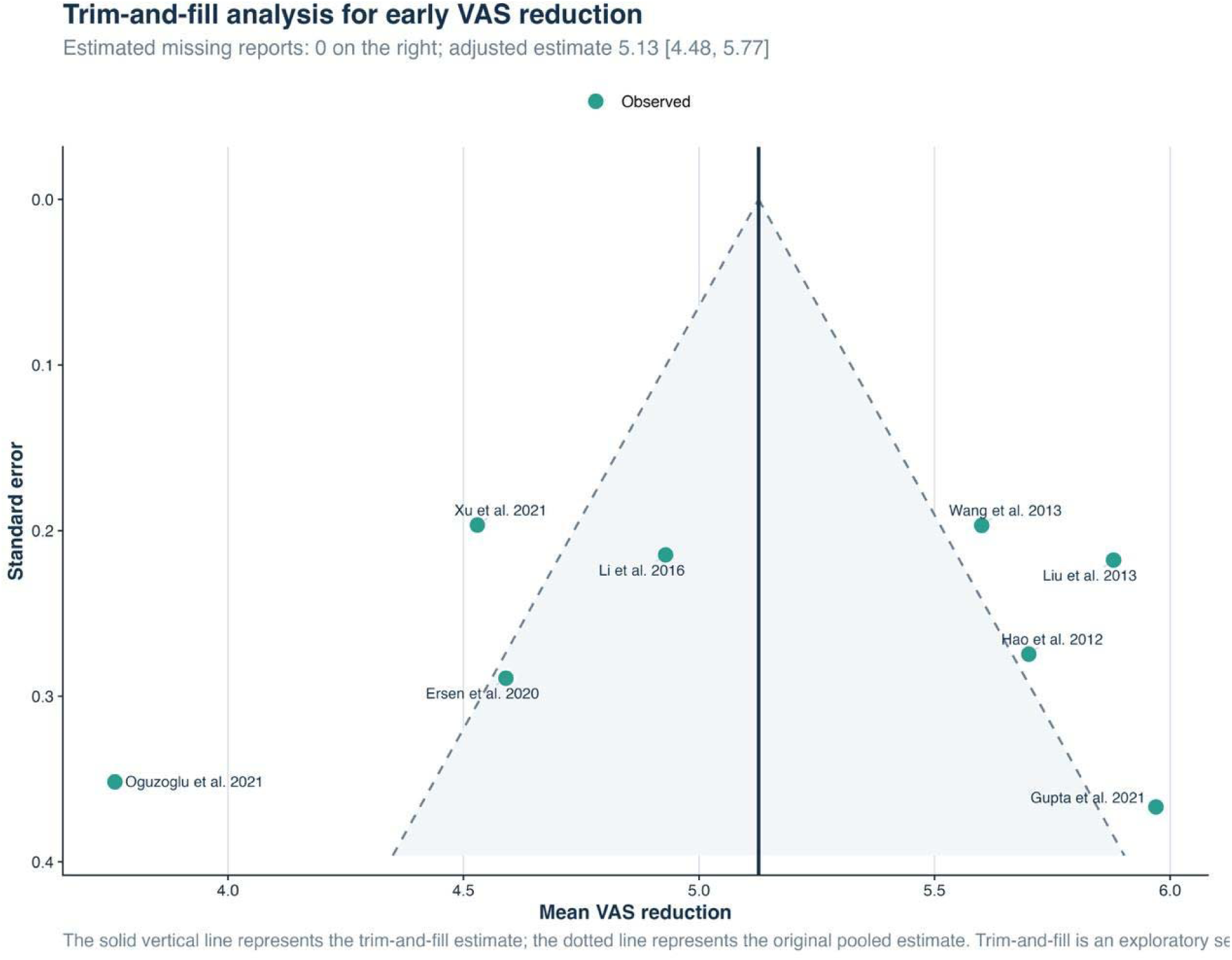
Trim-and-fill analysis for early VAS reduction. *No missing study was imputed and the pooled estimate was unchanged*.

**Figure S14.**
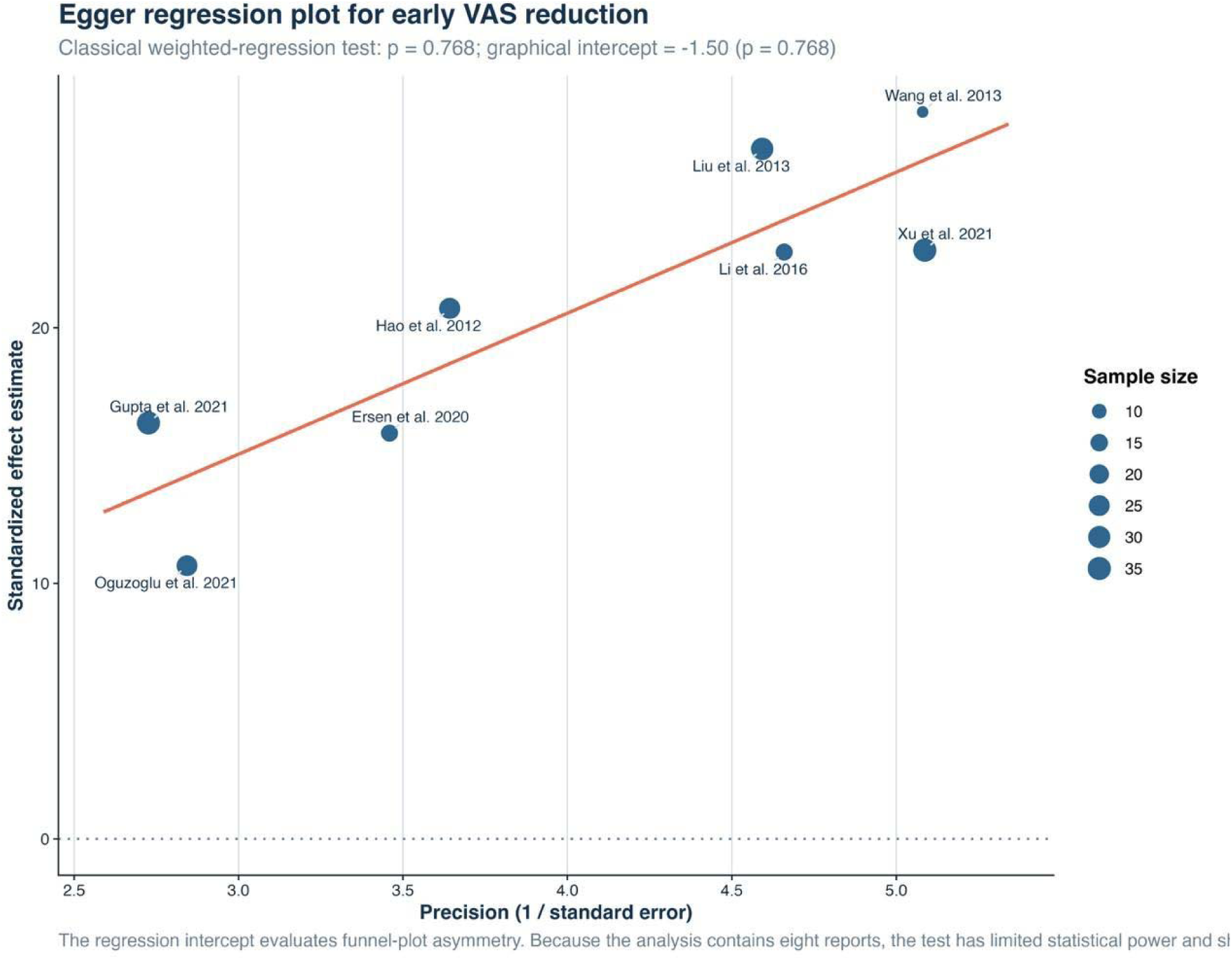
Egger regression for early VAS reduction. *The regression test did not indicate small-study effects (p=0.768)*.

**Figure S15.**
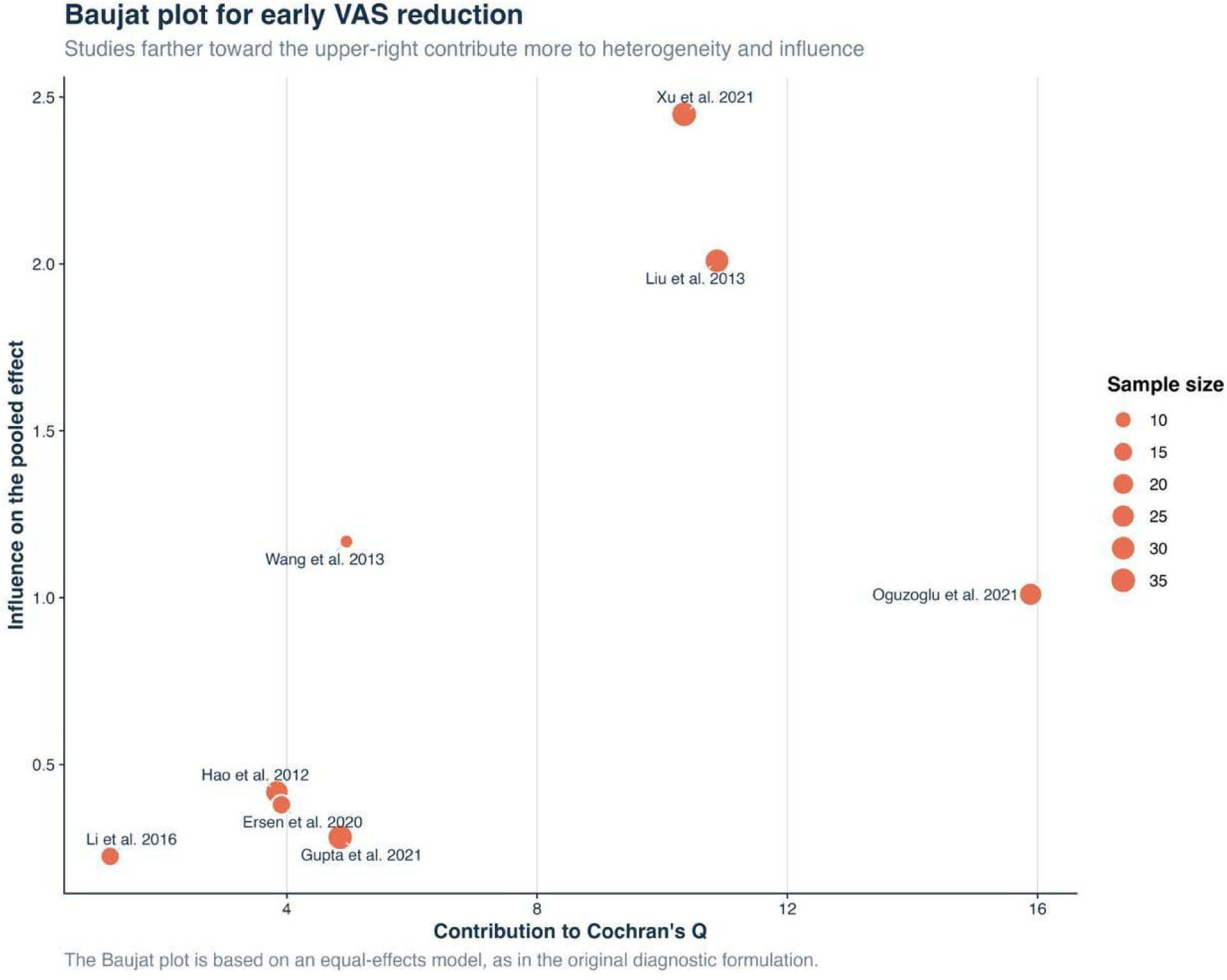
Baujat plot for early VAS reduction. *Xu et al. and Liu et al. were the largest contributors to between-study heterogeneity*.

**Figure S16.**
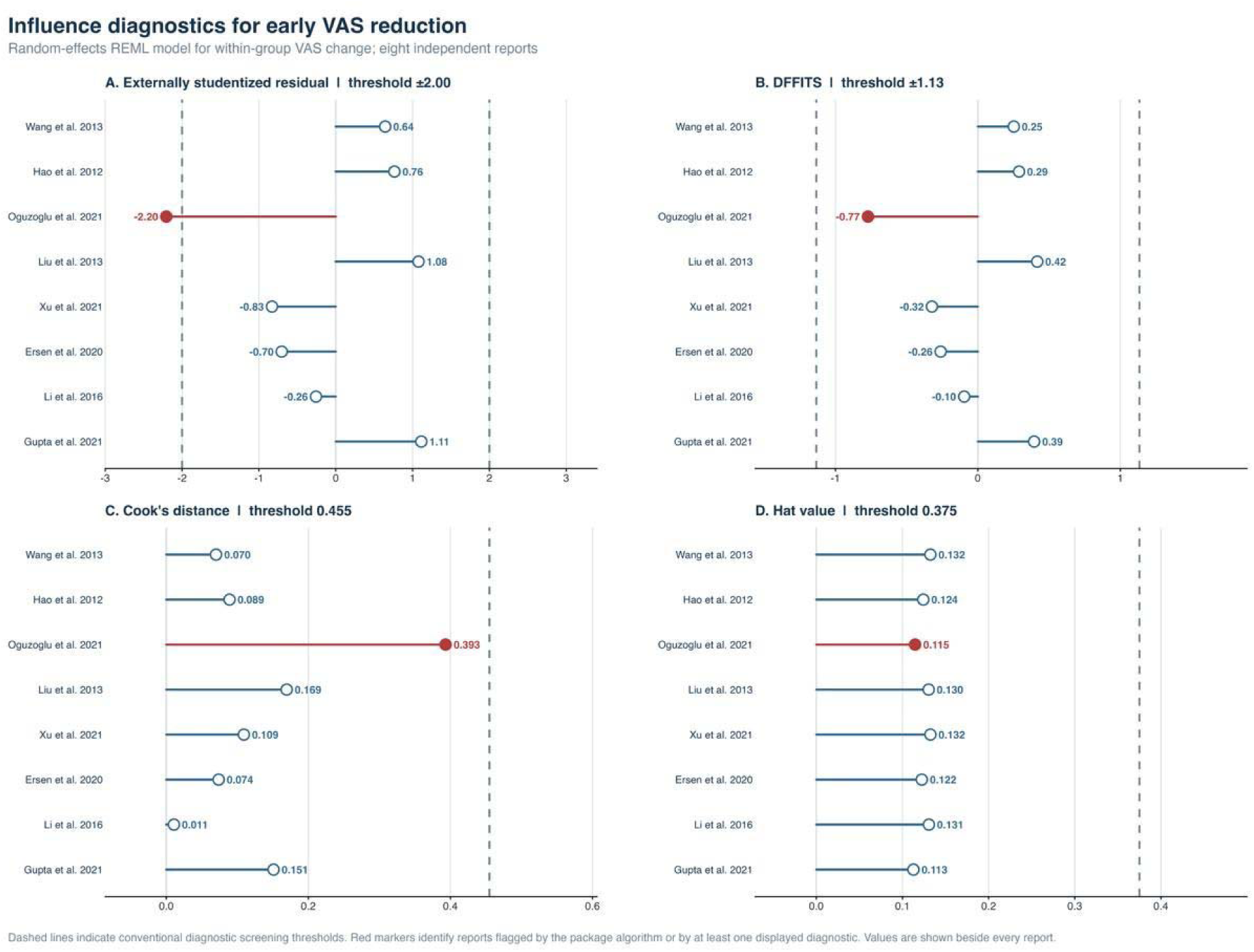
Influence diagnostics for early VAS reduction. *Oguzoglu et al. was flagged by an externally studentized residual of -2.20*.

**Figure S17.**
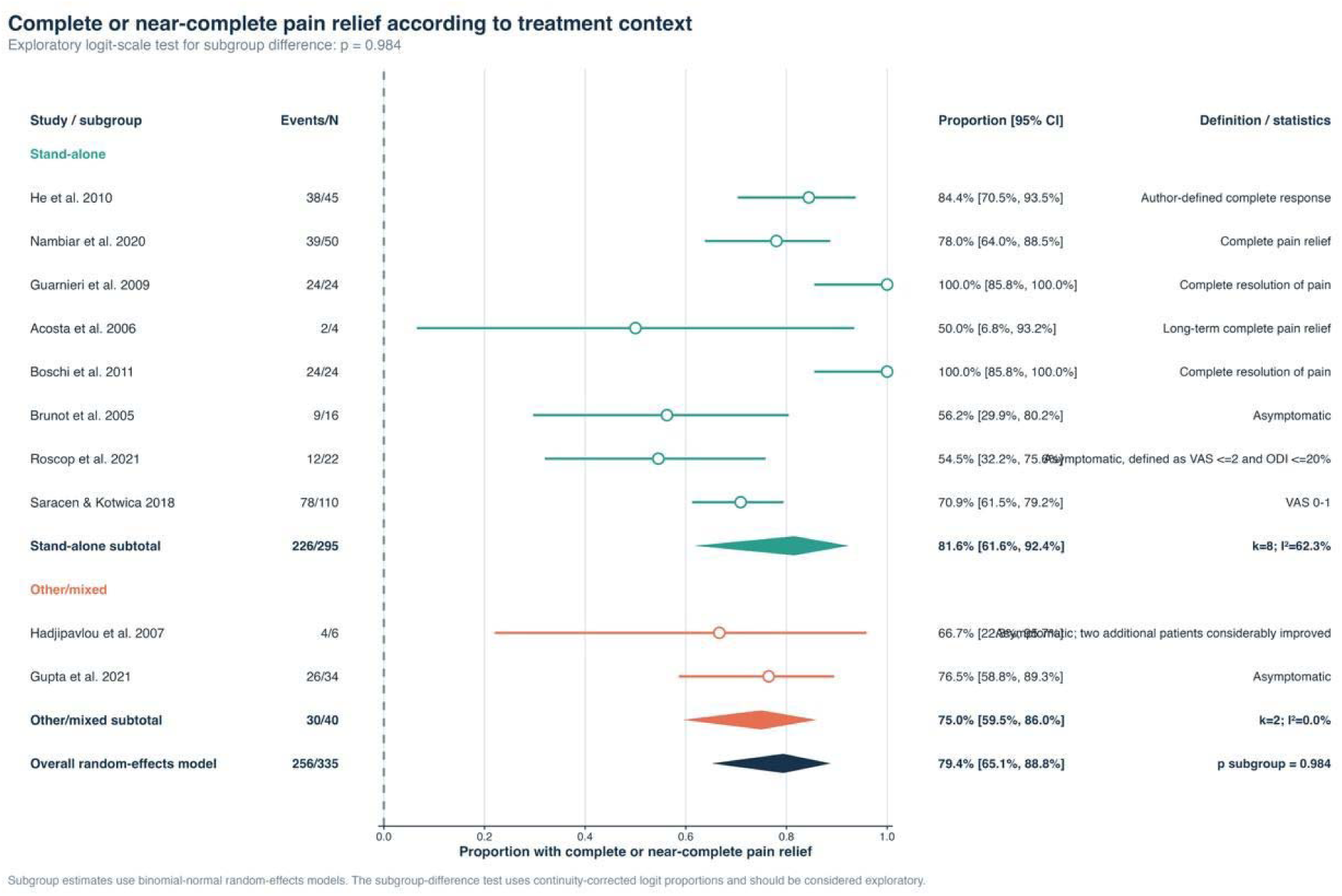
Subgroup analysis of complete or near-complete pain relief by treatment context. *Pooled relief was 81.6% for stand-alone treatment and 75.0% for other or mixed treatment (p=0.984)*.

**Figure S18.**
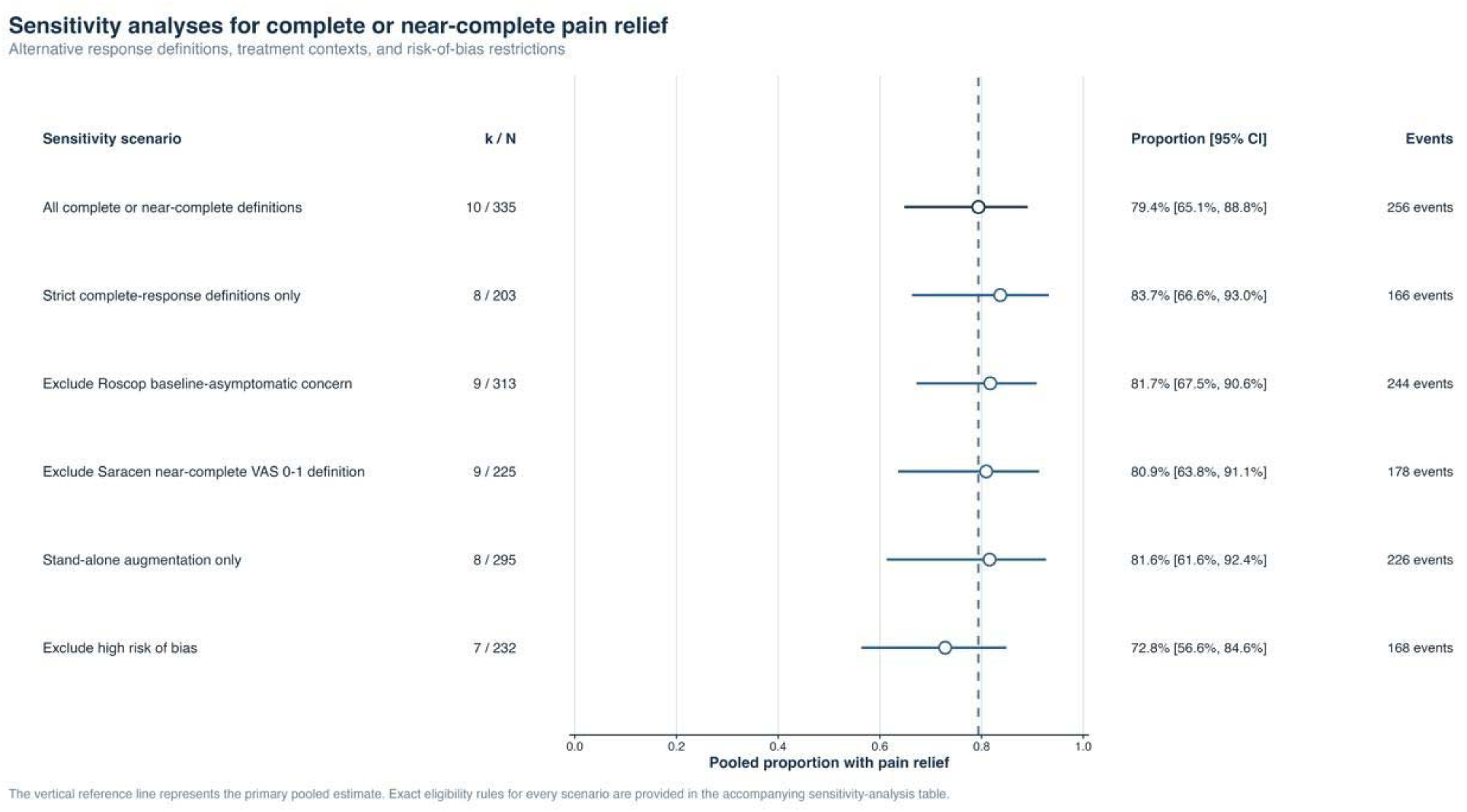
Sensitivity analyses for complete or near-complete pain relief. *Alternative response definitions and study restrictions produced estimates from 72.8% to 83.7%*.

**Figure S19.**
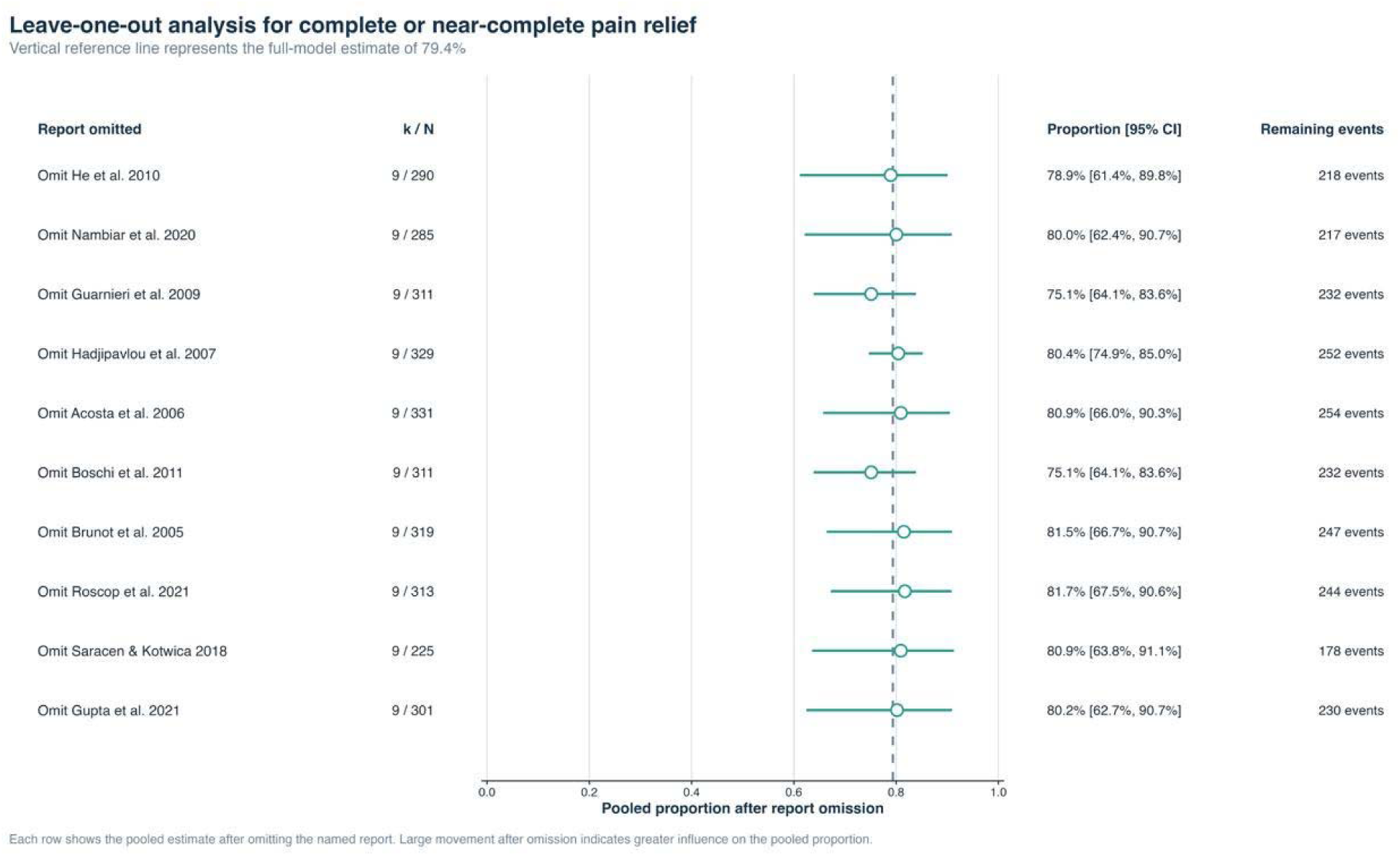
Leave-one-out analysis for complete or near-complete pain relief. *Pooled estimates ranged from 75.1% to 81.7%, with no dominant report*.

**Figure S20.**
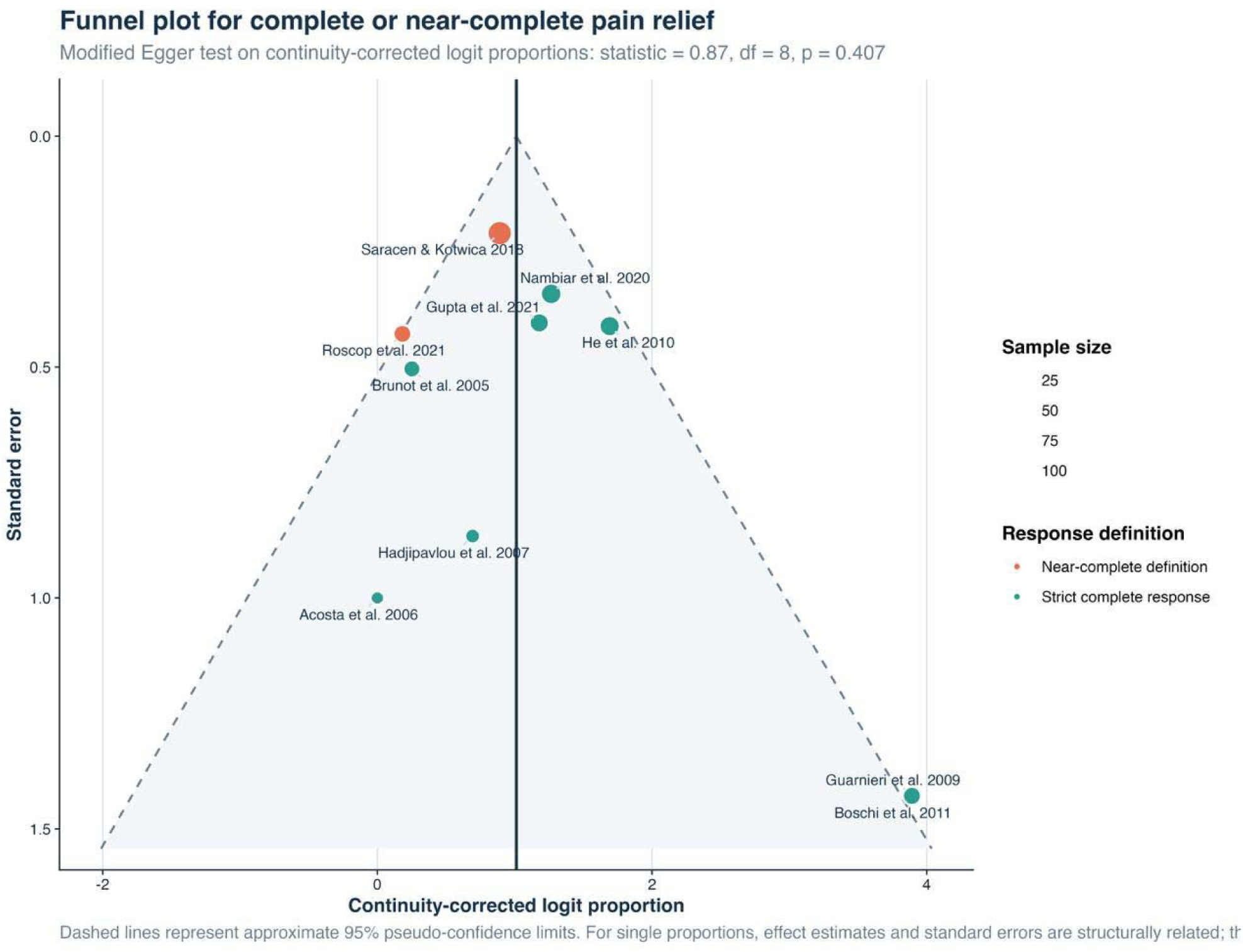
Funnel plot for complete or near-complete pain relief. *The plot did not show a clear small-study pattern; Egger regression was not significant (p=0.407)*.

**Figure S21.**
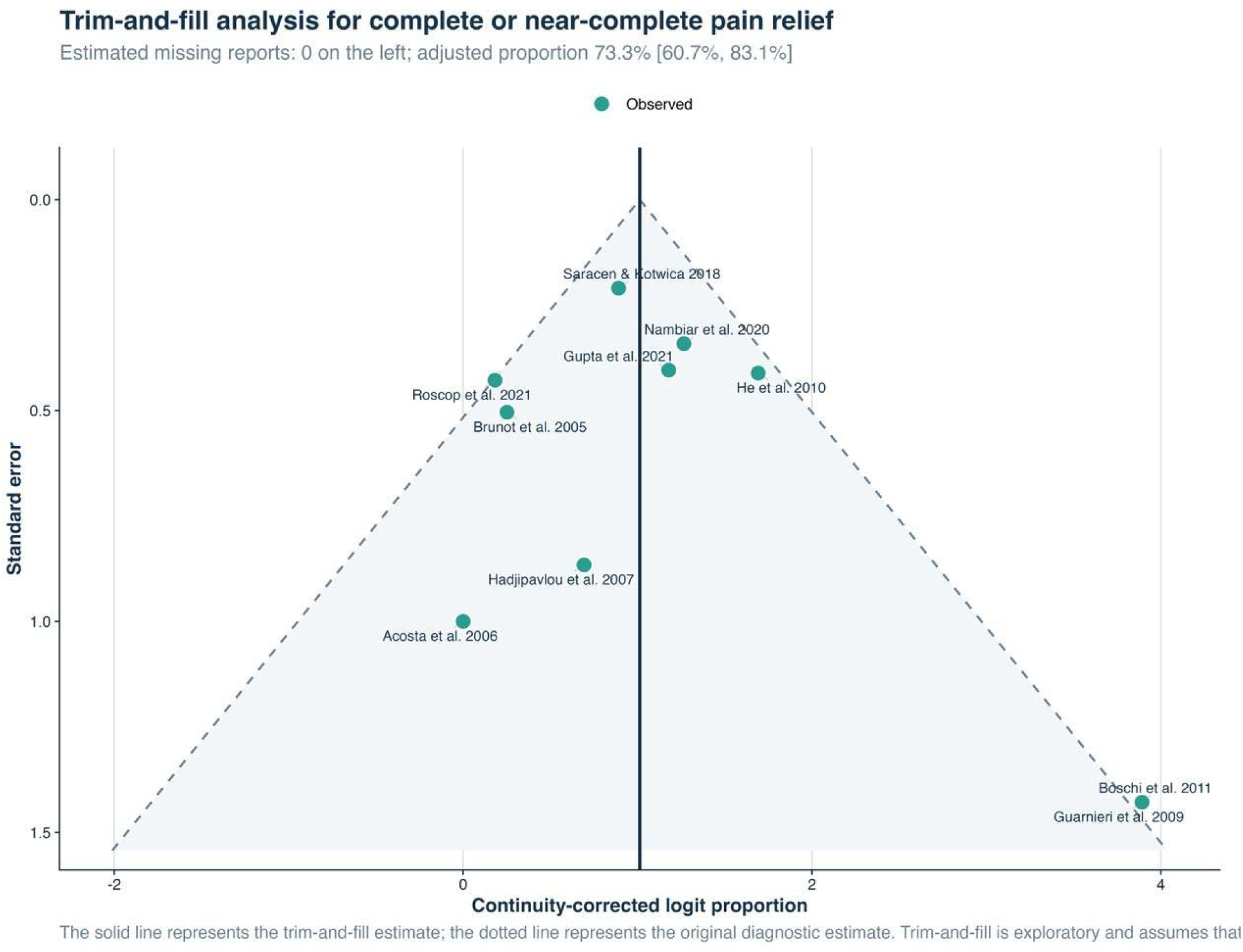
Trim-and-fill analysis for complete or near-complete pain relief. *No missing study was imputed*.

**Figure S22.**
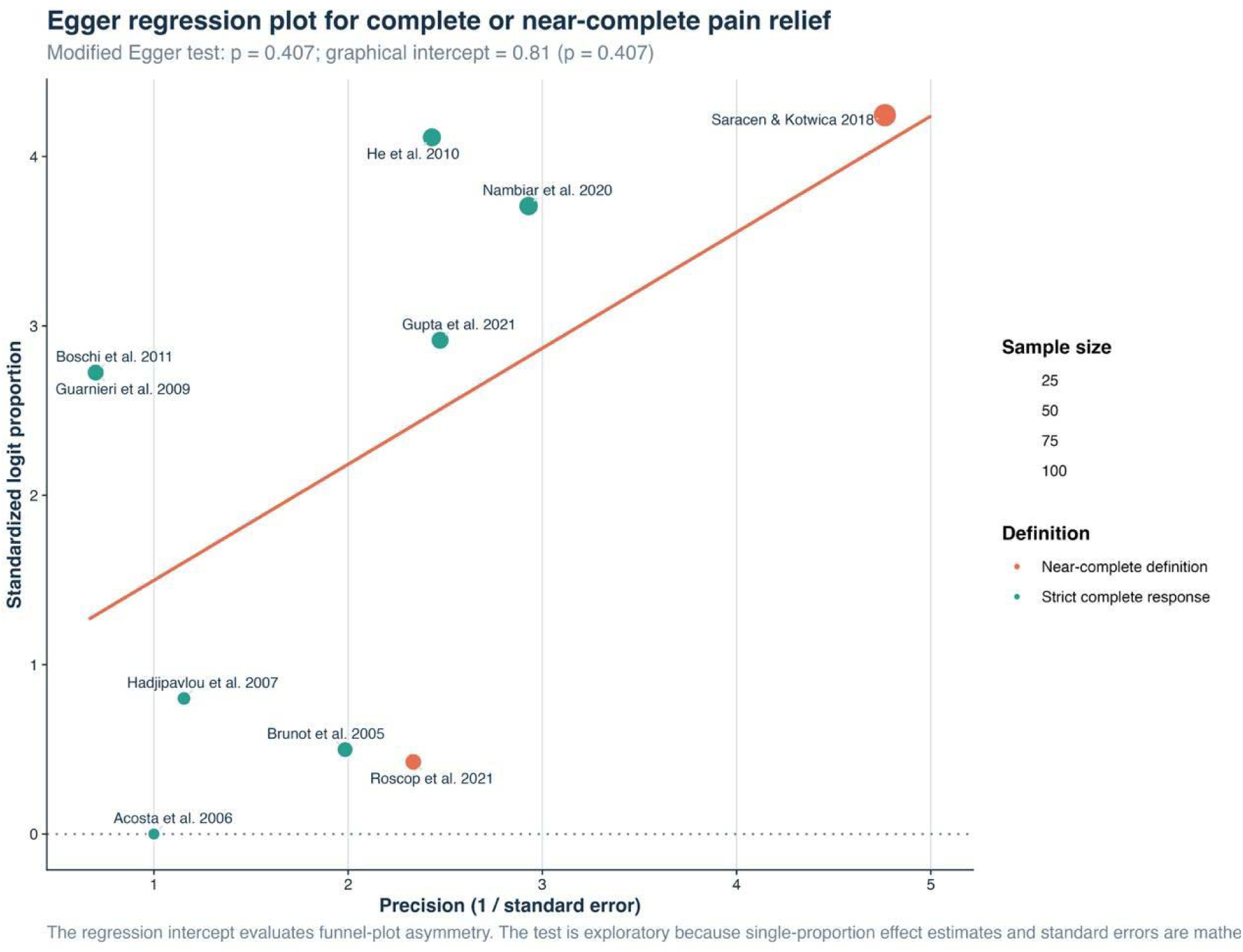
Egger regression for complete or near-complete pain relief. *The test did not indicate significant small-study effects (p=0.407)*.

**Figure S23.**
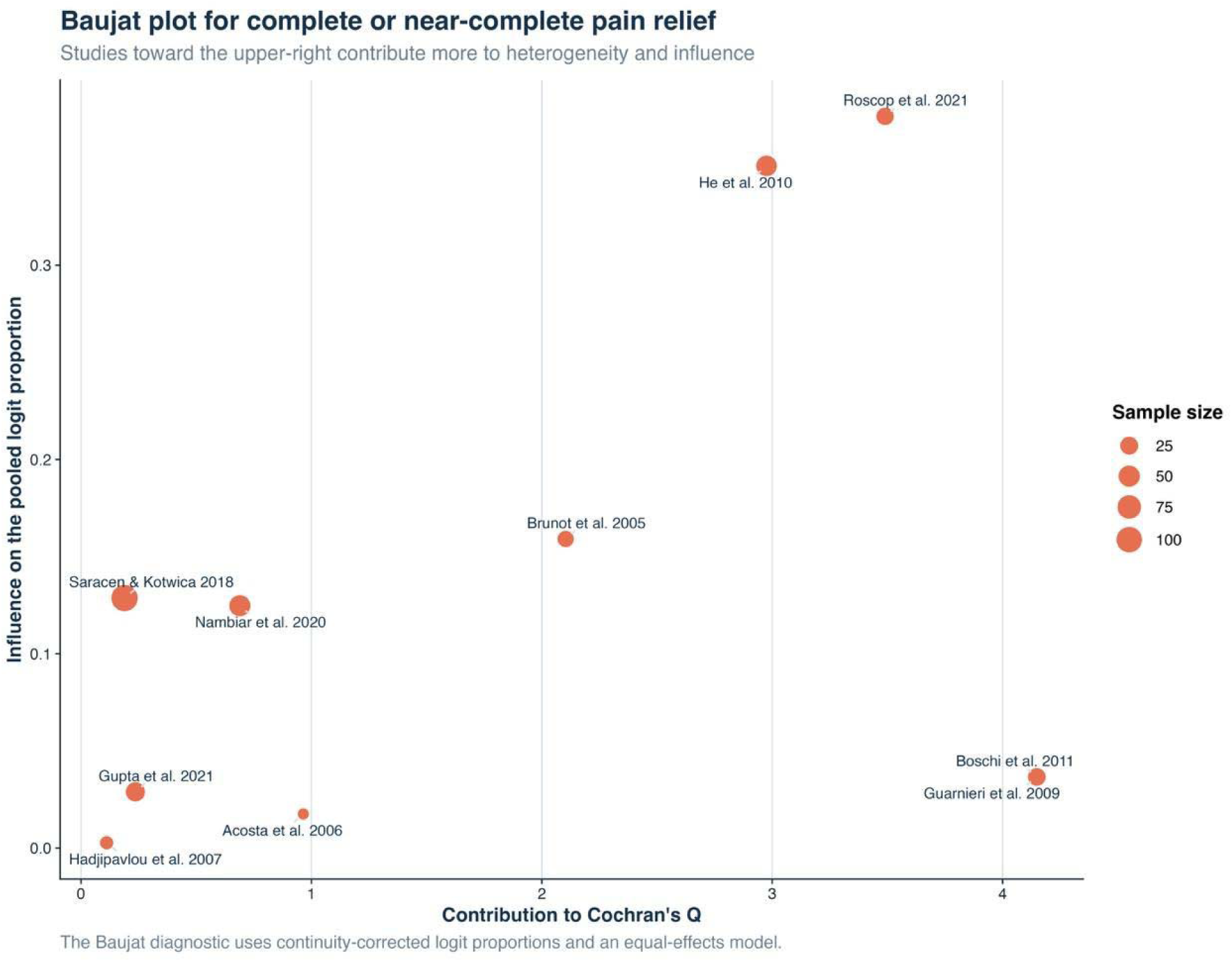
Baujat plot for complete or near-complete pain relief. *Roscop et al. and He et al. contributed most to heterogeneity*.

**Figure S24.**
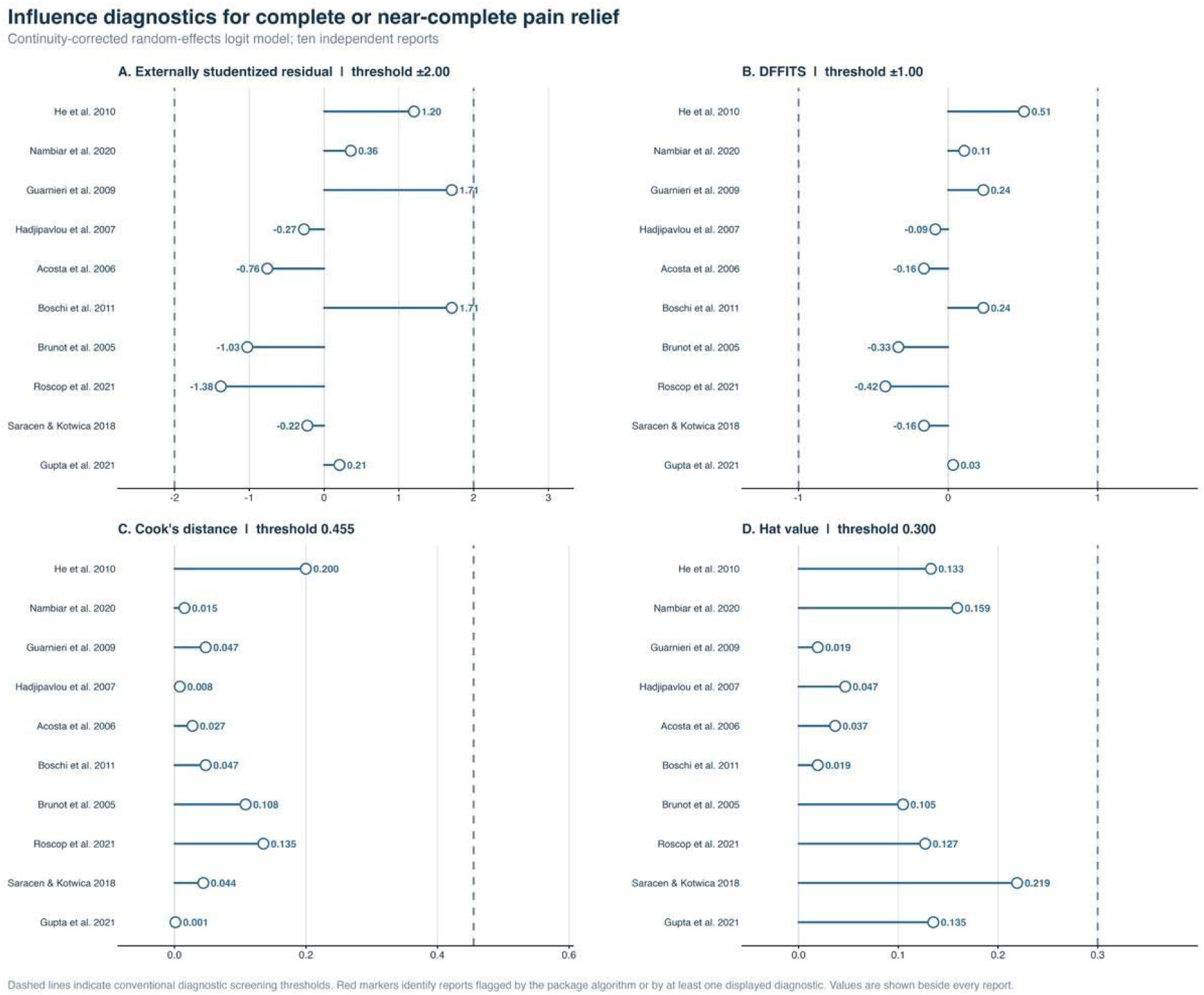
Influence diagnostics for complete or near-complete pain relief. *No study exceeded the prespecified influence thresholds*.

**Figure S25.**
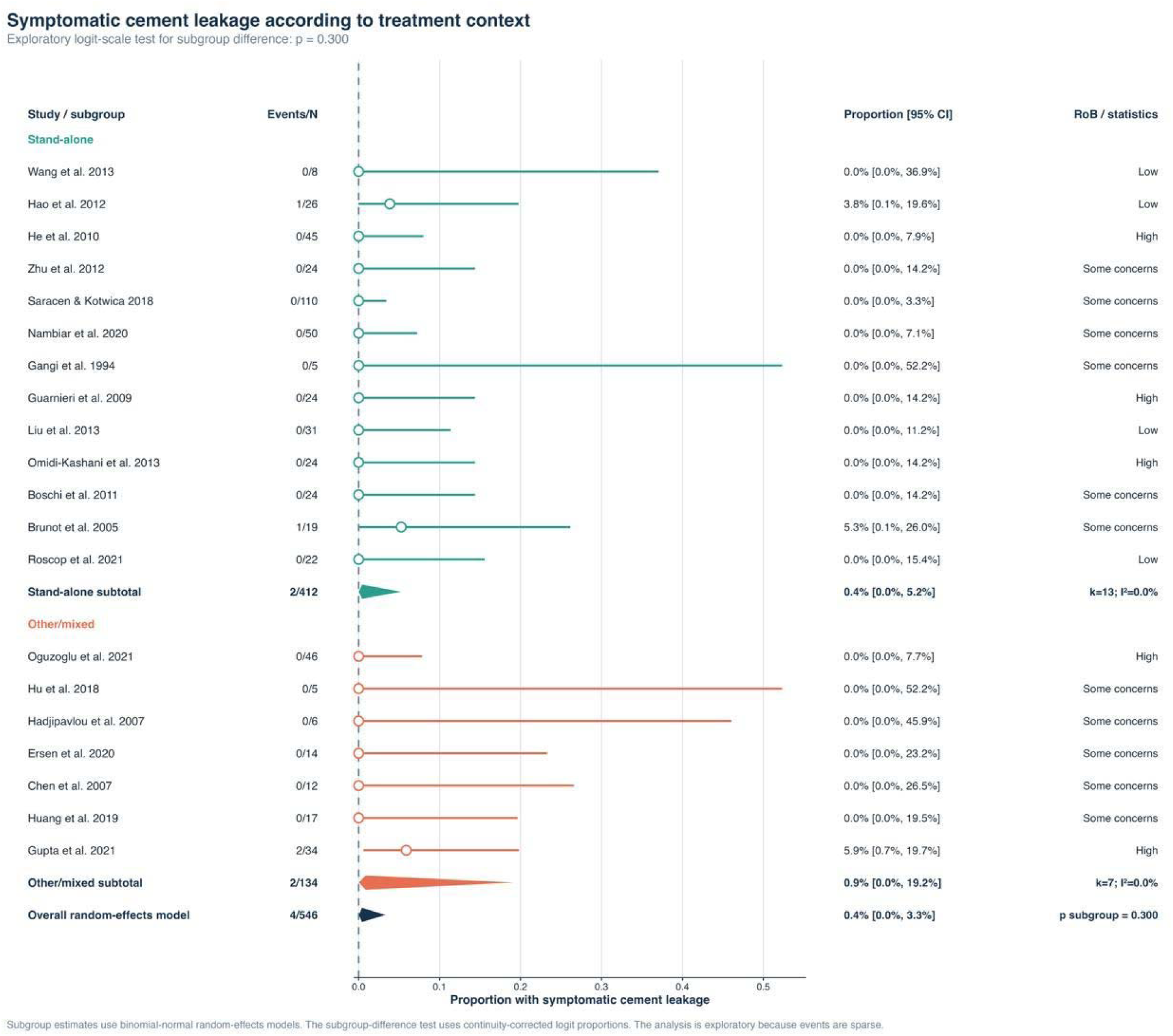
Subgroup analysis of symptomatic cement leakage by treatment context. *Pooled proportions were 0.4% for stand-alone treatment and 0.9% for other or mixed treatment (p=0.300)*.

**Figure S26.**
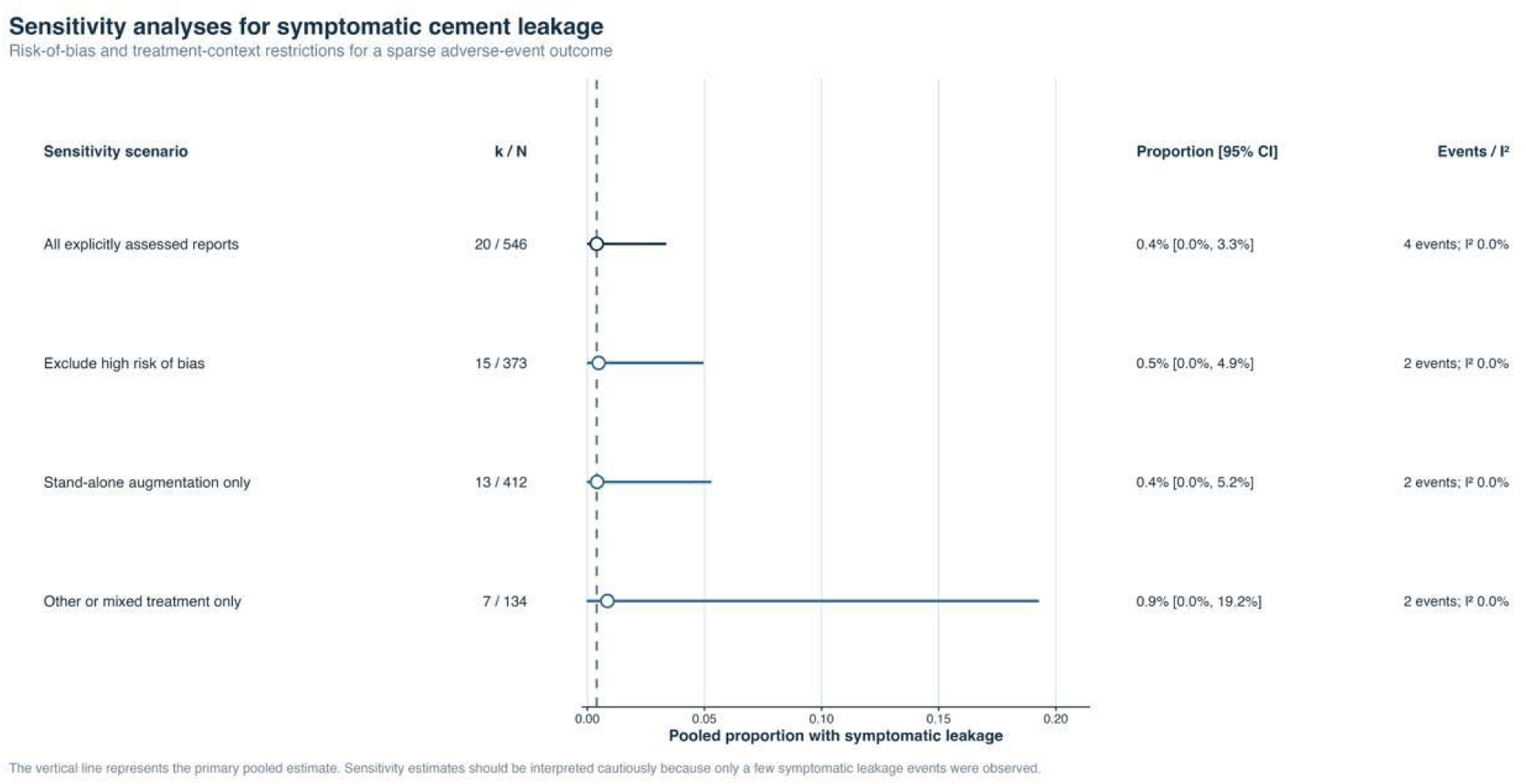
Sensitivity analyses for symptomatic cement leakage. *Estimates ranged from 0.4% to 0.9%. Results were sensitive to the small number of events*.

**Figure S27.**
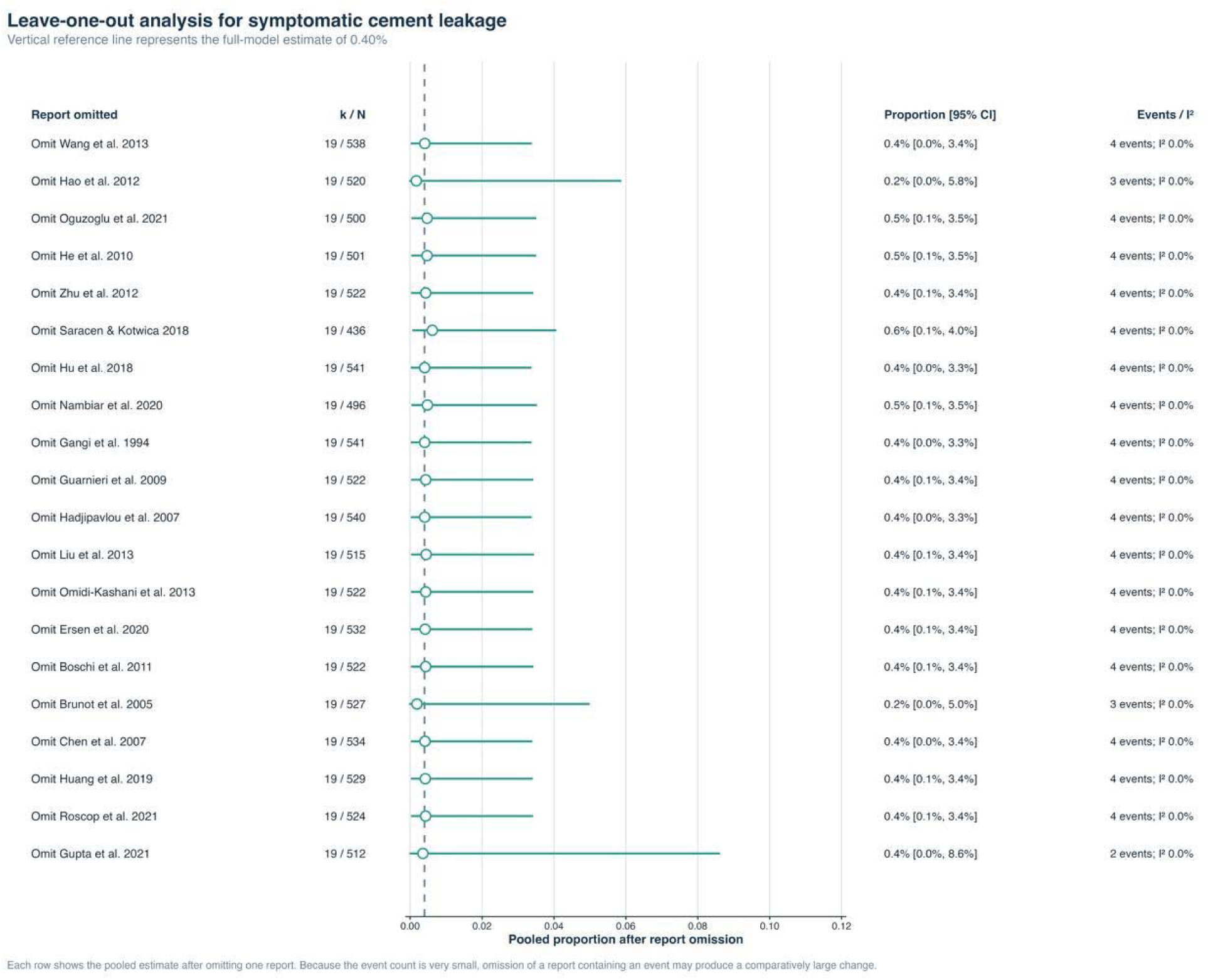
Leave-one-out analysis for symptomatic cement leakage. *Removal of reports containing symptomatic events had the greatest effect, as expected with only four events overall*.

**Figure S28.**
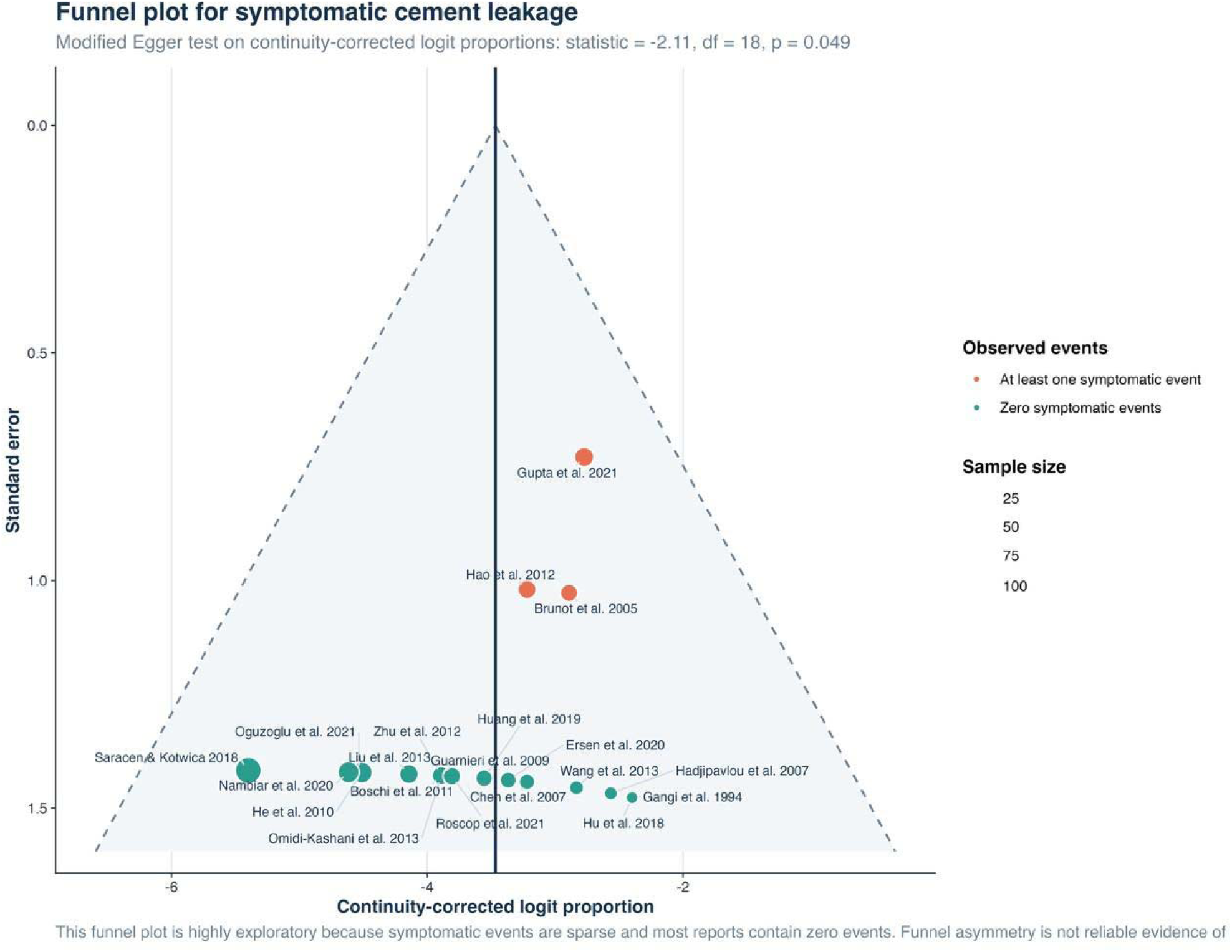
Funnel plot for symptomatic cement leakage. *The plot should be interpreted cautiously because events were sparse*.

**Figure S29.**
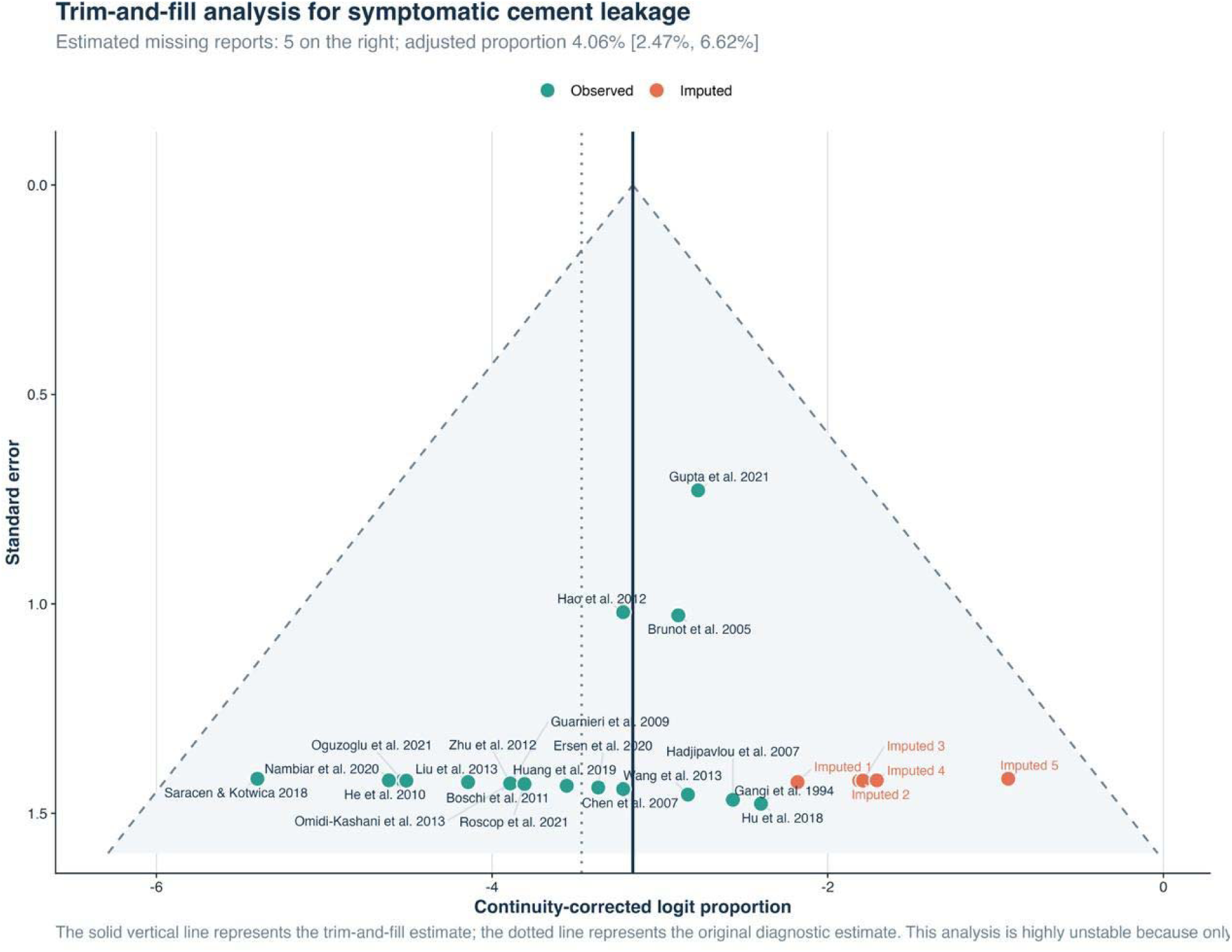
Trim-and-fill analysis for symptomatic cement leakage. *Five studies were imputed, giving an exploratory adjusted estimate of 4.06% (95% CI 2.47-6.62%)*.

**Figure S30.**
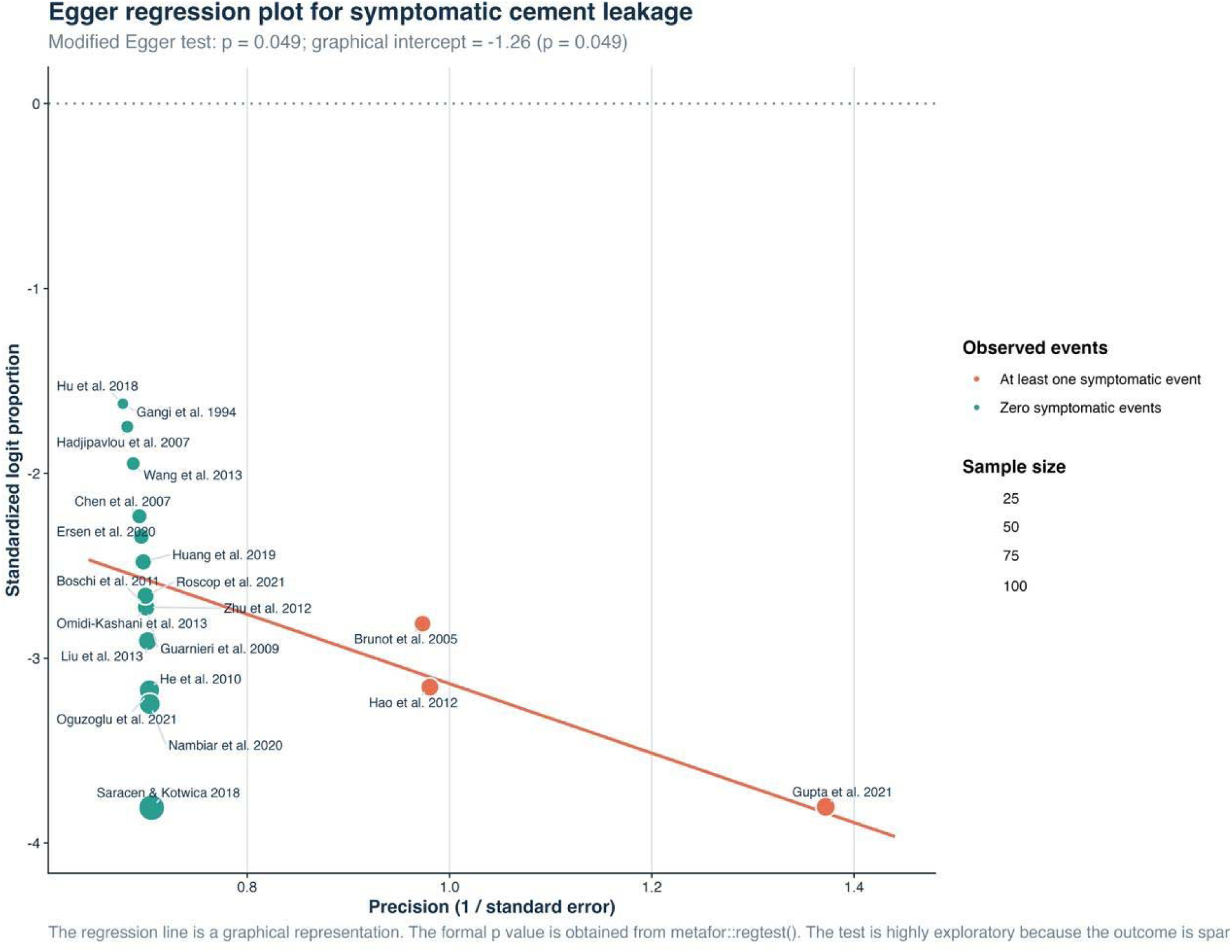
Egger regression for symptomatic cement leakage. *The modified test was nominally significant (p=0.049), although reliability is limited for sparse proportions*.

**Figure S31.**
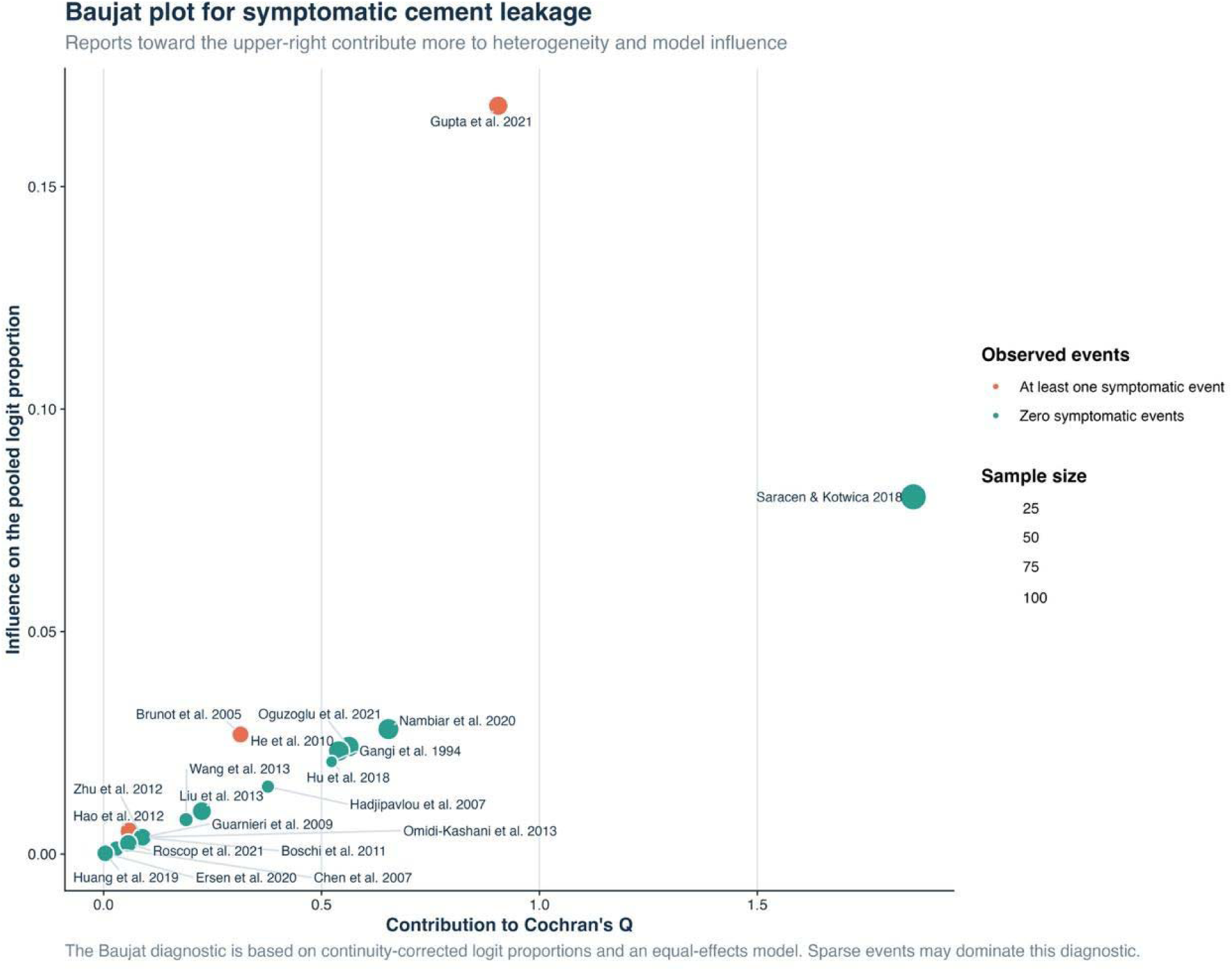
Baujat plot for symptomatic cement leakage. *Gupta et al. was the most influential report*.

**Figure S32.**
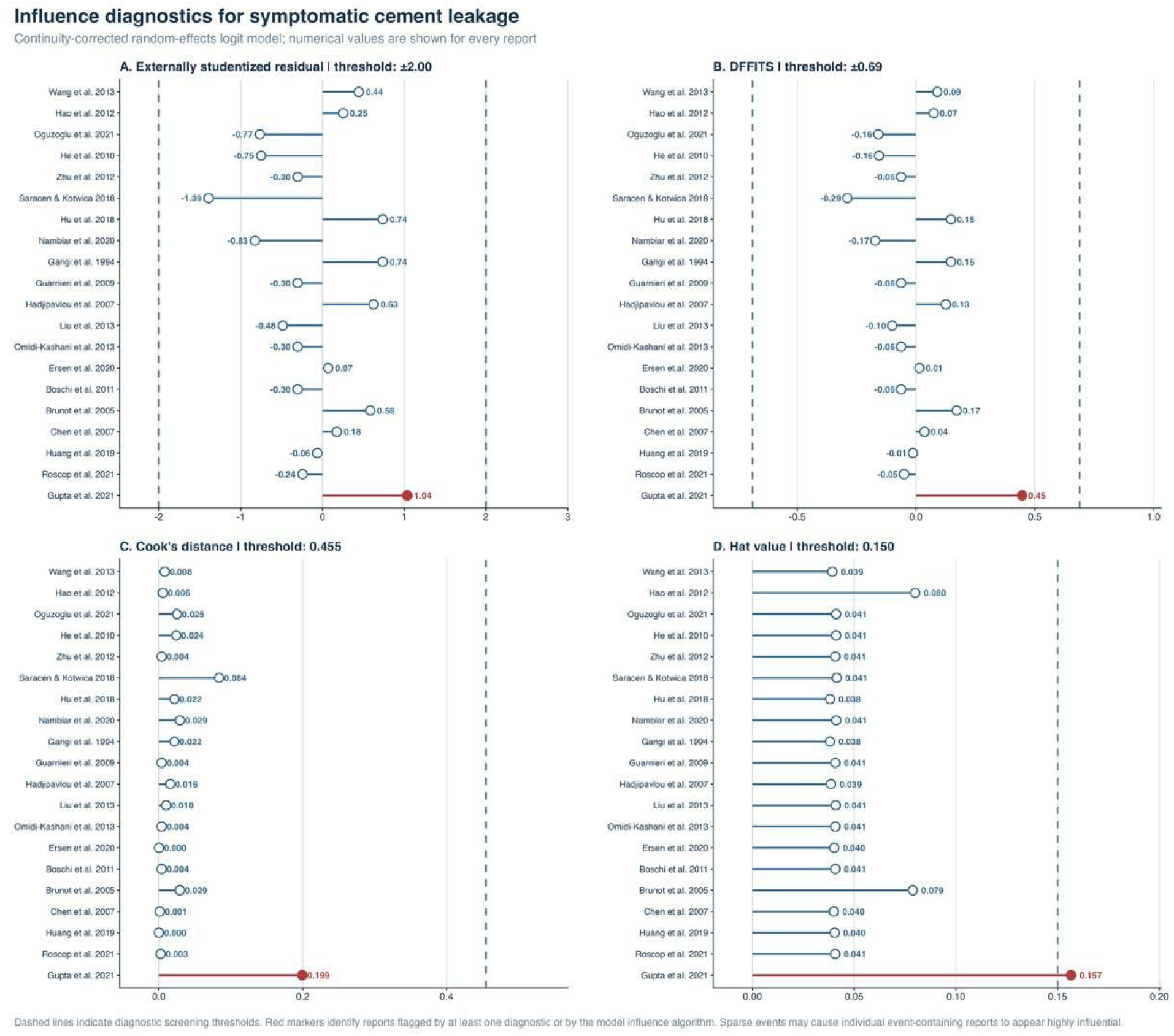
Influence diagnostics for symptomatic cement leakage. *Gupta et al. was flagged by the studentized residual and hat-value diagnostics*.

**Figure S33.**
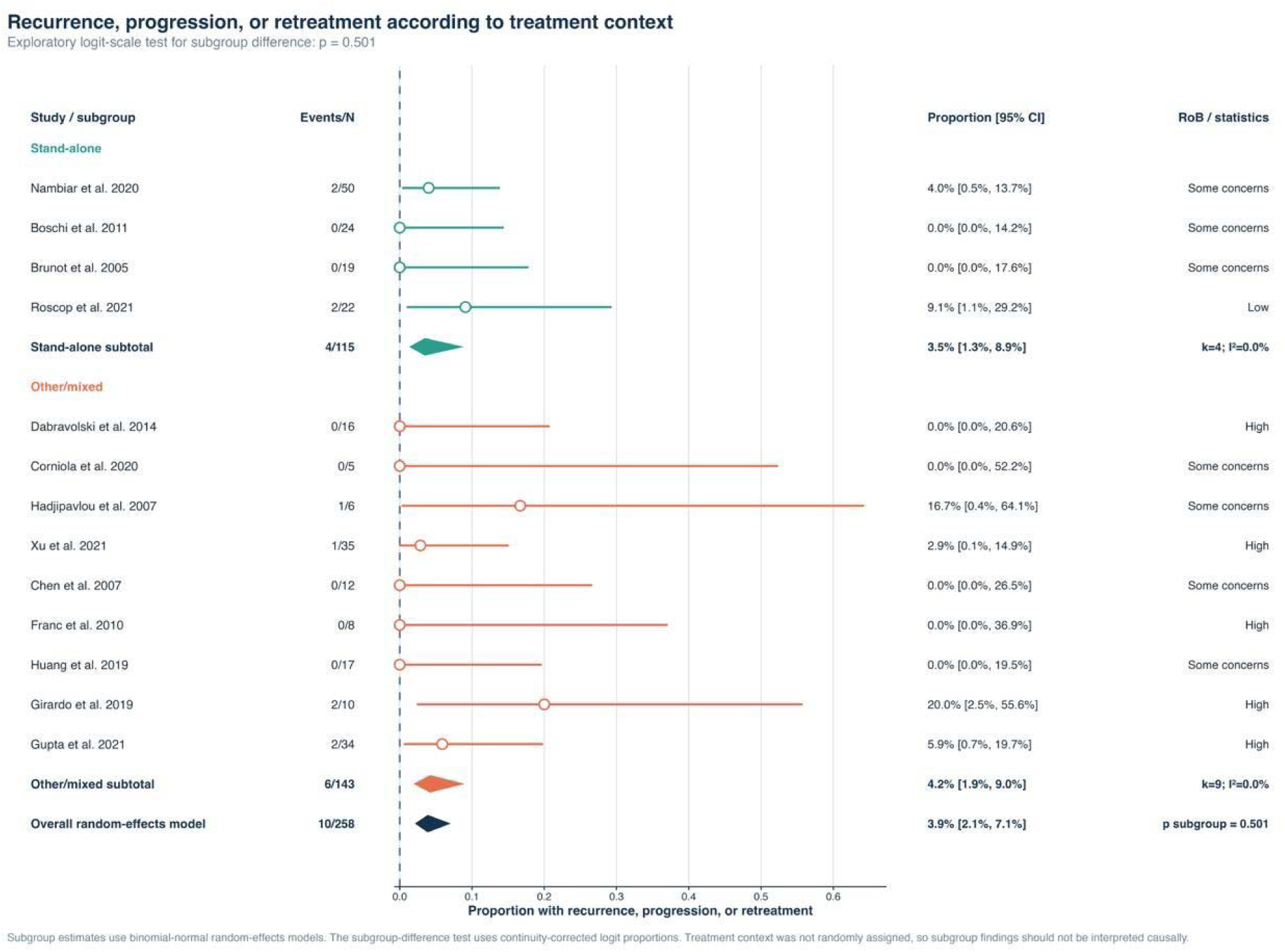
Subgroup analysis of recurrence, progression, or retreatment by treatment context. *Pooled proportions were 3.5% for stand-alone treatment and 4.2% for other or mixed treatment (p=0.501)*.

**Figure S34.**
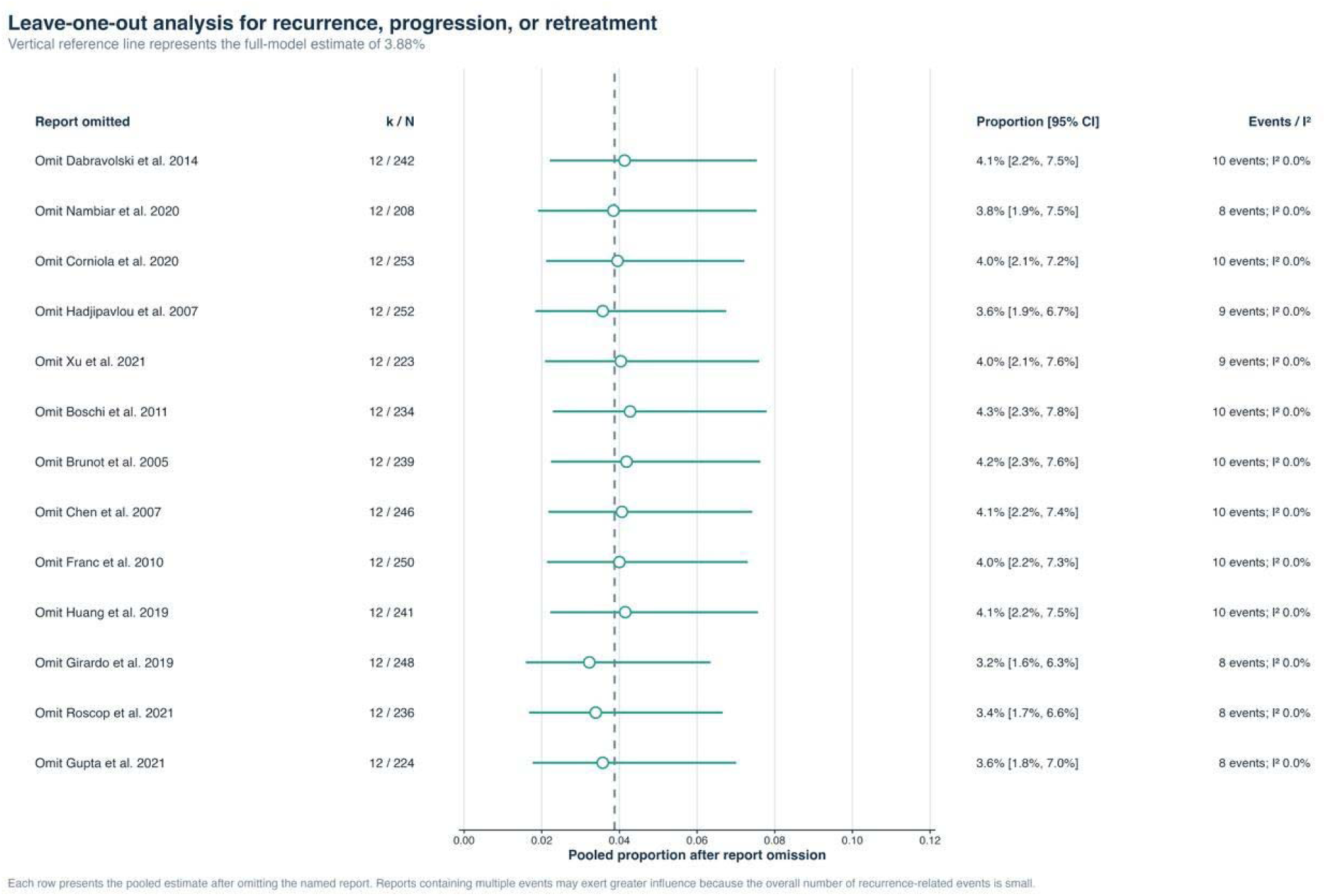
Leave-one-out analysis for recurrence, progression, or retreatment. *Pooled estimates ranged from 3.2% to 4.3%*.

**Figure S35.**
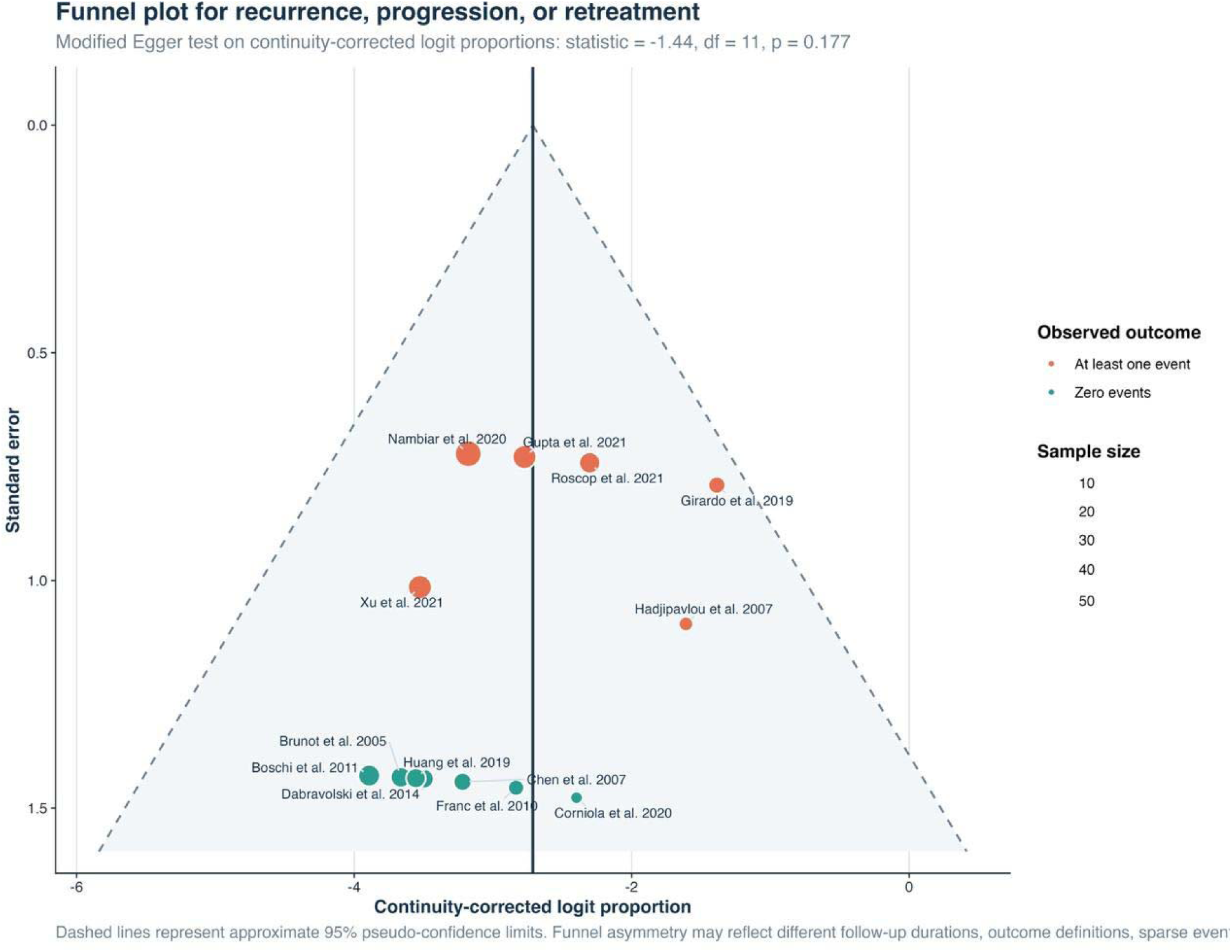
Funnel plot for recurrence, progression, or retreatment. *Egger regression did not indicate significant asymmetry (p=0.177)*.

**Figure S36.**
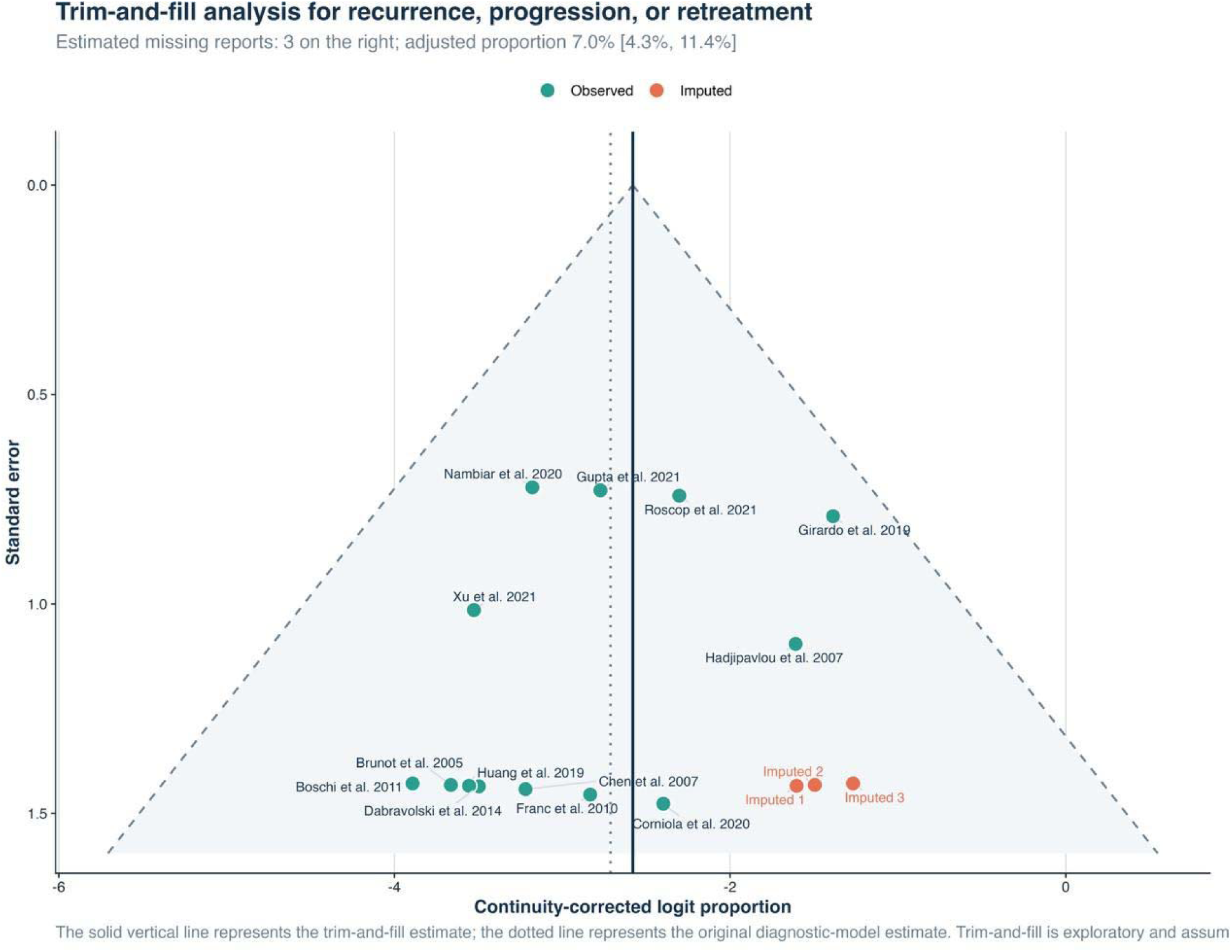
Trim-and-fill analysis for recurrence, progression, or retreatment. *Three studies were imputed, giving an exploratory adjusted estimate of 7.0% (95% CI 4.3-11.4%)*.

**Figure S37.**
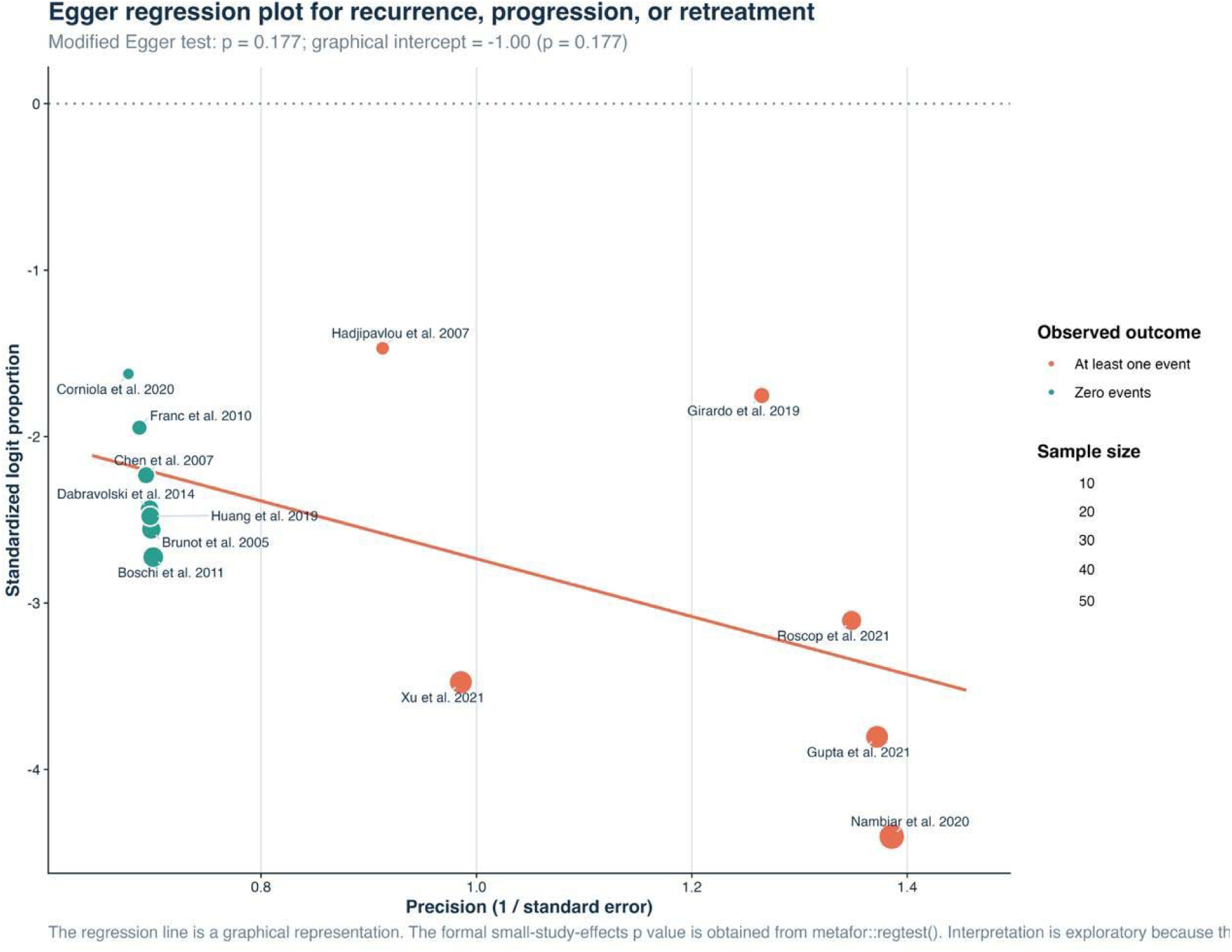
Egger regression for recurrence, progression, or retreatment. *The regression test was not significant (p=0.177)*.

**Figure S38.**
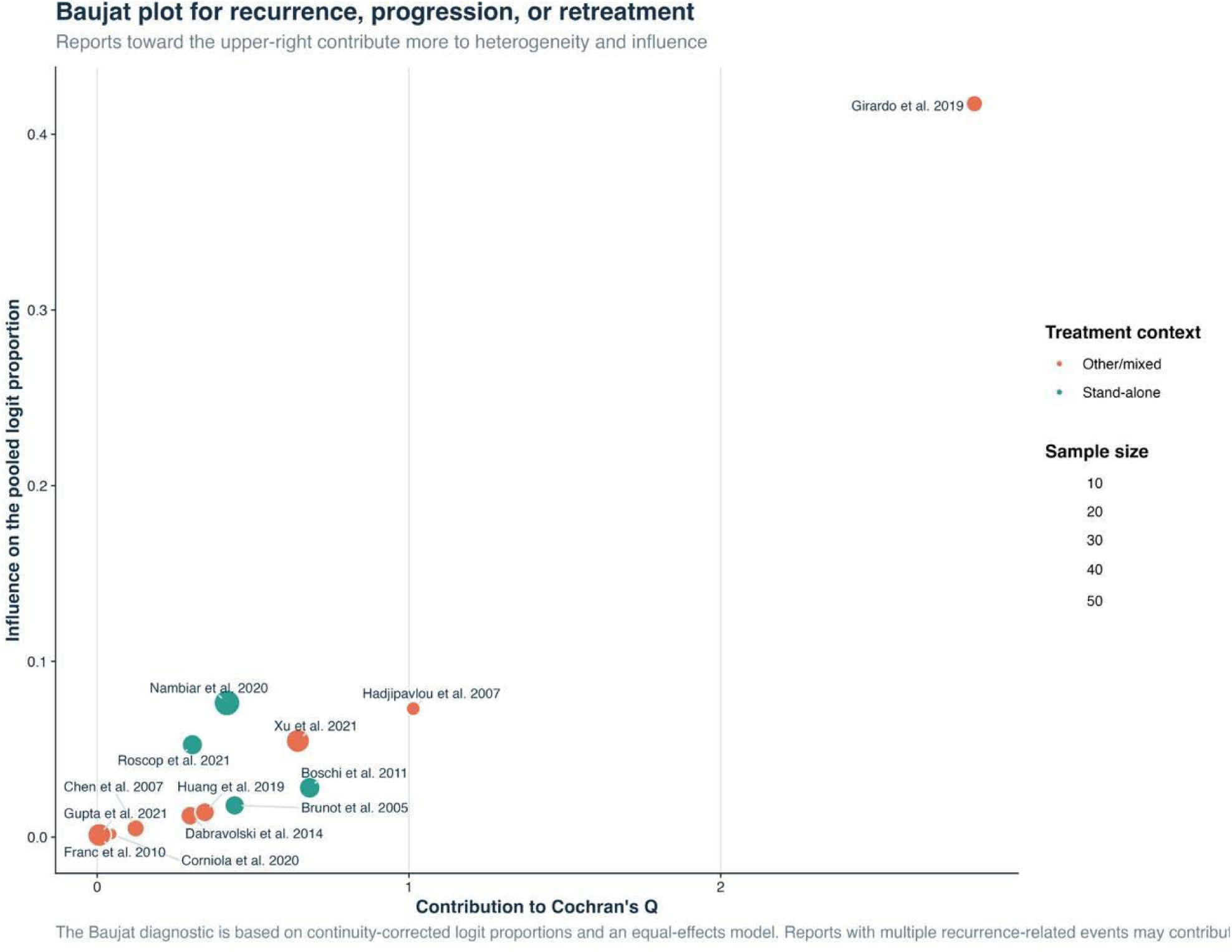
Baujat plot for recurrence, progression, or retreatment. *Girardo et al. contributed most to influence*.

**Figure S39.**
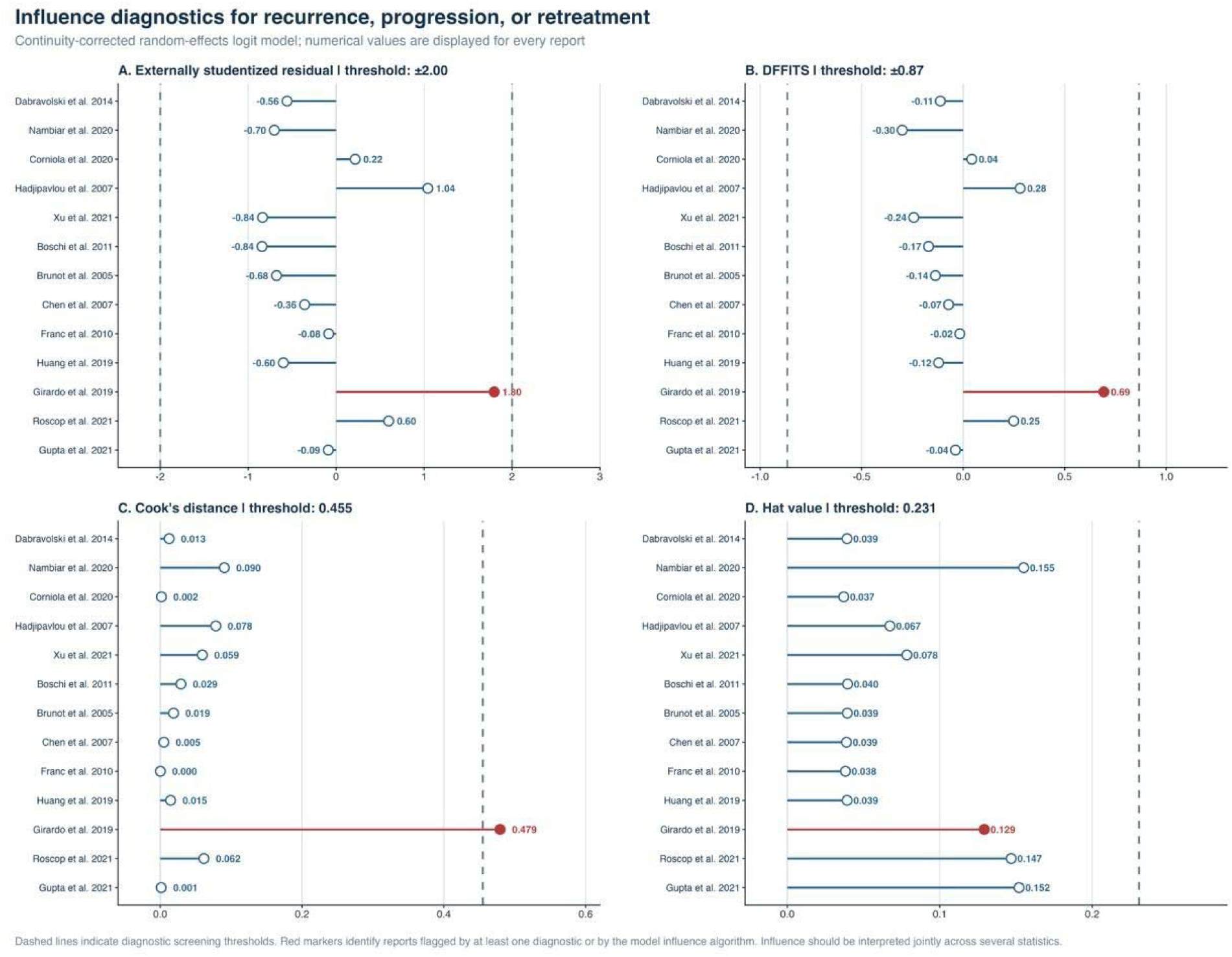
Influence diagnostics for recurrence, progression, or retreatment. *Girardo et al. was the most influential report by studentized residual and Cook’s distance*.

## Notes

### Competing Interest Statement

The authors have declared no competing interest.

