## Supplementary File 2 for "Vertebral Augmentation for Symptomatic Vertebral Hemangiomas: A Systematic Review and Meta-analysis of Pain Relief, Cement Leakage, and Recurrence"

| PUBMED | (  ("primary vertebral hemangioma"[Title/Abstract] OR "primary spinal hemangioma"[Title/Abstract]  OR "hemangioma of vertebra"[Title/Abstract] OR "hemangioma of the spine"[Title/Abstract]  OR "cavernous hemangioma"[Title/Abstract] OR "capillary hemangioma"[Title/Abstract]  OR "benign vertebral vascular tumor"[Title/Abstract] OR "benign spinal vascular tumor"[Title/Abstract]  OR "solitary vertebral hemangioma"[Title/Abstract] OR "solitary spinal hemangioma"[Title/Abstract]  OR "non-metastatic vertebral hemangioma"[Title/Abstract] OR "vertebral hemangioma"[Title/Abstract] OR "spinal hemangioma"[Title/Abstract] OR "vertebral tumor"[Title/Abstract] OR "Hemangioma, Cavernous"[MeSH] OR "Spinal Neoplasms"[MeSH])  )  AND  (  ("vertebroplasty"[Title/Abstract] OR "percutaneous vertebroplasty"[Title/Abstract]  OR "vertebral augmentation"[Title/Abstract] OR "cement augmentation"[Title/Abstract]  OR "bone cement"[Title/Abstract] OR "PMMA"[Title/Abstract] OR "polymethylmethacrylate"[Title/Abstract]  OR "kyphoplasty"[Title/Abstract] OR "balloon kyphoplasty"[Title/Abstract]  OR "percutaneous kyphoplasty"[Title/Abstract])  ) |
| --- | --- |
| SCOPOUS | ( ( "primary vertebral hemangioma" OR "primary spinal hemangioma" OR "hemangioma of vertebra" OR "hemangioma of the spine" OR "cavernous hemangioma" OR "capillary hemangioma" OR "benign vertebral vascular tumor" OR "benign spinal vascular tumor" OR "solitary vertebral hemangioma" OR "solitary spinal hemangioma" OR "non-metastatic vertebral hemangioma" OR "vertebral hemangioma" OR "spinal hemangioma" OR "vertebral tumor" OR "Hemangioma, Cavernous" OR "Spinal Neoplasms" ) ) AND ( ( vertebroplasty OR "percutaneous vertebroplasty" OR "vertebral augmentation" OR "cement augmentation" OR "bone cement" OR PMMA OR polymethylmethacrylate OR kyphoplasty OR "balloon kyphoplasty" OR "percutaneous kyphoplasty" ) ) |
| WOS ADVANCED | (  ((TI="primary vertebral hemangioma" OR AB="primary vertebral hemangioma") OR (TI="primary spinal hemangioma" OR AB="primary spinal hemangioma")  OR (TI="hemangioma of vertebra" OR AB="hemangioma of vertebra") OR (TI="hemangioma of the spine" OR AB="hemangioma of the spine")  OR (TI="cavernous hemangioma" OR AB="cavernous hemangioma") OR (TI="capillary hemangioma" OR AB="capillary hemangioma")  OR (TI="benign vertebral vascular tumor" OR AB="benign vertebral vascular tumor") OR (TI="benign spinal vascular tumor" OR AB="benign spinal vascular tumor")  OR (TI="solitary vertebral hemangioma" OR AB="solitary vertebral hemangioma") OR (TI="solitary spinal hemangioma" OR AB="solitary spinal hemangioma")  OR (TI="non-metastatic vertebral hemangioma" OR AB="non-metastatic vertebral hemangioma") OR (TI="vertebral hemangioma" OR AB="vertebral hemangioma") OR (TI="spinal hemangioma" OR AB="spinal hemangioma") OR (TI="vertebral tumor" OR AB="vertebral tumor") OR ALL="Hemangioma, Cavernous" OR ALL="Spinal Neoplasms")  )  AND  (  ((TI=vertebroplasty OR AB=vertebroplasty) OR (TI="percutaneous vertebroplasty" OR AB="percutaneous vertebroplasty")  OR (TI="vertebral augmentation" OR AB="vertebral augmentation") OR (TI="cement augmentation" OR AB="cement augmentation")  OR (TI="bone cement" OR AB="bone cement") OR (TI=PMMA OR AB=PMMA) OR (TI=polymethylmethacrylate OR AB=polymethylmethacrylate)  OR (TI=kyphoplasty OR AB=kyphoplasty) OR (TI="balloon kyphoplasty" OR AB="balloon kyphoplasty")  OR (TI="percutaneous kyphoplasty" OR AB="percutaneous kyphoplasty"))  ) |
| COCHRANE | Box 1 of search manager  ("primary vertebral hemangioma" OR "primary spinal hemangioma" OR "hemangioma of vertebra" OR "hemangioma of the spine" OR "cavernous hemangioma" OR "capillary hemangioma" OR "benign vertebral vascular tumor" OR "benign spinal vascular tumor" OR "solitary vertebral hemangioma" OR "solitary spinal hemangioma" OR "non-metastatic vertebral hemangioma" OR "vertebral hemangioma" OR "spinal hemangioma" OR "vertebral tumor")  Box 2  (vertebroplasty OR "percutaneous vertebroplasty" OR "vertebral augmentation" OR "cement augmentation" OR "bone cement" OR PMMA OR polymethylmethacrylate OR kyphoplasty OR "balloon kyphoplasty" OR "percutaneous kyphoplasty")  Box 3  (("primary vertebral hemangioma" OR "primary spinal hemangioma" OR "hemangioma of vertebra" OR "hemangioma of the spine" OR "cavernous hemangioma" OR "capillary hemangioma" OR "benign vertebral vascular tumor" OR "benign spinal vascular tumor" OR "solitary vertebral hemangioma" OR "solitary spinal hemangioma" OR "non-metastatic vertebral hemangioma" OR "vertebral hemangioma" OR "spinal hemangioma" OR "vertebral tumor")):ti,ab,kw AND ((vertebroplasty OR "percutaneous vertebroplasty" OR "vertebral augmentation" OR "cement augmentation" OR "bone cement" OR PMMA OR polymethylmethacrylate OR kyphoplasty OR "balloon kyphoplasty" OR "percutaneous kyphoplasty")):ti,ab,kw |
| EMBASE | ( ('primary vertebral hemangioma':ti,ab OR 'primary spinal hemangioma':ti,ab OR 'hemangioma of vertebra':ti,ab OR 'hemangioma of the spine':ti,ab OR 'cavernous hemangioma':ti,ab OR 'capillary hemangioma':ti,ab OR 'benign vertebral vascular tumor':ti,ab OR 'benign spinal vascular tumor':ti,ab OR 'solitary vertebral hemangioma':ti,ab OR 'solitary spinal hemangioma':ti,ab OR 'non-metastatic vertebral hemangioma':ti,ab OR 'vertebral hemangioma':ti,ab OR 'spinal hemangioma':ti,ab OR 'vertebral tumor':ti,ab OR 'Hemangioma, Cavernous'/exp OR 'Spinal Neoplasms'/exp) ) AND ( (vertebroplasty:ti,ab OR 'percutaneous vertebroplasty':ti,ab OR 'vertebral augmentation':ti,ab OR 'cement augmentation':ti,ab OR 'bone cement':ti,ab OR PMMA:ti,ab OR polymethylmethacrylate:ti,ab OR kyphoplasty:ti,ab OR 'balloon kyphoplasty':ti,ab OR 'percutaneous kyphoplasty':ti,ab) ) |
