## Supplementary File 5 for "Vertebral Augmentation for Symptomatic Vertebral Hemangiomas: A Systematic Review and Meta-analysis of Pain Relief, Cement Leakage, and Recurrence": Acosta 2006.docx

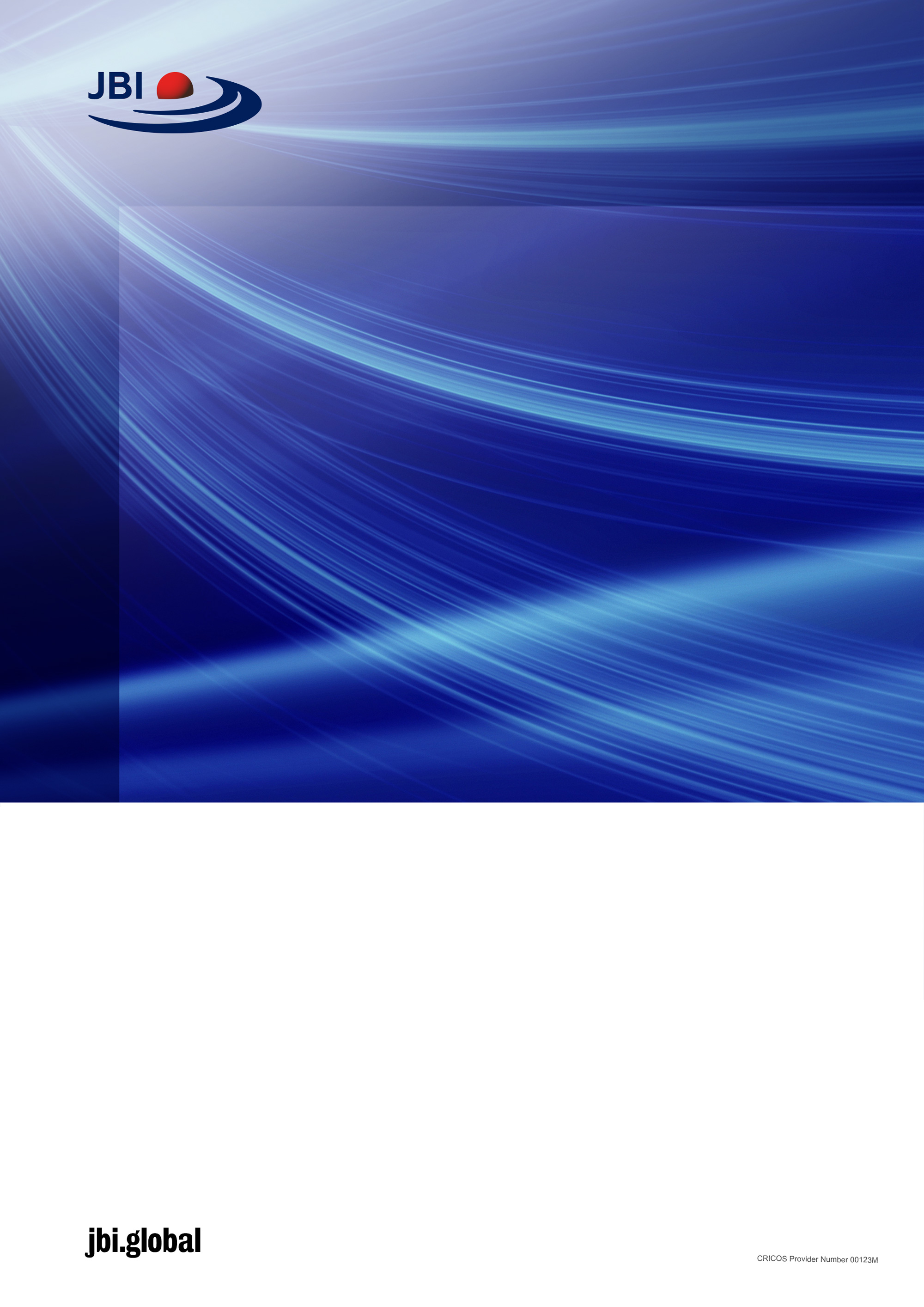

Checklist for case series

Critical Appraisal tools for use in JBI Systematic Reviews

Introduction

JBI is an international research organisation based in the Faculty of Health and Medical Sciences at the University of Adelaide, South Australia. JBI develops and delivers unique evidence-based information, software, education and training designed to improve healthcare practice and health outcomes. With over 70 Collaborating Entities, servicing over 90 countries, JBI is a recognised global leader in evidence-based healthcare.

### JBI Systematic Reviews

The core of evidence synthesis is the systematic review of literature of a particular intervention, condition or issue. The systematic review is essentially an analysis of the available literature (that is, evidence) and a judgment of the effectiveness or otherwise of a practice, involving a series of complex steps. JBI takes a particular view on what counts as evidence and the methods utilised to synthesise those different types of evidence. In line with this broader view of evidence, JBI has developed theories, methodologies and rigorous processes for the critical appraisal and synthesis of these diverse forms of evidence in order to aid in clinical decision-making in healthcare. There now exists JBI guidance for conducting reviews of effectiveness research, qualitative research, prevalence/incidence, etiology/risk, economic evaluations, text/opinion, diagnostic test accuracy, mixed-methods, umbrella reviews and scoping reviews. Further information regarding JBI systematic reviews can be found in the [JBI Evidence Synthesis Manual](https://jbi-global-wiki.refined.site/space/MANUAL).

### JBI Critical Appraisal Tools

All systematic reviews incorporate a process of critique or appraisal of the research evidence. The purpose of this appraisal is to assess the methodological quality of a study and to determine the extent to which a study has addressed the possibility of bias in its design, conduct and analysis. All papers selected for inclusion in the systematic review (that is – those that meet the inclusion criteria described in the protocol) need to be subjected to rigorous appraisal by two critical appraisers. The results of this appraisal can then be used to inform synthesis and interpretation of the results of the study. JBI Critical appraisal tools have been developed by the JBI and collaborators and approved by the JBI Scientific Committee following extensive peer review. Although designed for use in systematic reviews, JBI critical appraisal tools can also be used when creating Critically Appraised Topics (CAT), in journal clubs and as an educational tool.

**JBI Critical Appraisal Checklist for Case Series**

Reviewer	___________Farzin MohammadMoradi_________ Date___________14^th^ July 2026_______

Author_______ Acosta	Year____2006_____ Record Number_____22____

|  | Yes | No | Unclear | Not applicable |
| --- | --- | --- | --- | --- |
| - Were there clear criteria for inclusion in the case series? | □ | □ | □ | □ |
| - Was the condition measured in a standard, reliable way for all participants included in the case series? | □ | □ | □ | □ |
| - Were valid methods used for identification of the condition for all participants included in the case series? | □ | □ | □ | □ |
| - Did the case series have consecutive inclusion of participants? | □ | □ | □ | □ |
| - Did the case series have complete inclusion of participants? | □ | □ | □ | □ |
| - Was there clear reporting of the demographics of the participants in the study? | □ | □ | □ | □ |
| - Was there clear reporting of clinical information of the participants? | □ | □ | □ | □ |
| - Were the outcomes or follow up results of cases clearly reported? | □ | □ | □ | □ |
| - Was there clear reporting of the presenting site(s)/clinic(s) demographic information? | □ | □ | □ | □ |
| - Was statistical analysis appropriate? | □ | □ | □ | □ |

Overall appraisal:	Include	□	Exclude	□	Seek further info □

Comments (Including reason for exclusion)

____________LOW RISK (70%)

_____________________________________________________________________________________________________________________________________________

Introduction to the Case Series
Critical Appraisal Tool

*How to cite:* Munn Z, Barker T, Moola S, Tufanaru C, Stern C, McArthur A, Stephenson M, Aromataris E. Methodological quality of case series studies, JBI Evidence Synthesis, doi: 10.11124/JBISRIR-D-19-00099

The definition of a case series varies across the medical literature, which has resulted in inconsistent use of this term (Appendix 1).^1-3^ The gamut of case studies is wide, with some studies claiming to be a case series realistically being nothing more than a collection of case reports, with others more akin to cohort studies or even quasi-experimental before and after studies. This has created difficulty in assigning ‘case series’ a position in the hierarchy of evidence and identifying and appropriate critical appraisal tool.^1, 2^

Dekkers et al. define a case series as a study in which ‘only patients with the outcome are sampled (either those who have an exposure or those who are selected without regard to exposure), which does not permit calculation of an absolute risk.’^1p.39^ The outcome could be a disease or a disease related outcome. This is contrasted to cohort studies where sampling is based on exposure (or characteristic), and case- control studies where there is a comparison group without the disease.

The completeness of a case series contributes to its reliability.^1^ Studies that indicate a consecutive and complete inclusion are more reliable than those that do not. For example, a case series that states ‘we included all patients (24) with osteosarcoma who presented to our clinic between March 2005 and June 2006’ is more reliable than a study that simply states ‘we report a case series of 24 people with osteosarcoma.’

For the purposes of this checklist, we agree with the principles outlined in the Dekker et al. paper, and define case series as studies where only patients with a certain disease or disease-related outcome are sampled. Some of the items below relate to risk of bias, whilst others relate to ensuring adequate reporting and statistical analysis. A response of ‘no’ to any of the questions below negatively impacts the quality of a case series.

Tool Guidance

Answers: Yes, No, Unclear or Not/Applicable

1. **Were there clear criteria for inclusion in the case series?**

The authors should provide clear inclusion (and exclusion criteria where appropriate) for the study participants. The inclusion/exclusion criteria should be specified (e.g., risk, stage of disease progression) with sufficient detail and all the necessary information critical to the study.

### 2. Was the condition measured in a standard, reliable way for all participants included in the case series?

The study should clearly describe the method of measurement of the condition. This should be done in a standard (i.e. same way for all patients) and reliable (i.e. repeatable and reproducible results) way.

### 3. Were valid methods used for identification of the condition for all participants included in the case series?

Many health problems are not easily diagnosed or defined and some measures may not be capable of including or excluding appropriate levels or stages of the health problem. If the outcomes were assessed based on existing definitions or diagnostic criteria, then the answer to this question is likely to be yes. If the outcomes were assessed using observer reported, or self-reported scales, the risk of over- or under-reporting is increased, and objectivity is compromised. Importantly, determine if the measurement tools used were validated instruments as this has a significant impact on outcome assessment validity.

### 4. Did the case series have consecutive inclusion of participants?

Studies that indicate a consecutive inclusion are more reliable than those that do not. For example, a case series that states ‘we included all patients (24) with osteosarcoma who presented to our clinic between March 2005 and June 2006’ is more reliable than a study that simply states ‘we report a case series of 24 people with osteosarcoma.’

### 5. Did the case series have complete inclusion of participants?

The completeness of a case series contributes to its reliability (1). Studies that indicate a complete inclusion are more reliable than those that do not. A stated above, a case series that states ‘we included all patients (24) with osteosarcoma who presented to our clinic between March 2005 and June 2006’ is more reliable than a study that simply states ‘we report a case series of 24 people with osteosarcoma.’

### 6. Was there clear reporting of the demographics of the participants in the study?

The case series should clearly describe relevant participant’s demographics such as the following information where relevant: participant’s age, sex, education, geographic region, ethnicity, time period, education.

### 7. Was there clear reporting of clinical information of the participants?

There should be clear reporting of clinical information of the participants such as the following information where relevant: disease status, comorbidities, stage of disease, previous interventions/treatment, results of diagnostic tests, etc.

### 8. Were the outcomes or follow-up results of cases clearly reported?

The results of any intervention or treatment should be clearly reported in the case series. A good case study should clearly describe the clinical condition post-intervention in terms of the presence or lack of symptoms. The outcomes of management/treatment when presented as images or figures can help in conveying the information to the reader/clinician. It is important that adverse events are clearly documented and described, particularly a new or unique condition is being treated or when a new drug or treatment is used. In addition, unanticipated events, if any that may yield new or useful information should be identified and clearly described.

### 9. Was there clear reporting of the presenting site(s)/clinic(s) demographic information?

Certain diseases or conditions vary in prevalence across different geographic regions and populations (e.g. women vs. men, sociodemographic variables between countries). The study sample should be described in sufficient detail so that other researchers can determine if it is comparable to the population of interest to them.

### 10.  Was statistical analysis appropriate?

As with any consideration of statistical analysis, consideration should be given to whether there was a more appropriate alternate statistical method that could have been used. The methods section of studies should be detailed enough for reviewers to identify which analytical techniques were used and whether these were suitable.

### Appendix 1: Case series definitions:

‘A report on a series of patients with an outcome of interest. No control group is involved.’(4) ^(p 279)^

‘A case series is a descriptive study involving a group of patients who all have the same disease or condition: the aim is to describe common and differing characteristics of a particular group of individuals’ (Oxford Handbook of medical statistics)

‘A group or series of case reports involving patients who were given similar treatment. Reports of case series usually contain detailed information about the individual patients. This includes demographic information (for example, age, gender, ethnic origin) and information on diagnosis, treatment, response to treatment, and follow-up after treatment.’ Law K, Howick J. OCEBM Table of Evidence Glossary. 2013 [cited 2014 10th January]; Available from: <http://www.cebm.net/index.aspx?o=1116>

‘A **case series** (also known as a clinical **series**) is a type of medical research study that tracks subjects with a known exposure, such as patients who have received a similar treatment, or examines their medical records for exposure and outcome.’ Wikipedia

‘A study which makes observations on a series of individuals, usually all receiving the same intervention, with no control group. Comments: At this stage it is unclear whether case series should be included in Cochrane systematic reviews, but we have left them in the list so that working groups can consider whether there are circumstances in which it would be appropriate to include them, and to assess risk of bias. A particular reason for including case series might be where they provide evidence relating to adverse effects of an intervention. Potential examples of risk of bias might be that if a case series does not [attempt to] recruit consecutive participants, this might introduce a risk of selection bias, while some case series could be at risk of detection bias, if the circumstances in which adverse effects are reported (or elicited) are not standardised.’ <http://bmg.cochrane.org/research-projectscochrane-risk-bias-tool>
