## Supplementary File 5 for "Vertebral Augmentation for Symptomatic Vertebral Hemangiomas: A Systematic Review and Meta-analysis of Pain Relief, Cement Leakage, and Recurrence": Anselmetti 2011.docx

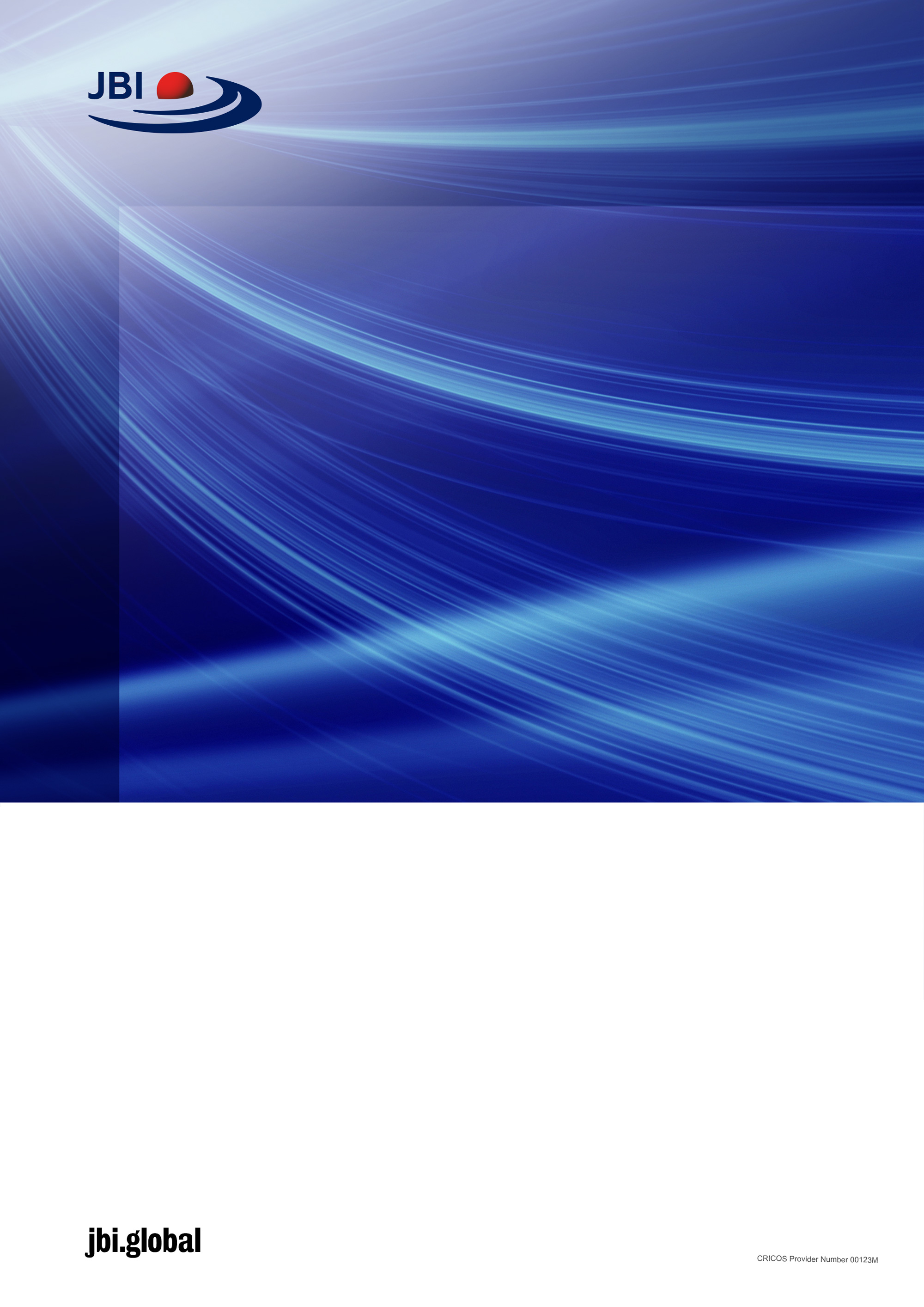

Checklist for case series

Critical Appraisal tools for use in JBI Systematic Reviews

Introduction

JBI is an international research organisation based in the Faculty of Health and Medical Sciences at the University of Adelaide, South Australia. JBI develops and delivers unique evidence-based information, software, education and training designed to improve healthcare practice and health outcomes. With over 70 Collaborating Entities, servicing over 90 countries, JBI is a recognised global leader in evidence-based healthcare.
