## Supplementary File 5 for "Vertebral Augmentation for Symptomatic Vertebral Hemangiomas: A Systematic Review and Meta-analysis of Pain Relief, Cement Leakage, and Recurrence": Byval'tsev 2015.docx

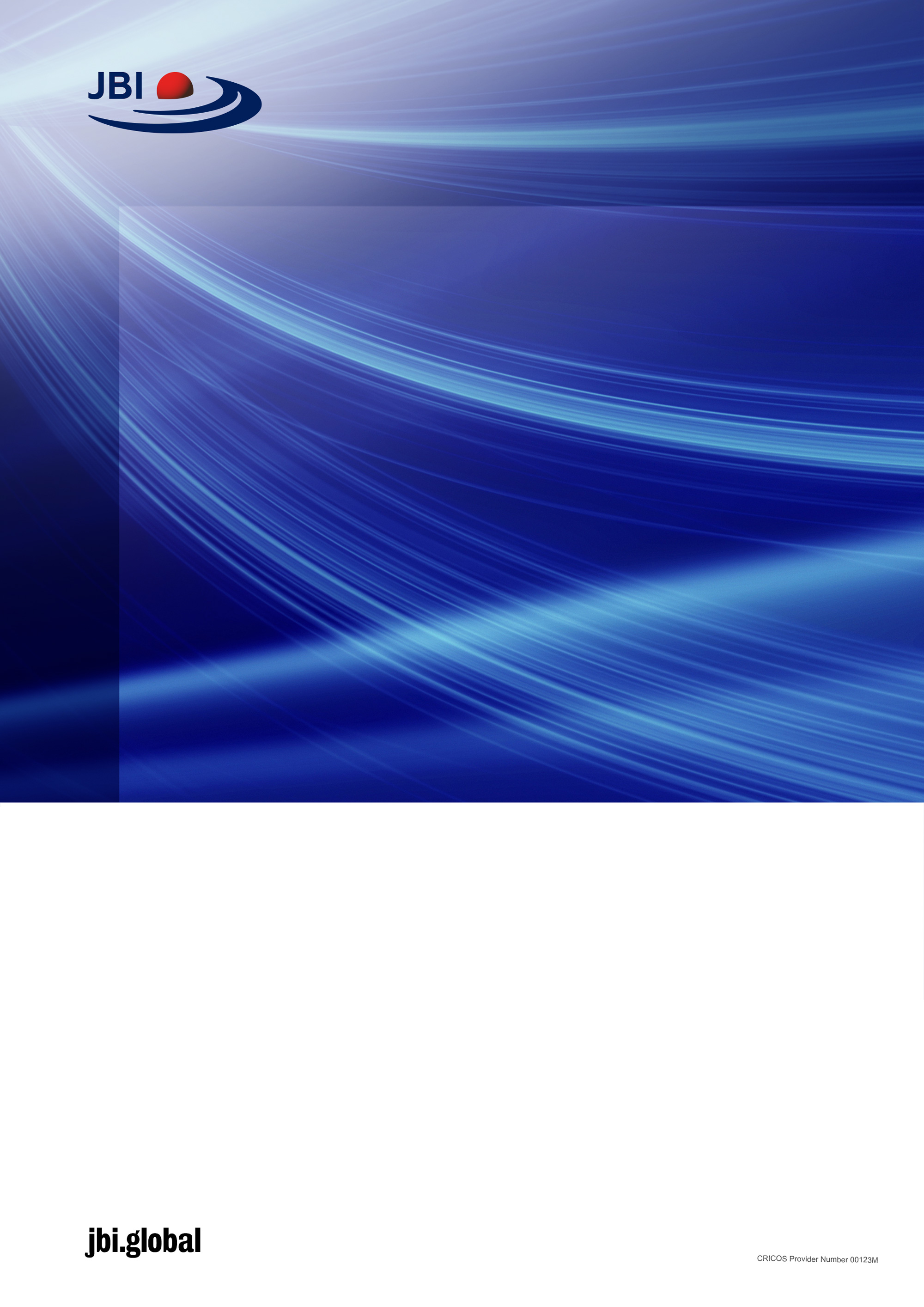

checklist for

cohort studies

Critical Appraisal tools for use in JBI Systematic Reviews

Introduction

JBI is an international research organisation based in the Faculty of Health and Medical Sciences at the University of Adelaide, South Australia. JBI develops and delivers unique evidence-based information, software, education and training designed to improve healthcare practice and health outcomes. With over 70 Collaborating Entities, servicing over 90 countries, JBI is a recognised global leader in evidence-based healthcare.

### JBI Systematic Reviews

The core of evidence synthesis is the systematic review of literature of a particular intervention, condition or issue. The systematic review is essentially an analysis of the available literature (that is, evidence) and a judgment of the effectiveness or otherwise of a practice, involving a series of complex steps. JBI takes a particular view on what counts as evidence and the methods utilised to synthesise those different types of evidence. In line with this broader view of evidence, JBI has developed theories, methodologies and rigorous processes for the critical appraisal and synthesis of these diverse forms of evidence in order to aid in clinical decision-making in healthcare. There now exists JBI guidance for conducting reviews of effectiveness research, qualitative research, prevalence/incidence, etiology/risk, economic evaluations, text/opinion, diagnostic test accuracy, mixed-methods, umbrella reviews and scoping reviews. Further information regarding JBI systematic reviews can be found in the [JBI Evidence Synthesis Manual](https://jbi-global-wiki.refined.site/space/MANUAL).

### JBI Critical Appraisal Tools

All systematic reviews incorporate a process of critique or appraisal of the research evidence. The purpose of this appraisal is to assess the methodological quality of a study and to determine the extent to which a study has addressed the possibility of bias in its design, conduct and analysis. All papers selected for inclusion in the systematic review (that is – those that meet the inclusion criteria described in the protocol) need to be subjected to rigorous appraisal by two critical appraisers. The results of this appraisal can then be used to inform synthesis and interpretation of the results of the study. JBI Critical appraisal tools have been developed by the JBI and collaborators and approved by the JBI Scientific Committee following extensive peer review. Although designed for use in systematic reviews, JBI critical appraisal tools can also be used when creating Critically Appraised Topics (CAT), in journal clubs and as an educational tool.

JBI Critical Appraisal Checklist for cohort studies

Reviewer	__Farzin MohammadMoradi_______________________________ Date______________14^th^ July_________________

Author____________________ Byval'tsev	Year____2015_____ Record Number____10_____

|  | Yes | No | Unclear | Not applicable |
| --- | --- | --- | --- | --- |
| 1. Were the two groups similar and recruited from the same population? | □ | □ | □ | □ |
| 2. Were the exposures measured similarly to assign people to both exposed and unexposed groups? | □ | □ | □ | □ |
| 3. Was the exposure measured in a valid and reliable way? | □ | □ | □ | □ |
| 4. Were confounding factors identified? | □ | □ | □ | □ |
| 5. Were strategies to deal with confounding factors stated? | □ | □ | □ | □ |
| 6. Were the groups/participants free of the outcome at the start of the study (or at the moment of exposure)? | □ | □ | □ | □ |
| 7. Were the outcomes measured in a valid and reliable way? | □ | □ | □ | □ |
| 8. Was the follow up time reported and sufficient to be long enough for outcomes to occur? | □ | □ | □ | □ |
| 9. Was follow up complete, and if not, were the reasons to loss to follow up described and explored? | □ | □ | □ | □ |
| 10. Were strategies to address incomplete follow up utilized? | □ | □ | □ | □ |
| 11. Was appropriate statistical analysis used? | □ | □ | □ | □ |

Overall appraisal:	Include	□	Exclude	□	Seek further info □

Comments (Including reason for exclusion) LOW RISK (90%)

________________________________________

Explanation of cohort studies critical appraisal

How to Cite: *Moola S, Munn Z, Tufanaru C, Aromataris E, Sears K, Sfetcu R, Currie M, Qureshi R, Mattis P, Lisy K, Mu P-F. Chapter 7: Systematic reviews of etiology and risk . In: Aromataris E, Munn Z (Editors)*. JBI Manual for Evidence Synthesis. JBI, 2020. Available from <https://synthesismanual.jbi.global>

### Cohort Studies Critical Appraisal Tool

Answers: Yes, No, Unclear or Not/Applicable

### 1. Were the two groups similar and recruited from the same population?

Check the paper carefully for descriptions of participants to determine if patients within and across groups have similar characteristics in relation to exposure (e.g. risk factor under investigation). The two groups selected for comparison should be as similar as possible in all characteristics except for their exposure status, relevant to the study in question. The authors should provide clear inclusion and exclusion criteria that they developed prior to recruitment of the study participants.

### 2. Were the exposures measured similarly to assign people to both exposed and unexposed groups?

A high quality study at the level of cohort design should mention or describe how the exposures were measured. The exposure measures should be clearly defined and described in detail. This will enable reviewers to assess whether or not the participants received the exposure of interest.

### 3. Was the exposure measured in a valid and reliable way?

The study should clearly describe the method of measurement of exposure. Assessing validity requires that a 'gold standard' is available to which the measure can be compared. The validity of exposure measurement usually relates to whether a current measure is appropriate or whether a measure of past exposure is needed.

Reliability refers to the processes included in an epidemiological study to check repeatability of measurements of the exposures. These usually include intra-observer reliability and inter-observer reliability.

### 4. Were confounding factors identified?

Confounding has occurred where the estimated intervention exposure effect is biased by the presence of some difference between the comparison groups (apart from the exposure investigated/of interest). Typical confounders include baseline characteristics, prognostic factors, or concomitant exposures (e.g. smoking). A confounder is a difference between the comparison groups and it influences the direction of the study results. A high quality study at the level of cohort design will identify the potential confounders and measure them (where possible). This is difficult for studies where behavioral, attitudinal or lifestyle factors may impact on the results.

### 5. Were strategies to deal with confounding factors stated?

Strategies to deal with effects of confounding factors may be dealt within the study design or in data analysis. By matching or stratifying sampling of participants, effects of confounding factors can be adjusted for. When dealing with adjustment in data analysis, assess the statistics used in the study. Most will be some form of multivariate regression analysis to account for the confounding factors measured. Look out for a description of statistical methods as regression methods such as logistic regression are usually employed to deal with confounding factors/variables of interest.

### 6. Were the groups/participants free of the outcome at the start of the study (or at the moment of exposure)?

The participants should be free of the outcomes of interest at the start of the study. Refer to the ‘methods’ section in the paper for this information, which is usually found in descriptions of participant/sample recruitment, definitions of variables, and/or inclusion/exclusion criteria.

### 7. Were the outcomes measured in a valid and reliable way?

Read the methods section of the paper. If for e.g. lung cancer is assessed based on existing definitions or diagnostic criteria, then the answer to this question is likely to be yes. If lung cancer is assessed using observer reported, or self-reported scales, the risk of over- or under-reporting is increased, and objectivity is compromised. Importantly, determine if the measurement tools used were validated instruments as this has a significant impact on outcome assessment validity.

Having established the objectivity of the outcome measurement (e.g. lung cancer) instrument, it’s important to establish how the measurement was conducted. Were those involved in collecting data trained or educated in the use of the instrument/s? (e.g. radiographers). If there was more than one data collector, were they similar in terms of level of education, clinical or research experience, or level of responsibility in the piece of research being appraised?

### 8. Was the follow up time reported and sufficient to be long enough for outcomes to occur?

The appropriate length of time for follow up will vary with the nature and characteristics of the population of interest and/or the intervention, disease or exposure. To estimate an appropriate duration of follow up, read across multiple papers and take note of the range for duration of follow up. The opinions of experts in clinical practice or clinical research may also assist in determining an appropriate duration of follow up. For example, a longer timeframe may be needed to examine the association between occupational exposure to asbestos and the risk of lung cancer. It is important, particularly in cohort studies that follow up is long enough to enable the outcomes. However, it should be remembered that the research question and outcomes being examined would probably dictate the follow up time.

### 9. Was follow up complete, and if not, were the reasons to loss to follow up described and explored?

It is important in a cohort study that a greater percentage of people are followed up. As a general guideline, at least 80% of patients should be followed up. Generally a dropout rate of 5% or less is considered insignificant. A rate of 20% or greater is considered to significantly impact on the validity of the study. However, in observational studies conducted over a lengthy period of time a higher dropout rate is to be expected. A decision on whether to include or exclude a study because of a high dropout rate is a matter of judgement based on the reasons why people dropped out, and whether dropout rates were comparable in the exposed and unexposed groups.

Reporting of efforts to follow up participants that dropped out may be regarded as an indicator of a well conducted study. Look for clear and justifiable description of why people were left out, excluded, dropped out etc. If there is no clear description or a statement in this regards, this will be a 'No'.

### 10. Were strategies to address incomplete follow up utilized?

Some people may withdraw due to change in employment or some may die; however, it is important that their outcomes are assessed. Selection bias may occur as a result of incomplete follow up. Therefore, participants with unequal follow up periods must be taken into account in the analysis, which should be adjusted to allow for differences in length of follow up periods. This is usually done by calculating rates which use person-years at risk, i.e. considering time in the denominator.

### 11. Was appropriate statistical analysis used?

As with any consideration of statistical analysis, consideration should be given to whether there was a more appropriate alternate statistical method that could have been used. The methods section of cohort studies should be detailed enough for reviewers to identify which analytical techniques were used (in particular, regression or stratification) and how specific confounders were measured.

For studies utilizing regression analysis, it is useful to identify if the study identified which variables were included and how they related to the outcome. If stratification was the analytical approach used, were the strata of analysis defined by the specified variables? Additionally, it is also important to assess the appropriateness of the analytical strategy in terms of the assumptions associated with the approach as differing methods of analysis are based on differing assumptions about the data and how it will respond.
