## Supplementary File 5 for "Vertebral Augmentation for Symptomatic Vertebral Hemangiomas: A Systematic Review and Meta-analysis of Pain Relief, Cement Leakage, and Recurrence": Li C.2016.docx

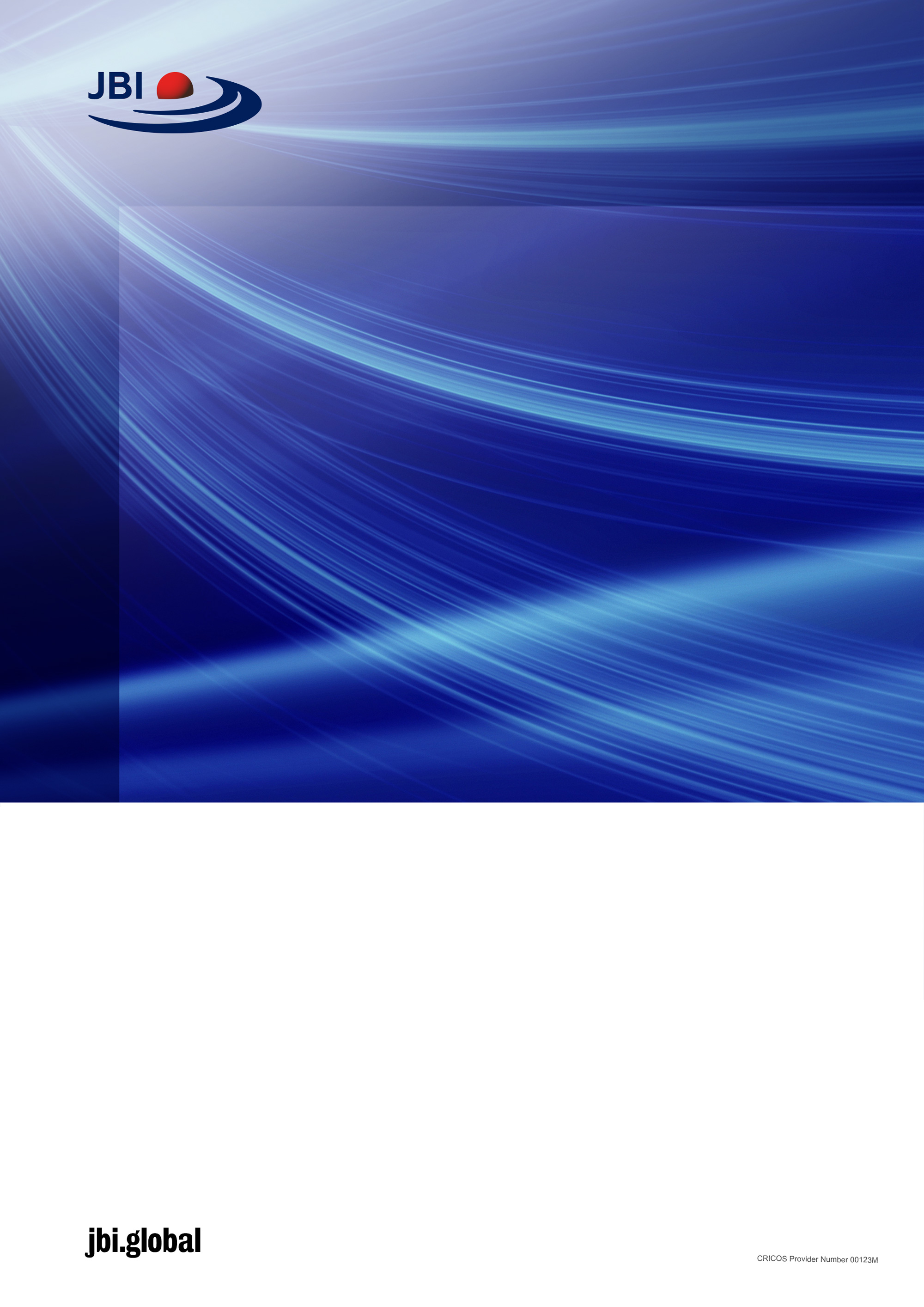

checklist for randomized controlled trials

Critical Appraisal tools for use in JBI Systematic Reviews

Introduction

JBI is an international research organisation based in the Faculty of Health and Medical Sciences at the University of Adelaide, South Australia. JBI develops and delivers unique evidence-based information, software, education and training designed to improve healthcare practice and health outcomes. With over 70 Collaborating Entities, servicing over 90 countries, JBI is a recognised global leader in evidence-based healthcare.

### JBI Systematic Reviews

The core of evidence synthesis is the systematic review of literature of a particular intervention, condition or issue. The systematic review is essentially an analysis of the available literature (that is, evidence) and a judgment of the effectiveness or otherwise of a practice, involving a series of complex steps. JBI takes a particular view on what counts as evidence and the methods utilised to synthesise those different types of evidence. In line with this broader view of evidence, JBI has developed theories, methodologies and rigorous processes for the critical appraisal and synthesis of these diverse forms of evidence in order to aid in clinical decision-making in healthcare. There now exists JBI guidance for conducting reviews of effectiveness research, qualitative research, prevalence/incidence, etiology/risk, economic evaluations, text/opinion, diagnostic test accuracy, mixed-methods, umbrella reviews and scoping reviews. Further information regarding JBI systematic reviews can be found in the [JBI Evidence Synthesis Manual](https://jbi-global-wiki.refined.site/space/MANUAL).

### JBI Critical Appraisal Tools

All systematic reviews incorporate a process of critique or appraisal of the research evidence. The purpose of this appraisal is to assess the methodological quality of a study and to determine the extent to which a study has addressed the possibility of bias in its design, conduct and analysis. All papers selected for inclusion in the systematic review (that is – those that meet the inclusion criteria described in the protocol) need to be subjected to rigorous appraisal by two critical appraisers. The results of this appraisal can then be used to inform synthesis and interpretation of the results of the study. JBI Critical appraisal tools have been developed by the JBI and collaborators and approved by the JBI Scientific Committee following extensive peer review. Although designed for use in systematic reviews, JBI critical appraisal tools can also be used when creating Critically Appraised Topics (CAT), in journal clubs and as an educational tool.

JBI Critical Appraisal Checklist for
randomized Controlled trials

Reviewer	__Farzin MohammadMoradi_______________________________ Date______________13^th^ July_________________

Author____________________Li C. Year____2016_____ Record Number____1_____

|  | Yes | No | Unclear | NA |
| --- | --- | --- | --- | --- |
| 1. Was true randomization used for assignment of participants to treatment groups? | □ | □ | □ | □ |
| 2. Was allocation to treatment groups concealed? | □ | □ | □ | □ |
| 3. Were treatment groups similar at the baseline? | □ | □ | □ | □ |
| 4. Were participants blind to treatment assignment? | □ | □ | □ | □ |
| 5. Were those delivering treatment blind to treatment assignment? | □ | □ | □ | □ |
| 6. Were outcomes assessors blind to treatment assignment? | □ | □ | □ | □ |
| 7. Were treatment groups treated identically other than the intervention of interest? | □ | □ | □ | □ |
| 8. Was follow up complete and if not, were differences between groups in terms of their follow up adequately described and analyzed? | □ | □ | □ | □ |
| 9. Were participants analyzed in the groups to which they were randomized? | □ | □ | □ | □ |
| 10. Were outcomes measured in the same way for treatment groups? | □ | □ | □ | □ |
| 11. Were outcomes measured in a reliable way? | □ | □ | □ | □ |
| 12. Was appropriate statistical analysis used? | □ | □ | □ | □ |
| 13. Was the trial design appropriate, and any deviations from the standard RCT design (individual randomization, parallel groups) accounted for in the conduct and analysis of the trial? | □ | □ | □ | □ |

Overall appraisal:	Include	□	Exclude	□	Seek further info □

Comments (Including reason for exclusion)

_______________________LOW RISK (75%)

Explanation for the critical appraisal tool
for RCTs with individual participants in
parallel groups

How to cite: *Tufanaru C, Munn Z, Aromataris E, Campbell J, Hopp L. Chapter 3: Systematic reviews of effectiveness. In: Aromataris E, Munn Z (Editors)*. JBI Manual for Evidence Synthesis. JBI, 2020. Available from <https://synthesismanual.jbi.global>

Answers: Yes, No, Unclear or Not/Applicable

**Critical Appraisal Tool for RCTs (individual participants in parallel groups)**

1. **Was true randomization used for assignment of participants to treatment groups?**

The differences between participants included in compared groups constitutes a threat to the internal validity of a study exploring causal relationships. If participants are not allocated to treatment and control groups by random assignment there is a risk that the allocation is influenced by the known characteristics of the participants and these differences between the groups may distort the comparability of the groups. A true random assignment of participants to the groups means that a procedure is used that allocates the participants to groups purely based on chance, not influenced by the known characteristics of the participants. Check the details about the randomization procedure used for allocation of the participants to study groups. Was a true chance (random) procedure used? For example, was a list of random numbers used? Was a computer-generated list of random numbers used?

2. **Was allocation to groups concealed?**

If those allocating participants to the compared groups are aware of which group is next in the allocation process, that is, treatment or control, there is a risk that they may deliberately and purposefully intervene in the allocation of patients by preferentially allocating patients to the treatment group or to the control group and therefore this may distort the implementation of allocation process indicated by the randomization and therefore the results of the study may be distorted. Concealment of allocation (allocation concealment) refers to procedures that prevent those allocating patients from knowing before allocation which treatment or control is next in the allocation process. Check the details about the procedure used for allocation concealment. Was an appropriate allocation concealment procedure used? For example, was central randomization used? Were sequentially numbered, opaque and sealed envelopes used? Were coded drug packs used?

3. **Were treatment groups similar at the baseline?**

The differences between participants included in compared groups constitute a threat to the internal validity of a study exploring causal relationships. If there are differences between participants included in compared groups there is a risk of selection bias. If there are differences between participants included in the compared groups maybe the ‘effect’ cannot

be attributed to the potential ‘cause’ (the examined intervention or treatment), as maybe it is plausible that the ‘effect’ may be explained by the differences between participants, that is, by selection bias. Check the characteristics reported for participants. Are the participants from the compared groups similar with regards to the characteristics that may explain the effect even in the absence of the ‘cause’, for example, age, severity of the disease, stage of the disease, co-existing conditions and so on? Check the proportions of participants with specific relevant characteristics in the compared groups. Check the means of relevant measurements in the compared groups (pain scores; anxiety scores; etc.). *[Note: Do NOT only consider the P-value for the statistical testing of the differences between groups with regards to the baseline characteristics.]*

4. **Were participants blind to treatment assignment?**

If participants are aware of their allocation to the treatment group or to the control group there is the risk that they may behave differently and respond or react differently to the intervention of interest or to the control intervention respectively compared to the situations when they are not aware of treatment allocation and therefore the results of the study may be distorted. Blinding of participants is used in order to minimize this risk. Blinding of the participants refers to procedures that prevent participants from knowing which group they are allocated. If blinding of participants is used, participants are not aware if they are in the group receiving the treatment of interest or if they are in any other group receiving the control interventions. Check the details reported in the article about the blinding of participants with regards to treatment assignment. Was an appropriate blinding procedure used? For example, were identical capsules or syringes used? Were identical devices used? Be aware of different terms used, blinding is sometimes also called masking.

5. **Were those delivering treatment blind to treatment assignment?**

If those delivering treatment are aware of participants’ allocation to the treatment group or to the control group there is the risk that they may behave differently with the participants from the treatment group and the participants from the control group, or that they may treat them differently, compared to the situations when they are not aware of treatment allocation and this may influence the implementation of the compared treatments and the results of the study may be distorted. Blinding of those delivering treatment is used in order to minimize this risk. Blinding of those delivering treatment refers to procedures that prevent those delivering treatment from knowing which group they are treating, that is those delivering treatment are not aware if they are treating the group receiving the treatment of interest or if they are treating any other group receiving the control interventions. Check the details reported in the article about the blinding of those delivering treatment with regards to treatment assignment. Is there any information in the article about those delivering the treatment? Were those delivering the treatment unaware of the assignments of participants to the compared groups?

6. **Were outcomes assessors blind to treatment assignment?**

If those assessing the outcomes are aware of participants’ allocation to the treatment group or to the control group there is the risk that they may behave differently with the participants from the treatment group and the participants from the control group compared to the situations when they are not aware of treatment allocation and therefore there is the risk that the measurement of the outcomes may be distorted and the results of the study may be distorted. Blinding of outcomes assessors is used in order to minimize this risk. Check the details reported in the article about the blinding of outcomes assessors with regards to treatment assignment. Is there any information in the article about outcomes assessors? Were those assessing the treatment’s effects on outcomes unaware of the assignments of participants to the compared groups?

7. **Were treatment groups treated identically other than the intervention of interest?**

In order to attribute the ‘effect’ to the ‘cause’ (the treatment or intervention of interest), assuming that there is no selection bias, there should be no other difference between the groups in terms of treatment or care received, other than the manipulated ‘cause’ (the treatment or intervention controlled by the researchers). If there are other exposures or treatments occurring at the same time with the ‘cause’ (the treatment or intervention of interest), other than the ‘cause’, then potentially the ‘effect’ cannot be attributed to the examined ‘cause’ (the investigated treatment), as it is plausible that the ‘effect’ may be explained by other exposures or treatments occurring at the same time with the ‘cause’ (the treatment of interest). Check the reported exposures or interventions received by the compared groups. Are there other exposures or treatments occurring at the same time with the ‘cause’? Is it plausible that the ‘effect’ may be explained by other exposures or treatments occurring at the same time with the ‘cause’? Is it clear that there is no other difference between the groups in terms of treatment or care received, other than the treatment or intervention of interest?

**8.	Was follow up complete and if not, were differences between groups in terms of their follow up adequately described and analyzed?**

For this question, follow up refers to the time period from the moment of random allocation (random assignment or randomization) to compared groups to the end time of the trial. This critical appraisal question asks if there is complete knowledge (measurements, observations etc.) for the entire duration of the trial as previously defined (that is, from the moment of random allocation to the end time of the trial), for all randomly allocated participants. If there is incomplete follow up, that is incomplete knowledge about all randomly allocated participants, this is known in the methodological literature as the post-assignment attrition. As RCTs are not perfect, there is almost always post-assignment attrition, and the focus of this question is on the appropriate exploration of post-assignment attrition (description of loss to follow up, description of the reasons for loss to follow up, the estimation of the impact of loss

to follow up on the effects etc.). If there are differences with regards to the loss to follow up between the compared groups in an RCT, these differences represent a threat to the internal validity of a randomized experimental study exploring causal effects, as these differences may provide a plausible alternative explanation for the observed ‘effect’ even in the absence of the ‘cause’ (the treatment or intervention of interest). When appraising an RCT, check if there were differences with regards to the loss to follow up between the compared groups. If follow up was incomplete (that is, there is incomplete information on all participants), examine the reported details about the strategies used in order to address incomplete follow up, such as descriptions of loss to follow up (absolute numbers; proportions; reasons for loss to follow up) and impact analyses (the analyses of the impact of loss to follow up on results). Was there a description of the incomplete follow up (number of participants and the specific reasons for loss to follow up)? It is important to note that with regards to loss to follow up, it is not enough to know the number of participants and the proportions of participants with incomplete data; the reasons for loss to follow up are essential in the analysis of risk of bias; even if the numbers and proportions of participants with incomplete data are similar or identical in compared groups, if the patterns of reasons for loss to follow up are different (for example, side effects caused by the intervention of interest, lost contact etc.), these may impose a risk of bias if not appropriately explored and considered in the analysis. If there are differences between groups with regards to the loss to follow up (numbers/proportions and reasons), was there an analysis of patterns of loss to follow up? If there are differences between the groups with regards to the loss to follow up, was there an analysis of the impact of the loss to follow up on the results? [Note: Question 8 is NOT about intention-to-treat (ITT) analysis; question 9 is about ITT analysis.]

**9. Were participants analyzed in the groups to which they were randomized?**

This question is about the intention-to-treat (ITT) analysis. There are different statistical analysis strategies available for the analysis of data from randomized controlled trials, such as intention-to-treat analysis (known also as intent to treat; abbreviated, ITT), per-protocol analysis, and as-treated analysis. In the ITT analysis the participants are analyzed in the groups to which they were randomized, regardless of whether they actually participated or not in those groups for the entire duration of the trial, received the experimental intervention or control intervention as planned or whether they were compliant or not with the planned experimental intervention or control intervention. The ITT analysis compares the outcomes for participants from the initial groups created by the initial random allocation of participants to those groups. Check if ITT was reported; check the details of the ITT. Were participants analyzed in the groups to which they were initially randomized, regardless of whether they actually participated in those groups, and regardless of whether they actually received the planned interventions? *[Note: The ITT analysis is a type of statistical analysis recommended in the Consolidated Standards of Reporting Trials (CONSORT) statement on best practices in trials reporting, and it is considered a marker of good methodological quality of the analysis of results of a randomized trial. The ITT is estimating the effect of offering the intervention, that is, the effect of instructing the participants to use or take the intervention; the ITT it is not estimating the effect of actually receiving the intervention of interest.]*

**10. Were outcomes measured in the same way for treatment groups?**

If the outcome (the ‘effect’) is not measured in the same way in the compared groups there is a threat to the internal validity of a study exploring a causal relationship as the differences in outcome measurements may be confused with an effect of the treatment (the ‘cause’). Check if the outcomes were measured in the same way. Same instrument or scale used? Same measurement timing? Same measurement procedures and instructions?

**11. Were outcomes measured in a reliable way?**

Unreliability of outcome measurements is one threat that weakens the validity of inferences about the statistical relationship between the ‘cause’ and the ‘effect’ estimated in a study exploring causal effects. Unreliability of outcome measurements is one of the different plausible explanations for errors of statistical inference with regards to the existence and the magnitude of the effect determined by the treatment (‘cause’). Check the details about the reliability of measurement such as the number of raters, training of raters, the intra-rater reliability, and the inter-raters reliability within the study (not as reported in external sources). This question is about the reliability of the measurement performed in the study, it is not about the validity of the measurement instruments/scales used in the study. *[Note: Two other important threats that weaken the validity of inferences about the statistical relationship between the ‘cause’ and the ‘effect’ are low statistical power and the violation of the assumptions of statistical tests. These other two threats are explored within Question 12).]*

**12. Was appropriate statistical analysis used?**

Inappropriate statistical analysis may cause errors of statistical inference with regards to the existence and the magnitude of the effect determined by the treatment (‘cause’). Low statistical power and the violation of the assumptions of statistical tests are two important threats that weaken the validity of inferences about the statistical relationship between the ‘cause’ and the ‘effect’. Check the following aspects: if the assumptions of statistical tests were respected; if appropriate statistical power analysis was performed; if appropriate effect sizes were used; if appropriate statistical procedures or methods were used given the number and type of dependent and independent variables, the number of study groups, the nature of the relationship between the groups (independent or dependent groups), and the objectives of statistical analysis (association between variables; prediction; survival analysis etc.).

**13. Was the trial design appropriate for the topic, and any deviations from the standard RCT design accounted for in the conduct and analysis?**

Certain RCT designs, such as the crossover RCT, should only be conducted when appropriate. Alternative designs may also present additional risks of bias if not accounted for in the design and analysis.

Crossover trials should only be conducted in people with a chronic, stable condition, where the intervention produces a short term effect (i.e. relief in symptoms). Crossover trials should ensure there is an appropriate period of washout between treatments.

Cluster RCTs randomize groups of individuals, forming ‘clusters.’ When we are assessing outcomes on an individual level in cluster trials, there are unit-of-analysis issues, as individuals within a cluster are correlated. This should be taken into account by the study authors when conducting analysis, and ideally authors will report the intra-cluster correlation coefficient.

Stepped-wedge RCTs may be appropriate when it is expected the intervention will do more good than harm, or due to logistical, practical or financial considerations in the roll out of a new treatment/intervention. Data analysis in these trials should be conducted appropriately, taking into account the effects of time.
